# Harnessing non-standard nucleic acids for highly sensitive icosaplex (20-plex) detection of microbial threats

**DOI:** 10.1101/2024.09.09.24313328

**Authors:** Hinako Kawabe, Luran Manfio, Sebastian Magana Pena, Nicolette A. Zhou, Kevin M. Bradley, Cen Chen, Chris McLendon, Steven A. Benner, Karen Levy, Zunyi Yang, Jorge A. Marchand, Erica R. Fuhrmeister

## Abstract

Environmental surveillance and clinical diagnostics heavily rely on the polymerase chain reaction (PCR) for target detection. A growing list of microbial threats warrants new PCR-based detection methods that are highly sensitive, specific, and multiplexable. Here, we introduce a PCR-based icosaplex (20-plex) assay for detecting 18 enteropathogen and two antimicrobial resistance genes. This multiplexed PCR assay leverages the self-avoiding molecular recognition system (SAMRS) to avoid primer dimer formation, the artificially expanded genetic information system (AEGIS) for amplification specificity, and next-generation sequencing for amplicon identification. We benchmarked this assay using a low-cost, portable sequencing platform (Oxford Nanopore) on wastewater, soil, and human stool samples. Using parallelized multi-target TaqMan Array Cards (TAC) to benchmark performance of the 20-plex assay, there was 74% agreement on positive calls and 97% agreement on negative calls. Additionally, we show how sequencing information from the 20-plex can be used to further classify allelic variants of genes and distinguish sub-species. The strategy presented offers sensitive, affordable, and robust multiplex detection that can be used to support efforts in wastewater-based epidemiology, environmental monitoring, and human/animal diagnostics.

## Introduction

Emerging infectious diseases, coupled with rising antibiotic resistance, are a threat to global public health.^1^ The diversity of pathogens that can cause illness necessitates pathogen detection methods that can identify multiple genetic targets. There is a need for low-cost, multi-target, detection methods, especially in regions where the burden of infectious diseases is high, resources are constrained, and there is a high diversity in the pathogens that are present.

A high burden of diarrheal illness exists in low- and middle-income countries (LMICs).^2^ The types of enteric pathogens (bacteria, virus, protozoa, helminths) present in LMICs contributing to disease are geographically diverse and location specific.^3–5^ As an example of geographic diversity, previous studies used highly parallelized quantitative polymerase chain reaction (qPCR) to survey a wide range of pathogens around the world.^6^ In Mozambique, *Shigella* spp. and *Giardia* spp. were the most prevalent pathogens whereas *Campylobacter* spp. and *Giardia* spp. dominated infections in children across eight other settings in South America, sub-Saharan Africa, and Asia.^3,5^

The need for multi-target detection assays is not limited to LMICs. In high-income countries (HICs), respiratory illnesses are common and diarrheal illnesses predominate through foodborne outbreaks.^7,8^ Characteristically, urban areas in HICs have sewered sanitation systems, which allow for active monitoring of the infectious disease burden through wastewater-based epidemiology (WBE).^9^ As an early example, Poliovirus was isolated from wastewater samples as part of the World Health Organization’s Global Polio Eradication Initiative (GPEI) to monitor for emergence/reemergence of the virus.^10–12^ Recently, SARS-CoV-2 was monitored in wastewater at ≈14,000 sites in 59 countries in March 2021.^13^ These latest efforts rely on methods such as qPCR and digital PCR (dPCR) to determine the level of infections in a population.^14–16^ Looking beyond the COVID-19 pandemic, monitoring programs are eager to expand surveillance to include enteric pathogens, other respiratory viruses, sexually transmitted infections, arboviruses, and antimicrobial resistance.^17^ However, expanding the range of targets for surveillance is both labor-intensive and costly, underscoring the urgent need for new multi-target capabilities that can efficiently and affordably address this challenge.

Complementing WBE efforts, other environmental surveillance strategies are premised on the environment (e.g., soil, water, air, fomites) serving as an intermediary between infected hosts.^18^ Environmental detection can facilitate surveying disease burden. For example, a study in Kenya found that positive detection of helminths (*Ascaris lumbricoides*, *Trichuris trichiura*, and *Necator americanus*) in a household’s soil was significantly associated with cases of helminth infections of household members.^19^ As a result, qPCR-based surveillance of helminths in soil is a promising alternative to more invasive stool-based surveillance.

Environmental detection is also used for determining dominant transmission pathways of pathogens. During the COVID-19 pandemic, studies sampled fomites using qPCR for SARS-CoV-2 RNA and concluded that fomites were unlikely to be a dominant transmission pathway.^20,21^ Similar methods have been proposed to survey the burden of antimicrobial resistance across the globe by sampling soil and water.^22–24^ To expand the scale and scope of environmental surveillance, highly multiplexed detection assays that can capture the diversity of possible microbial threats of interest are needed. Despite this pressing need, methods for multi-target detection generally fall short in achieving meaningful reductions in assay cost, necessitating a more selective approach in deciding which targets to prioritize in monitoring, surveillance, and detection efforts.

PCR-based multi-target detection strategies are typically limited by the number of targets that can be simultaneously amplified and identified. In conventional multiplexed PCR amplification, increasing the number of targets increases the likelihood of off-target reactions (e.g., primer dimer formation, non-specific amplification product). These off-target reactions result in reduced sensitivity or assay failure.^25^ Further, highly multiplexed PCR assays tend to have a narrow tolerance for changing reaction conditions and sample composition. For example, adding primers for new targets to an established multiplexed assay can result in primer cascading failure. Performing an assay in highly heterogeneous sample matrices, such as environmental DNA extracts, can also reduce PCR efficiency and result in false negatives. Additionally, multiplexed fluorescence-based detection assays, such as qPCR, are typically constrained (up to 5 targets) by the limited orthogonality of fluorescent reporter dye spectra.^26^

Other detection technologies have approached the ‘many-target’ problem by scaling down reaction volumes and parallelizing reactions in microfluidic devices.^27^ One widely used commercial platform, the TaqMan^TM^ Array Card (TAC), parallelizes qPCR reactions into micro-scale (≈1.5 µL) reactions.^28^ Even with this compartmentalization, qPCR and dPCR-based platforms still encounter significant challenges in scaling and accessibility due to the high capital costs of equipment, high variable costs for consumables, and the substantial cost and personnel time required for adding additional targets. To achieve highly multiplexed detection, beyond what is accessible with qPCR and dPCR, alternative solutions are needed.

Research in synthetic biology has produced various non-standard nucleotides that can be used to circumvent major obstacles associated with scaling multiplexed PCR amplification to larger ‘*n*’-plex reactions (**Figure 1**). The non-standard nucleic acids from the Self-Avoiding Molecular Recognition Systems (SAMRS: A*, T*, G*, and C*) are structurally modified versions of the standard DNA nucleobases (A, T, G, and C).^29^ Though structurally distinct, SAMRS nucleobases maintain an ability to base pair with standard DNA nucleobases (**Figure 1c**) but not with their SAMRS complement. For nucleic acid amplification, primers modified with SAMRS components anneal and amplify natural DNA/RNA. Conversely, formation of SAMRS:SAMRS pairs (T*:A* and C*:G*, **Figure 1d**) are thermodynamically disfavored. In amplification assays with SAMRS-containing primers, this results in a decrease in off-target primer-primer interactions. Selectively inserting SAMRS bases into primer sequences has been shown to reduce primer dimer formation and increase multiplexed assay sensitivity.^30,31^

**Figure 1.**
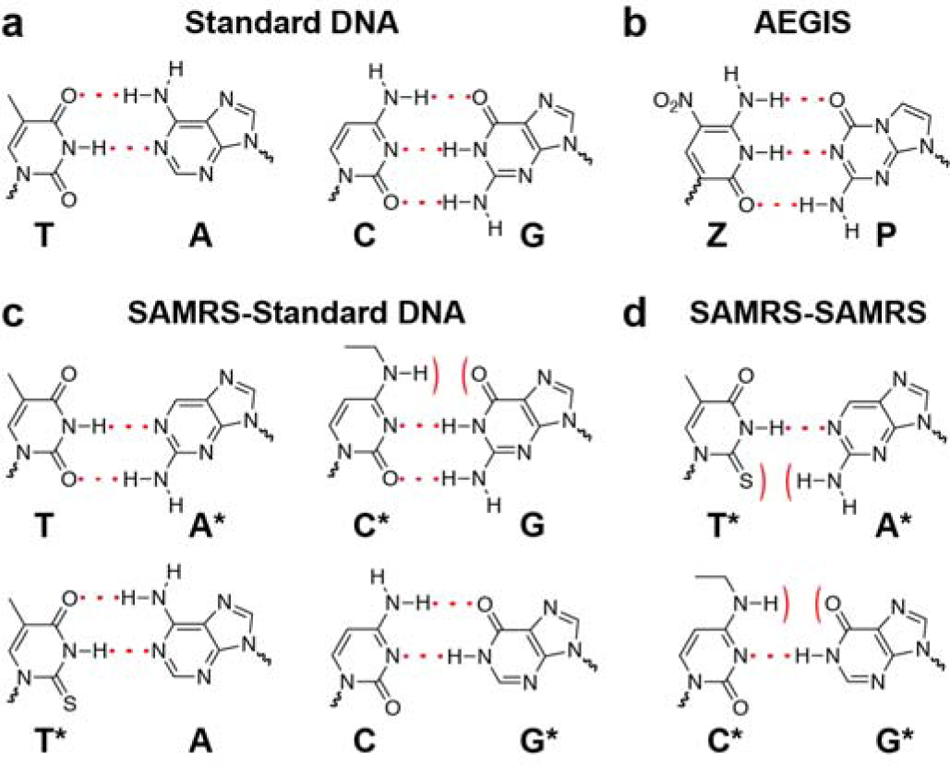
Structures and hydrogen-bonding interactions for standard DNA, AEGIS, and SAMRS nucleobases. (**a**) Structures of standard DNA hydrogen bonding base pairs. (**b**) Structures of Z and P AEGIS bases that form base pairs orthogonal to the standard DNA bases. (**c**) SAMRS bases (*) form base pairs with their natural standard DNA complements. (**d**) The strategic removal of hydrogen bonding groups hinders SAMRS bases from base pairing with their SAMRS complement. Dotted lines indicate hydrogen bonding between base pairs, and curved lines indicate a lack of hydrogen bond formation between base pairs.

Additionally, non-standard nucleic acids from the Artificially Expanded Genetic Information System (AEGIS) can be used to improve primer binding specificity.^32^ AEGIS nucleotides (Z:P pair, **Figure 1b**), such as Z (6-amino-5-nitro-3-(1′-β-d-2′-deoxyribofuranosyl)-2(1H)pyridine) and P (2-amino-8-(1′-β-d-2′-deoxyribofuranosyl), can form highly-specific base pairs *orthogonal* to the standard, natural set (T:A and C:G).^33,34^ Since AEGIS bases are not found in nature, primers containing AEGIS nucleotides can be used to amplify template targets containing complementary AEGIS sequences while avoiding off-target amplification.^35^

Various detection assays have been developed that leverage properties of SAMRS and AEGIS bases for viral pathogen detection, including arboviruses (e.g., Zika, Dengue, Chikungyunya),^36,37^ coronaviruses (e.g., RSV, MERS-CoV, Influenza A/B, SARS-CoV-2),^38,39^ human papillomavirus (HPV),^40^ and norovirus.^41^ Despite proving utility of SAMRS and AEGIS nucleobases in assay design, these assays were either non-multiplexed or multiplexed but required an expensive strategy for target readout (XMAP Luminex array detection).

In this work, we develop an ‘*icosaplex*’ (20-plex) PCR-based sequencing assay able to detect 20 enteric pathogen and antimicrobial resistance gene targets. This 20-plex assay greatly expands on prior work that targeted viruses to new pathogens (bacteria, protozoa, and helminths) and to environmental sample types. The 20-plex assay amalgamates individual primer sets for 20 targets from a previous collection of work and achieves effective multiplexing by incorporating SAMRS-AEGIS nucleotides into primers chosen for biological reasons. To circumvent the target identification limitations of PCR, we leveraged nanopore sequencing (Oxford Nanopore Technologies), a third-generation sequencing method that is inexpensive, portable, and can provide sequencing results in real time. This study is the first to use SAMRS-AEGIS primers for highly multiplexed PCR in combination with nanopore sequencing for microbial surveillance applications. Sequencing information provides additional insight into gene alleles and subspecies that would otherwise be missed through presence/absence methods. The target panel and detection method were chosen for application areas in environmental detection and surveillance efforts in resource constrained settings, such as LMICs. We benchmarked performance of the 20-plex assay in three sample matrices: wastewater, soil, and human feces.

## Results and Discussion

### Pathogen and Antimicrobial Resistance Gene Target Selection

We developed a multiplexable assay that can detect a broad range of microbial threats relevant to global health. We chose 20 genes of interest that encompass a wide range of enteropathogens (bacteria, protozoa, and one helminth) and two clinically important antimicrobial resistance genes (ARGs) (**Table 1**). QPCR assays for these 20 targets have previously been reported (**Table S1)**.

**Table 1.**
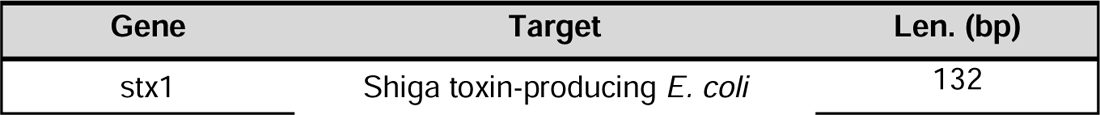
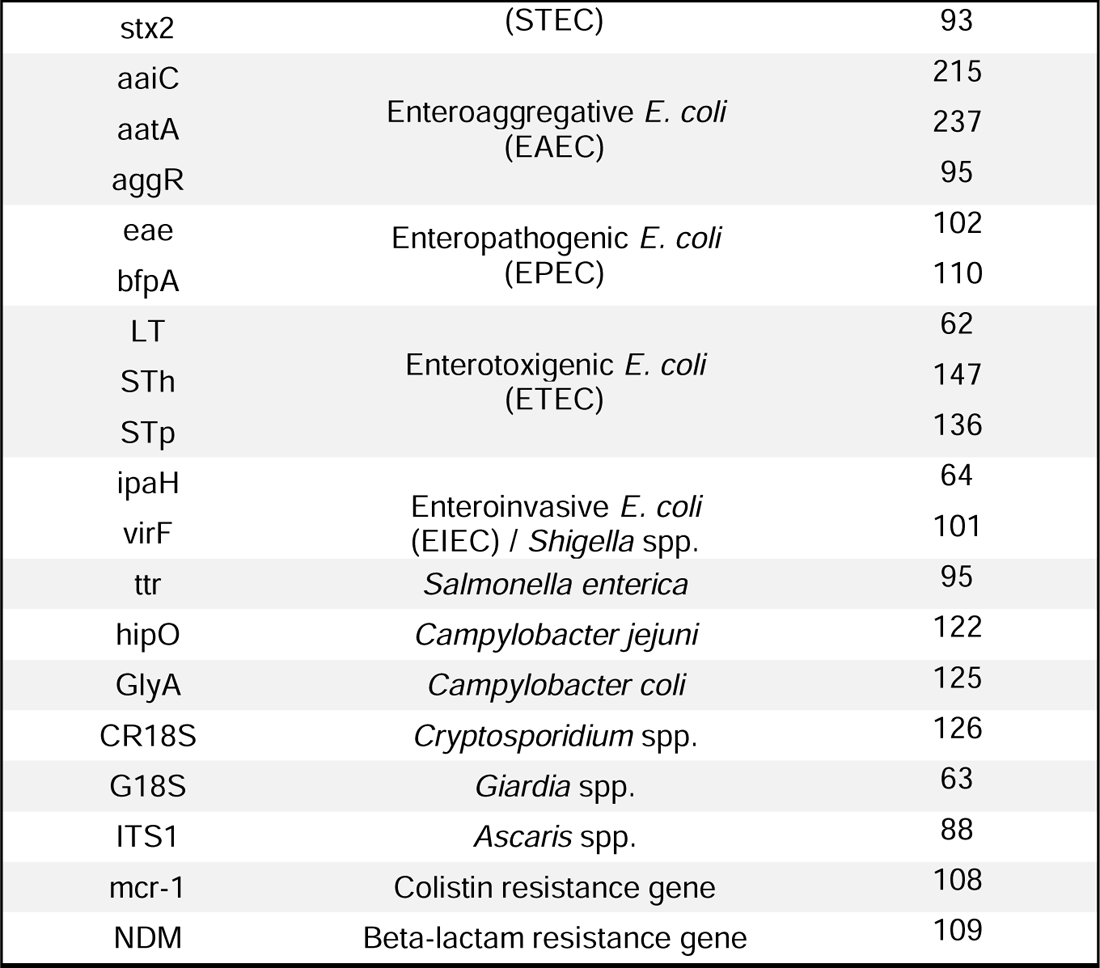
Enteric pathogen and antimicrobial resistance gene panel in the 20-plex assay. The 20-plex assay is designed to detect 18 enteropathogen genes and two antimicrobial resistance genes. Amplicon lengths for each PCR product are reported.

Twelve of our targets are genes specific to pathogenic *E. coli.* These represent five of the major pathogenic *E. coli* subtypes and *Shigella* spp. In LMICs, pathogenic *E. coli* is a leading cause of diarrheal illness.^5,42,43^ The enterotoxigenic *E. coli* (ETEC) subtype is associated with moderate-to-severe diarrhea which can lead to additional severe clinical outcomes.^43^ LMICs also experience a high incidence of soil-transmitted helminth infections.^44^ It is estimated that 738 million people globally are infected with helminths of the genus *Ascaris*.^45^ In HICs like the United States, *Campylobacter* spp., non-typhoidal *Salmonella*, *Shigella* spp., and *Giardia intestinalis*, are leading causes of reported foodborne illnesses.^46^ In both LMICs and HICs, pathogenic bacteria pose an even larger threat to human health if they acquire antimicrobial resistance activity. *Bla*_NDM_ and *mcr-1* are globally distributed ARGs that confer resistance to the last line of defense antibiotics reserved for difficult-to-treat infections.^47,48^ Many of the 20 gene targets chosen in our panel were used by other studies to detect pathogens and ARGs in human feces,^3,6^ wastewater,^49^ and environmental samples. ^50–52^

### Design and validation of 20-plex primers

Primer sequences from previously reported PCR and qPCR assays for the 20 gene targets were used as a starting point for 20-plex PCR primer design (**Table S1**). Initial 40 primer sequences (1 forward, 1 reverse for each gene target) were chosen to accommodate a single annealing temperature (60 °C) during PCR cycling. At these temperatures and at a high relative abundance of primer to target, various cross-primer interactions can occur to form primer dimers and off-target amplicons, each reducing assay sensitivity (Figure 2a). To combat primer dimer formation (Figure 2b), we modified all 40 standard DNA primers with SAMRS nucleobases using the PrimerCompare software developed at the Foundation for Applied Molecular Evolution (FfAME). PrimerCompare took standard DNA primer sequences that have proven targets, primer concentrations, salt concentrations, and thermodynamic parameters (maximum ΔG for hairpins and dimers) as inputs to simulate potential primer-primer interactions. These interactions include self-dimerization, cross-primer dimerization, and hairpin structures.

**Figure 2.**
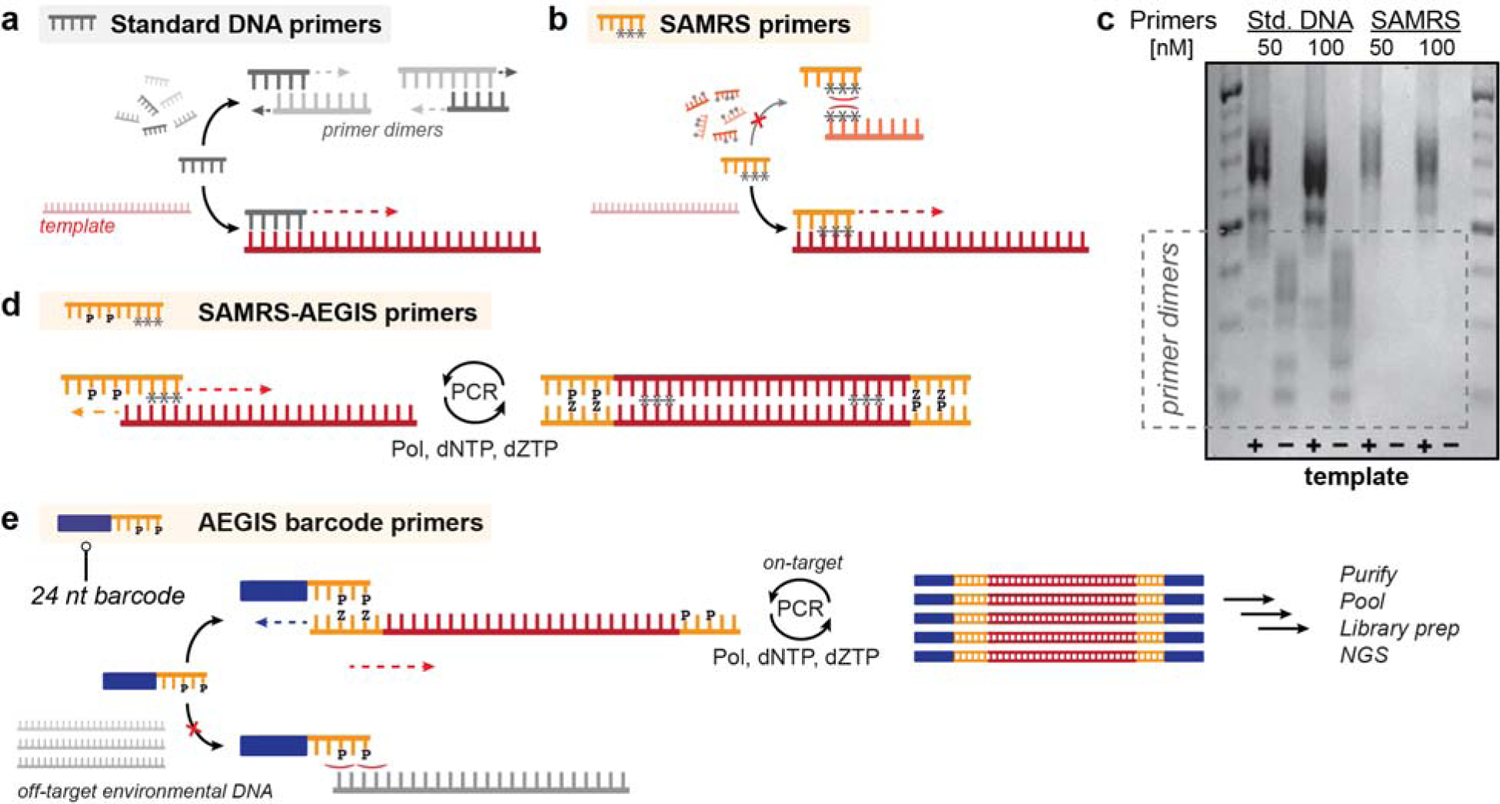
Role of SAMRS and AEGIS nucleotides in multiplex PCR design. (**a**) Standard DNA primers in a PCR reaction can dimerize and cross prime, consuming available primers and dNTPs. (**b**) SAMRS bases can be inserted in primer sequences to avoid primer dimer formation. (**c**) PCR amplification of all 20 targets using synthetic templates in nuclease-free water using standard DNA or SAMRS primers shows standard DNA primer dimerization, particularly prominent when no template is present (**Figure S3, Table S2-S6)**. SAMRS primers show no visible dimerization. (**d**) The first round of amplification in our SAMRS-AEGIS 20-plex reaction uses primers containing SAMRS bases in the target-binding region, and the AEGIS P base in an overhang tag region. The corresponding Z triphosphate (dZTP) is included during amplification. (**e**) The second amplification reaction uses primers containing the AEGIS P base to bind to the tag region added during the first round. These primers also contain 24-nt barcode overhangs. Here, the AEGIS bases prevent non-specific amplification due to their lack of pairing with standard DNA bases. After this second PCR amplification, samples are purified, pooled, and prepared for next-generation sequencing.

SAMRS containing primers for the 20 targets (**Table 1**) were then synthesized and validated in single target PCR amplification reactions (**Figure S1)**. A qPCR melt curve for each primer set, with and without added synthetic DNA template, showed no primer dimer formations in the no template controls (NTC, **Figure S2**). We then tested the effectiveness of the SAMRS primers compared to standard DNA primers at reducing primer dimers in multiplexed reaction conditions (**Table S2-S5).** 20-plex PCR was performed with and without synthetic template added (**Table S6**). By agarose gel electrophoresis, primer dimer products were observed with standard DNA primers, but not SAMRS-containing primers (Figure 2c**, Figure S3**). While both standard DNA and SAMRS-containing primers showed target amplification in the 20-plex, the identity of the individual amplicons could not be resolved through gel electrophoresis alone.

While incorporation of SAMRS bases helped decrease primer dimers in multiplex PCR reactions, addition of a 5′-overhang tag to primers could be used for downstream barcoding and attachment of sequencing adapters (**Table S2**). To avoid off-target amplification, 5′-overhang tag sequences should be distinct from sequences that could be present in samples of interest. The generalized design of a 5′-overhang tag, however, is challenging due to the unknown metagenetic composition of many sample matrices (e.g., wastewater, soil, surface water, fomites, feces). We overcame this obstacle by introducing non-standard AEGIS **P** nucleobases into the 5′-overhang tag. Since AEGIS bases form a highly specific orthogonal base pair to the standard DNA bases (**Z:P**, Figure 1b, Figure 2d), AEGIS containing primers should solely bind to complementary AEGIS-tagged regions.

Various design choices were made to minimize design complexity and reagents that would be required for performing multiplexed assays that use AEGIS components. First, the AEGIS tag sequences used a 5-letter alphabet composed of the standard DNA bases (A, T, G, C) and one of the AEGIS bases (P). In amplification reactions, end-users would therefore only need access to complementary nucleotide triphosphate, dZTP, rather than both dZTP and dPTP. Second, we chose to use a single AEGIS tag sequence for both forward and reverse primers. We previously observed that using a single tag sequence in multiplexed PCR reactions reduced overall primer dimer formation and increased detection sensitivity (data not shown). The final AEGIS tag sequence (AGC**P**CTCG**P**TTC) was selected due to low propensity for hairpin formation, as determined computationally. This AEGIS tag sequence is appended to the 5′-end of the 40 SAMRS-containing primers used in this work (**Table S2**).

To multiplex samples, we then created 10 unique barcoding primers that contained a 24-nt barcode region using sequences from an Oxford Nanopore Technologies barcoding kit. These barcoding primers contained the barcode sequence and a downstream region homologous to the common 5′-tag of the 20-plex SAMRS-AEGIS primers (**Table S3**). The universal 5′ AEGIS tag thus serves as the priming region for the barcoding primers either in the same PCR reaction (one-pot amplification) or a subsequent PCR reaction (sequential amplification, Figure 2e). Though barcoding primers discussed in this work were designed to be compatible with Oxford Nanopore demultiplexing workflows, a similar design strategy can be used for barcoding applications on other sequencing platforms (**Table S3**).

### Optimization of a sequential ‘two-step’ 20-plex PCR reaction

A unique challenge of multiplexing in complex samples of unknown metagenomic composition is that gene targets are not present in equimolar amounts. For certain sample types, targets in the same sample could be present at gene copy numbers that vary by orders of magnitude. If barcoding and target amplification occur in one reaction, rather than sequentially, higher abundance targets will bias amplification and consume barcoding primers, reducing assay sensitivity for lower abundance targets. We tested this hypothesis by performing both one-pot and sequential amplification of two targets using synthetic templates, stx2 (present at 10 or 10^2^ copies/µL) and aaiC (10^4^ or 10^5^ copies/µL). When compared to one-pot PCR, performing barcoding in a separate PCR reaction (sequential PCR) decreased differences in abundances of stx2 and aaiC amplicons (**Figure S4**).

Subsequently, PCR optimization was used to identify optimal reaction and cycling conditions. For the optimal number of cycles in each step, we found 40 cycles (as is used for qPCR) during the first round of amplification followed by 15 cycles in the second round for barcoding minimized amplification bias and maximized barcoded targets over other combinations tested (**Figure S4, S5**). In the first round of PCR, we found a uniform concentration of each primer (0.2 µM of each primer, 8 µM total primer) minimized observed amplification bias (**Figure S6**). Under these optimized 20-plex PCR reaction and cycling conditions, primer dimers were still observed with standard DNA primers, but not with SAMRS-AEGIS primers (**Figure S7**).

Finally, we incorporated nanopore sequencing, a low capital cost, portable sequencing platform, as a read-out for detection of the SAMRS-AEGIS 20-plex reaction. Amplification was performed on the 20 synthetic template mixtures at two initial concentrations for each target: 10 and 10^4^ copies/µL. Samples were sequenced on a MinION flow cell, basecalled, and demultiplexed. All 20 targets were detectable by nanopore sequencing at initial template concentrations of 10 and 10^4^ copies/µL (**Figure S8, S9, Table S6**). In both reaction conditions, less reads were observed for three assay targets: stx1, STh, aatA. Though additional optimization (e.g., adjusting primer concentrations) could be performed to improve relative amplification of these three targets, many factors in environmental samples that cannot be controlled likely play a larger role in determining differential amplification. For example, the absolute and relative abundance of each target in real samples cannot be optimized. For design simplicity, we opted to continue with equimolar SAMRS-AEGIS primer concentrations.

As designed, this SAMRS-AEGIS 20-plex PCR reaction overcomes challenges that must be addressed for sensitive detection of multiple targets in environmental samples. Pathogen and antimicrobial resistance genes can be in low abundance,^52–54^ necessitating modifications that avoid primer dimerization. Inclusion of 1-3 SAMRS nucleotides in the seed region of the 20-plex PCR was effective at eliminating detectable primer dimer formation as seen by both gel electrophoresis and qPCR-based melting curve analysis. Target species in environmental samples are often differentially abundant and many times orders of magnitude different.^54^ We performed two PCR reactions sequentially - Reaction 1 uses 40 cycles to detect low abundance species or amplicons with low amplification efficiency, while Reaction 2 uses AEGIS nucleotides to introduce 24-nt sample barcodes for nanopore sequencing in order to minimize background, non-specific amplification. With equimolar amounts of all SAMRS-AEGIS primers, this workflow was sensitive enough to detect all 20 targets using synthetic templates at 10 copies/µL for each target by nanopore sequencing.

### 20-plex assay performance in environmental samples

Previously, we hypothesized that AEGIS nucleotides in the primers could help avoid non-specific ‘background’ amplification in environmental samples. To test this hypothesis, we compared the sequencing outputs of the 20-plex assay using SAMRS-AEGIS primers to standard DNA primers, in three sample types: wastewater, soil, and human feces (**Figure S10-S14, Table S7-9**). Nanopore sequencing reads were demultiplexed and binned into one of four categories: (1) fully map to target; (2) partially map to target; (3) map to primer regions, but not target; (4) unmapped. For all sample types, the SAMRS-AEGIS 20-plex assay had significantly more reads align to targets and fewer reads mapping to only primer regions compared to standard DNA 20-plex assay (Figure 3). Though variations between sample matrices were readily observable, the SAMRS-AEGIS 20-plex assay had between 1.8 – 7.5 times more read alignments to the full-length targets compared to reads derived from the standard DNA 20-plex assay. Conversely, the standard DNA 20-plex assay resulted in an average of 2.4 – 4 times more reads aligning only to primers, but not target, compared to reads from the SAMRS-AEGIS 20-plex assay.

**Figure 3.**
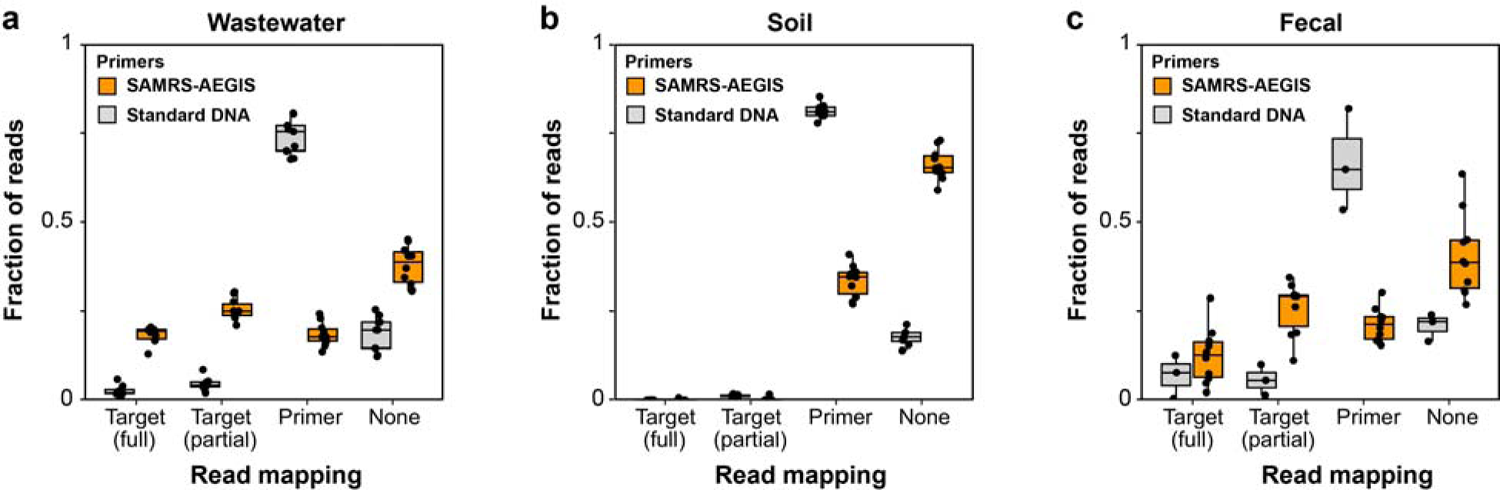
Outcome of nanopore reads in SAMRS-AEGIS and standard DNA 20-plex assays in wastewater, soil, and fecal matrices. Read mapping fractions for each sample, separated by sample matrix type: (**a**) wastewater (*n* = 10 SAMRS-AEGIS; *n* = 9 standard DNA), (**b**) soil (*n* = 10 SAMRS-AEGIS; *n* = 8 standard DNA), and (**c**) fecal (*n* = 10 SAMRS-AEGIS, *n* = 3 standard DNA). For each sample, reads are binned into one of four categories: “Target (full)” = aligns to full target sequence with at least 95% coverage; “Target (partial)” partially aligns to target sequence with < 95% coverage; “Primer” maps to primer regions (priming site, barcode) but not full target; “None” none of the prior bins. Fractions of reads within each sample that fall into each bin are plotted (points) with boxplot overlayed to show the distribution of fractions observed across each sample type-assay combination. Reads mapping to G18S, LT, and ipaH were excluded from this analysis as they were detected in the NTC of the standard DNA 20-plex assay. Box shows interquartile range (25^th^ to 75^th^ percentiles) with the median and whiskers extending to 1.5 times the interquartile range.

The observed increase in on-target alignment of the SAMRS-AEGIS 20-plex assay highlights the importance of non-standard nucleotides as an indispensable component of this 20-plex assay. Reads aligning to only primers constituted the majority of reads from the standard DNA 20-plex assay. Reads in this category constitute a mixture of off-target products, including non-specific amplicons of environmental DNA and primer dimers. Two purification steps involved in preparing the nanopore sequencing library involve steps that partially remove primer dimers that would have been present. As such, a lower fraction of read in the ‘map to primer only’ category could be traced to primer dimers. For sequencing-based detection, the sensitivity of this assay is dependent on sequencing depth. Minimizing wasted sequencing effort on off-target amplicons is critical for minimizing assay costs since it allows users to multiplex more samples per sequencing flow cell.

### Comparison between 20-plex assay and parallelized detection with TaqMan^TM^ Array Cards

To evaluate the performance of the SAMRS-AEGIS 20-plex assay against an established method (Figure 4a), we compared assay results obtained from the 20-plex assay to those from TaqMan^TM^ Array Cards (TAC). TAC assays are qPCR-based and use a highly parallelized architecture to detect multiple targets. Due to their convenience, sensitivity, and potential for semi-quantitative detection, TAC assays are widely used in diagnostic and environmental surveillance settings.^6,51^ Unlike the SAMRS-AEGIS 20-plex, TAC assays also require a fluorescent probe for target identification. Both assays allow for sample multiplexing, with TAC assays limited to eight samples per card. For these comparisons, 10 samples from the SAMRS-AEGIS 20-plex reactions were multiplexed in a single MinION nanopore sequencing flow cell.

**Figure 4.**
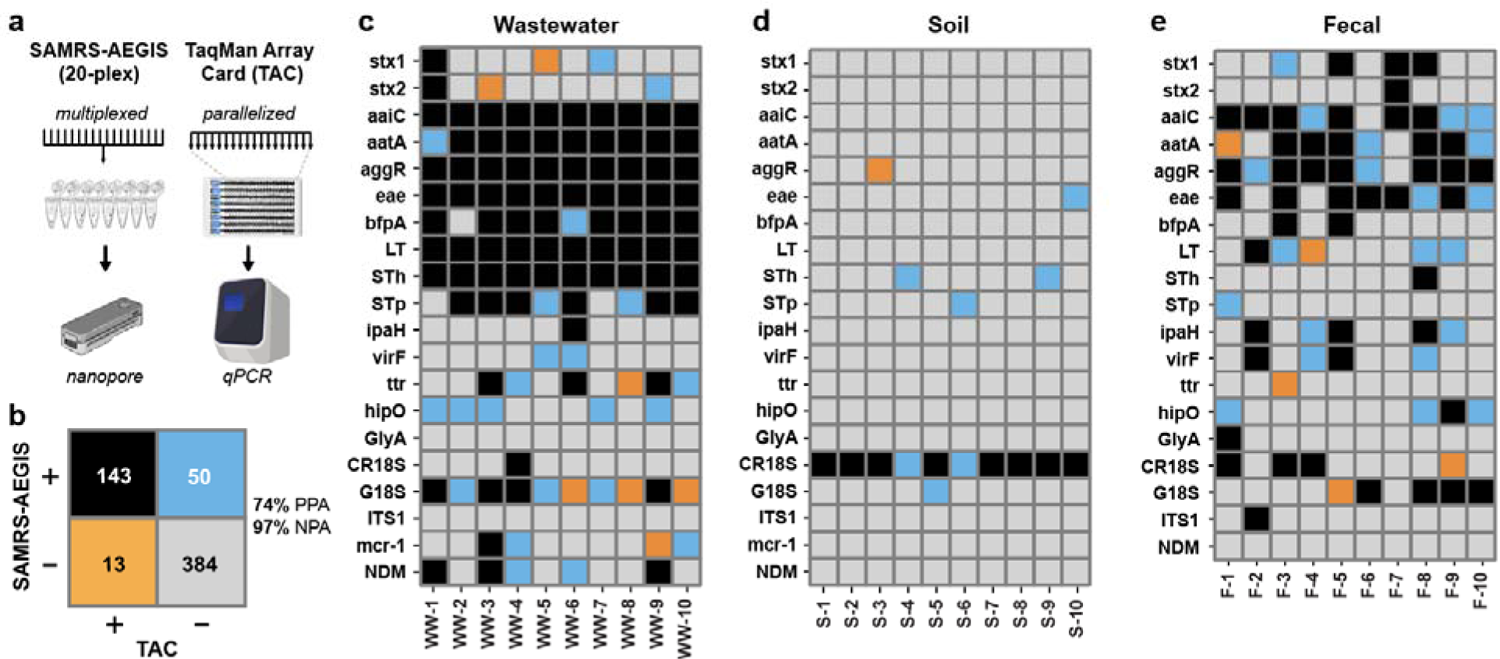
Comparison between SAMRS-AEGIS 20-plex assay and TaqMan^TM^ array cards (TAC) in wastewater, soil, and fecal matrices. (**a**) The SAMRS-AEGIS 20-plex assay uses a highly multiplexed architecture for multi-target detection, while TAC uses a highly parallelized architecture. TAC and SAMRS-AEGIS 20-plex assays were run on 30 environmental samples (10 wastewater, 10 soil, 10 fecal). (**b**) Total assays positive (+) and negative (-) for TAC and SAMRS-AEGIS 20-plex. Percent positive agreement (PPA) and percent negative agreement (NPA) shown. Individual assays positive and negative for TAC and SAMRS-AEGIS 20-plex in (**c**) wastewater, (**d**) soil, and (**e**) fecal samples. Square color indicates assay result, following the color coding used in the total assay result matrix. The TAC *mcr-1* assay was not available for fecal samples.

Comparing target detection between TAC and the SAMRS-AEGIS 20-plex assay, we observed a 74% PPA (positive percent agreement) and a 97% NPA (negative percent agreement) between these two methods (Figure 4b). Among the discrepancies, 13 out of 63 cases involved targets detected by TAC but not by the SAMRS-AEGIS 20-plex assay, while 50 out of 63 cases involved targets detected by the SAMRS-AEGIS 20-plex assay but not by TAC (Figure 4b-e). To confirm the read-to-target assignments in the SAMRS-AEGIS 20-plex assay was not due to mapping error or improper demultiplexing, reads were aligned against reference sequences and manually inspected for processing errors. All 50 targets identified in the SAMRS-AEGIS 20-plex assay, but not in the TAC assay, could be fully mapped to properly barcoded reads. Additionally, no reads in the no-template controls (NTCs) for the SAMRS-AEGIS 20-plex assay could be mapped to assay targets.

These results provide evidence that the SAMRS-AEGIS 20-plex has sensitivity in the tested environmental sample matrices similar to TAC; for certain targets, the SAMRS-AEGIS 20-plex may be more sensitive. One possible reason for the observed differences by method is the assayed template input: TAC uses a maximum of 21 ng template per singleplexed well, while the 20-plex assay uses a maximum of 100 ng template in the first round of PCR. Another possible explanation stems from differences in cycling conditions: TAC uses 40 qPCR cycles, while the SAMRS-AEGIS 20-plex assay uses 40 cycles for first round amplification and 15 for second round of tagged amplification (net: 45 cycles accounting for dilution).

### SAMRS-AEGIS 20-plex assays reveal additional information about microbial threats

We next asked whether additional information is provided by the sequencing data obtained from the SAMRS-AEGIS 20-plex assay. For each sample and each positive gene target with at least 10 mapped reads, consensus sequences were generated and dereplicated to explore diversity across samples. STh, also known as ST1b, encodes a heat stable enterotoxin produced by enterotoxigenic *E. coli* (ETEC).^55^ Of the 12 samples that were positive by STh, three unique STh variants could be identified (Figure 5a). These variants closely match unnamed STh variants present in databases (**Supplementary File 1**).

**Figure 5.**
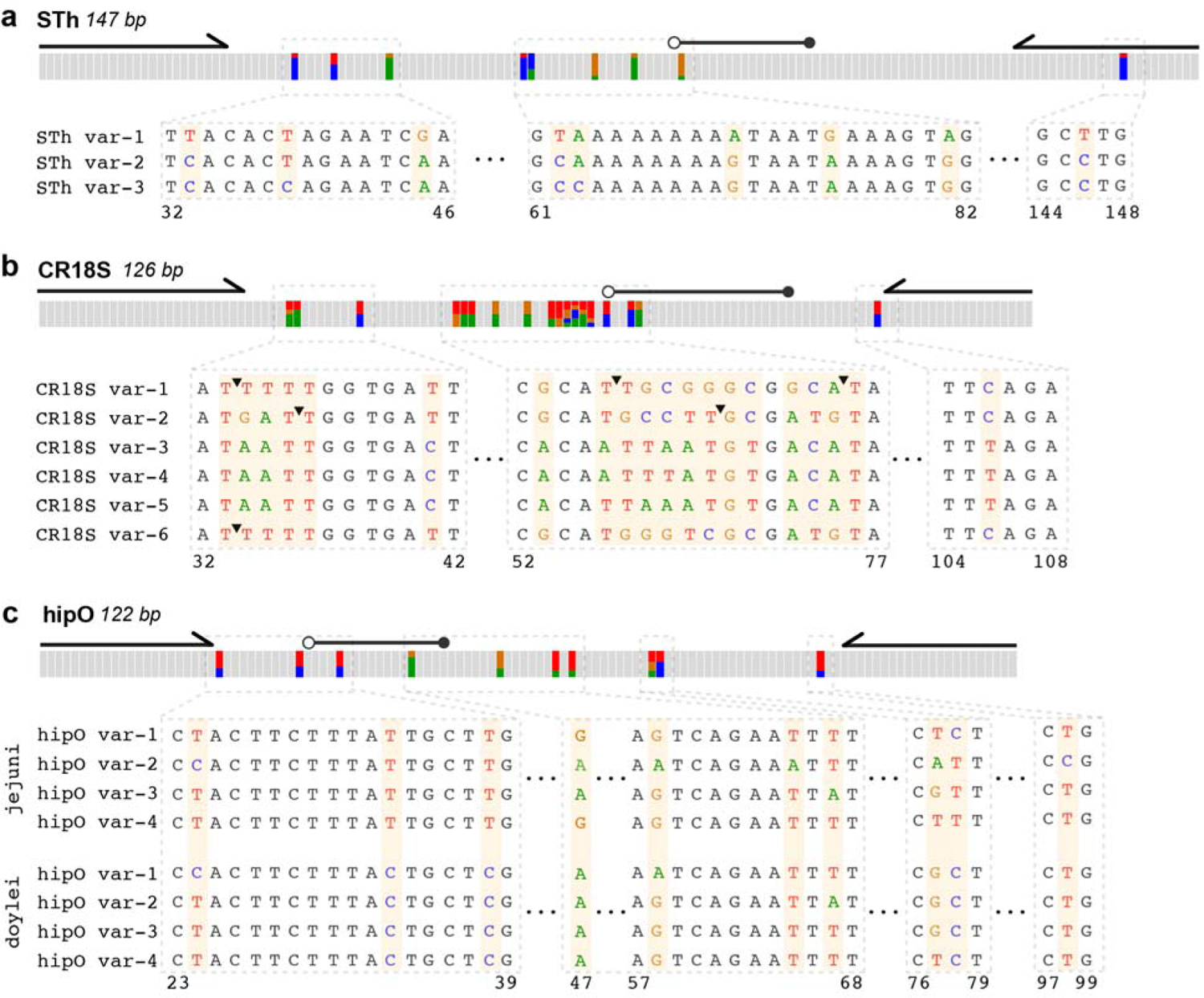
Unique consensus sequences of assay targets identified with the SAMRS-AEGIS 20-plex assay. Unique consensus sequences for targets are aligned against their respective reference sequence. Alignments are shown for (**a**) STh, an enterotoxin gene in *E. coli*; (b) CR18S, 18S rRNA gene of *Cryptosporidium*; and (c) *hipO*, a pathogen gene in *C. jejuni*. Primers and probe locations of the corresponding qPCR assay are marked above each target. Regions of interest are expanded to show single-base resolution. The top-alignment bar shows positions with high variation from consensus alignments in color, color coded by base, and gray otherwise. T - indicates the presence of an insertion.

For antimicrobial resistance genes, *mcr-1* and *bla*_NDM_, we sought to identify different alleles that could be amplified using the SAMRS-AEGIS 20-plex primer set by mapping reads from positive samples to all alleles in the Comprehensive Antibiotic Resistance Database (CARD).^56^ While reads mapped to more than one allele sequence, the putative alleles are highly similar (e.g., *mcr-1.20* and *mcr-1.14* have 1 bp different in the amplicon region) and could not be distinguished from nanopore sequencing error. One strategy to distinguish highly similar alleles within the same sample with nanopore sequencing is incorporating unique molecular identifiers (UMIs) in the primers.^57^ Alternatively, higher accuracy sequencing platforms such as Illumina or PacBio could be used.

Beyond toxins and ARGs, the SAMRS-AEGIS 20-plex also targeted the 18S rRNA gene to identify protozoan pathogens. The CR18S assay targets the 18S rRNA gene in *Cryptosporidium* spp.^58^ 12 of the 14 CR18S SAMRS-AEGIS 20-plex assays that were positive for CR18S were also positive by TAC. Consensus sequences revealed six unique 18S alleles from these samples. Three of these alleles mapped at 100% identity to previously observed variants found in sequence databases, including an unnamed *Cryptosporidium sp.* isolate, a *Cryptosporidium meleagridis* isolate, and a *Cryptosporidium hominis* isolate (Figure 5b). The remaining variants were found to map with lower homology (approximately 90% ID) to uncultured alveolates.

Finally, we were able to observe gene variants belonging to two subspecies of *Campylobacter jejuni.* The *hipO* gene encodes for hippurate hydrolase in *C. jejuni.*^59^ From sequencing we were able to observe eight unique variants (Figure 5c). Four of these variants, with nucleotides T33/T37 in the amplicon, closely map to *C. jejuni* subspecies *jejuni* while the other four, with nucleotides C33/C37, closely map to *C. jejuni* subsp. *doylei*. Two of the *C. jejuni* subsp. *jejuni hipO* variants and two of the *C. jejuni* subsp. *doylei hipO* variant matched with 100% ID to previously observed *hipO* genes. Two of the conserved polymorphisms in the *hipO doylei* variant overlap with the probe binding region for the TAC assay (probe: T48/G47; *doylei* C38/A47). Only one of eight samples positive for *hipO* with the SAMRS-AEGIS 20-plex was positive using TAC.

Sequencing results from the SAMRS-AEGIS 20-plex provided additional insight regarding microbial threats that would have otherwise been missed through a presence/absence-based approach. *C. jejuni* is an important pathogen in LMICs.^60^ A 2016 study of eight birth cohorts across South America, sub-Saharan Africa, and Asia found that 85% of children are carriers of *Campylobacter* spp. before the age of one.^60^ *C jejuni* is also an important food-borne pathogen in HICs primarily transmitted via poultry products.^61^ More samples were positive for *C. jejuni* (*hipO)* than *C. coli* (*GlyA*) in both wastewater samples from WA and child fecal samples from Ecuador. Within *C. jejuni,* two subspecies exist that display differing phenotypic and clinical case presentations. The lesser-known *C. jejuni* subsp*. doylei* is more associated with bacteremia and is known to cause gastritis, in addition to enteritis.^62^ Consensus sequences from our 20-plex assay showed half of the *hipO* amplicons were more similar to *C. jejuni* subsp*. doylei* than *C. jejuni* subsp*. jejuni* (**Supplementary File 1**), though some samples contained a mixture of both species. The ability to distinguish these two subspecies is a notable feature of the 20-plex and provides important information that is useful for both LMIC and HIC settings.

Two assays in this work target eukaryotic 18S rRNA genes: CR18S (*Cryptosporidium spp.*) and G18S (*Giardia spp.*). Of positive G18S samples from the 20-plex assay, all consensus sequences mapped with 100% identity to *Giardia intestinalis*, the causative species of disease in humans.^63^ We observed six variants in *Cryptosporidium* 18S rRNA amplicons, (**Supplementary File 1**), three of which mapped to uncultured alveolates at a lower identity (approx. 90% ID). While the alveolate genus and species is unknown, the positive detection in both SAMRS-AEGIS 20-plex and TAC is possibly the result of off-target amplification of related, but non-pathogenic organisms in the alveolate genera. Given the high variation in the observed amplicons for the CR18S genes, more specific assays that target *C. hominis* and *C. parvum,*^64^ the causative species of illness in humans, may be warranted.^65^ Nonetheless, the sequencing used in the SAMRS-AEGIS 20-plex assay provides a general means to interrogate variants within a sample and distinguish between false positives and true positives.

Though the SAMRS-AEGIS 20-plex assay proved to be sensitive and more information-rich than probe-based amplification strategies, there are some notable limitations. Probe-based strategies that are qPCR-based, such as the TAC assay and dPCR are quantitative. As developed, the 20-plex assay is not capable of relative quantification due to the use of sequential rounds of PCR, nor can the assay be used for absolute quantification since it relies on sequencing. Additionally, sensitivity of assay targets when multiplexing is highly dependent on the differential abundance of target species. Optimization of concentrations for specific targets, combined with *a priori* knowledge of expected environmental abundances, may be required for improving sensitivity in certain sample types. Though the use of nanopore sequencing is well-suited for resource limited settings, the low nominal basecalling accuracy (95%) limits our ability to resolve multiple alleles within the same sample. To distinguish between multiple alleles in a single sample, much higher sequencing coverage with nanopore or other higher accuracy NGS (e.g., Illumina) would be required. Finally, as the 20-plex assay is built around detection of extracted genetic material from samples, it is not suited for detection of viable organisms.

Despite these limitations, the SAMRS-AEGIS 20-plex assay strategy presented is a promising alternative to conventional multi-target PCR detection methods. We estimate that multiplexing 10 samples and 20 targets (20-plex) on a single nanopore MinION flow cell would cost approximately $4.00 per target and $80.00 per sample (**Table S10, Supplementary File 2**), which is similar to the per-target and per-sample costs of 20 parallelized assays on the TAC platform. Where the SAMRS-AEGIS 20-plex assay design excels is at *scale*. Assuming a fixed read coverage, sequencing the 20-plex reactions on an Illumina NovaSeq S4 would drop assay costs to approximately $0.55 per target or $11.00 per sample. Setting aside variable costs, using nanopore sequencing as a readout still offers a lower entry barrier in capital costs compared to qPCR, dPCR, and other NGS platforms, making it well-suited for work in resource constrained settings, such as LMICs.

Lastly, we highlight that the strategy presented in this work is not limited to the 20 targets described here. SAMRS-AEGIS primers are premised on orthogonality, offering an element of modularity for target choices that can be adapted to specific geographic contexts or modified to include emerging threats. Coupled with the additional insight gained from sequencing, this approach has the potential to significantly enhance our understanding of pathogens and antibiotic resistance globally, paving the way for more effective public health interventions.

## Methods

### Sample collection and nucleic acid extraction

10 wastewater samples were obtained from treatment plants from Washington (WA) State. 25-50 mL of wastewater was centrifuged at 5000 x g for 20 minutes at 4 °C. The resulting pellet was resuspended in 200 µL of supernatant. Nucleic acids from 200 µL of resuspended wastewater solids were extracted using AllPrep PowerViral DNA/RNA Kit (Qiagen, Hilden, Germany), omitting the use of β-mercaptoethanol. Purified wastewater DNA was eluted in RNase-free water to a final volume of 100 µL.

10 soil samples were collected from three dog parks located in Seattle, WA. Nucleic acids from 0.25 g of each sample were extracted using DNeasy PowerSoil Pro Kit (Qiagen) following standard protocol. Purified soil DNA was eluted in Solution C6 (10 mM Tris-HCl buffer) to a final volume of 50 µL.

10 fecal samples from children were obtained from the ECoMiD cohort study in northwest Ecuador.^66^ The child stool samples were collected at 18 months of age. Nucleic acids were extracted from 0.22 grams of stool samples using a modified QIAamp Fast DNA Stool Mini Kit (Qiagen). Purified fecal DNA was eluted in Buffer ATE (10 mM Tris-HCl, 0.1 mM EDTA, 0.04% NaN_3_) to a final volume of 200 µL. The ECoMiD study protocol was approved by the institutional review boards of the University of Washington (UW; IRB STUDY00014270), Emory University (IRB00101202), and the Universidad San Francisco de Quito (2018–022M). The study protocol was also reviewed and approved by the Ministry of Health of Ecuador (MSPCURI000253-4).

### SAMRS-AEGIS Primer Design

We selected 40 standard DNA primers from 20 qPCR assays reported in previous literature (**Table S1**). These primers were modified with SAMRS nucleobases to prevent primer dimer in a 20-plex PCR assay. SAMRS modifications were designed using an iterative approach. Software developed at FfAME (PrimerCompare) took all 40 standard DNA primers along with primer, salt, and Mg++ concentrations (200 nM, 60 mM, and 2 mM, respectively) and output potential primer-primer interactions including self-dimerization and hairpin structures. Using filters in the software, we concentrate on only the most detrimental structures with sufficiently low ΔG values for hairpins and dimers, as well as dimers with 3′ to 3′ overlaps within a short footprint (4-8 nt). These become our primary SAMRS substitution regions. We then identified between 1-3 bases for SAMRS substitutions in the 3′-overlap region that can destabilize the largest proportion of predicted structures. PrimerCompare incorporates SAMRS nearest neighbor thermodynamic data and allows us to run the SAMRS modified set as input to evaluate if further substitutions are required, along with checking the Tms and ΔGs of modified primers. This process continues until an optimal set of primers is designed.

Once all 40 standard DNA primers were modified with SAMRS, we added a common AEGIS tag to the 5′-end. The 5′ overhang sequence facilitates the attachment of barcode and sequencing adapters in PCR. The AEGIS tag (AGC**P**CTCG**P**TTC) was designed to allow 2 AEGIS bases separated by 3 or more standard DNA bases and to have a Tm of at least 60 °C. The designed 40 SAMRS-AEGIS primers are listed in **Table S2**. Further, AEGIS barcode sequences used for sample multiplexing were designed by concatenating a barcode from Oxford Nanopore Technologies Native Barcoding Kit (SQK-NBD112.24) with the AEGIS tag sequence, which are listed in **Table S3**.

### SAMRS-AEGIS Primer Synthesis

SAMRS or AEGIS containing oligonucleotides were synthesized on Mermade 12 instruments, using standard phosphoramidite methods with minor changes to the coupling time of AEGIS phosphoramidites (2 min for AEGIS, 1 min for standard DNA bases and SAMRS). Solid support was a Mermade style column packed with controlled pore glass (CPG) at 1000 Å pore size. Oligonucleotides were synthesized as either DMT-on or DMT-off, followed by diethylamine wash (10% in ACN) at the end of the synthesis. DMT-off oligonucleotides were deprotected in aqueous ammonium hydroxide (28-33% NH_3_ in water) at either 65 °C for 3 hours or 55 °C overnight, purified by ion-exchange HPLC (Dionex DNAPac PA-100, 22×250 column), and desalted over SepPak C18 cartridges (Waters Corp., Milford, MA). Oligonucleotides synthesized as DMT-on were deprotected using the same method, followed by purification on Glen-Pak cartridges (GlenResearch, Sterling, VA). The purity of each oligonucleotide was analyzed by analytical ion-exchange HPLC (Dionex DNAPac PA-100, 2×250 column). The oligonucleotides were sent out for ESI mass spectrometry (Novatia LLC, Newtown, PA) to confirm their molecular weights.

### Sequential Multiplex PCR reaction and cycling conditions

Unless otherwise specified, first round PCR was performed at 20 µL scale and contained 1X Quantitect Multiplex PCR NoROX master mix (Qiagen), 0.2 µM final concentration of each primer (40 primers listed in **Table S2, S4** for SAMRS-AEGIS and standard DNA assays, respectively), and nucleic acid template. For reactions that used synthetic templates, 10 - 10^5^ gene copies/µL of synthetic templates (IDT, **Table S6**) were added as specified. Synthetic templates were ordered as IDT gBlock Gene Fragments, except LT, ipaH, G18S, and ITS1, which were ordered as two single-stranded oligos. Oligos for LT, ipaH, G18S, and ITS1 were annealed by adding 20 µM of each oligo in 100 mM of NaCl and 10 mM Tris-EDTA (pH 8.0) buffer and incubating at 90 °C for 3 minutes, then cooling at 0.1 °C/s until reaching 20 °C. For reactions using environmental or fecal DNA extracts, up to 100 ng of DNA extract or 5 µL of volume were added (**Table S7**). First round PCR reactions with SAMRS-AEGIS primers also contained a 0.05 mM final concentration of dZTP (Firebird Biomolecular Sciences, Alachua, Fl). No template control (NTC) reactions were run in parallel with samples, with the template volume replaced by nuclease-free water.

With the exception of experiments where cycling conditions are explicitly varied, first round amplification in sequential PCR was amplified using the following cycling conditions: initial denaturation at 95 °C for 15 min; followed by 40 cycles of (1) 95 °C for 30 s and (2) 60 °C for 60 s; a final extension 72°C for 5 min; then holding step at 12 °C.

1 µL of the PCR product was then used as the template for a second PCR reaction. The second PCR reaction contained template, 1X Quantitect Multiplex PCR NoROX master mix (Qiagen), 2 µM of 24-mer barcoding primer **(Table S3, S5)** in 30 µL of volume, or 20 µL when specified. For reactions that contained SAMRS-AEGIS primers, 0.05 mM of dZTP was added. No template control reactions for each barcoding primer were run in parallel with the samples, with the template volume replaced by nuclease-free water.

The second round PCR reactions were amplified using the following cycling conditions: 95 °C for 15 min, followed by 15 cycles of (1) 95 °C for 30 s and (2) 60 °C for 60 s, followed by 72°C for 5 min and a final holding step at 12 °C. After each round of PCR, amplicons were analyzed by gel electrophoresis on a 3% (w/v) agarose gel stained with GelGreen, and visualized using a blue light transilluminator.

### Nanopore library preparation and data acquisition

Prior to library preparation, all barcoded samples were purified using magnetic DNA-binding beads (Sergi Lab Supplies, Seattle, WA) with a 2:1 bead-to-sample ratio (v/v). Samples were washed twice with 70% ethanol, and eluted in nuclease-free water to a final volume of 12 µL. Purified DNA was quantified on a DeNovix Fluorometer, and barcoded samples were pooled equally by weight. A subset of SAMRS-AEGIS and standard DNA NTCs were also sequenced. Nanopore sample preparation followed standard MinION Genomic DNA by Ligation protocol using the SQK-LSK114 kit with the two following modifications: 1) During the DNA repair and end prep step, the NEBNext FFPE Repair Mix was omitted to avoid potential SAMRS-AEGIS removal by repair enzymes. The volume of the repair mix was replaced by nuclease-free water. 2) To preserve short fragments, the magnetic DNA-binding bead-to-sample ratio was increased to 2:1. Up to 1.3 pmol of pooled samples were loaded into the flowcell. MinION flow cells used in this work were from the R10.4.1 series. Nanopore flow cells were used once per sample without washing, and data collection proceeded for 72 h. A summary of nanopore sequencing runs is shown in **Table S8**.

### Nanopore data collection, basecalling, and processing

Nanopore data acquisition was performed using MinKNOW version 23.07.12. Data was collected in FAST5 format for experiments with synthetic templates, and POD5 format for environmental/fecal samples. FAST5 files were converted to POD5 format using the pod5 package (ONT, version 0.3.2).^67^ Raw POD5 data files were basecalled using Dorado (ONT, version 0.6.2+14a7067) using the super accurate model (dna_r10.4.1_e8.2_400bps_sup@v4.2.0) and a minimum q-score threshold of 7.^68^ Sample barcodes were demultiplexed using the Dorado demux command with the “--no-trim” flag and a custom barcode configuration file that contained the 24-nt barcodes used in this work.

Demultiplexed reads were aligned to a database containing barcoded reference sequences using BLAST Command Line Tool (blastn, NCBI, version 2.9.0+) with the following flags: -- outfmt 10, --max-target-seqs 1.^69^ After alignment, top hits for each read with at least 95% coverage were stored as an initial match. The resulting reads were then passed through a more stringent alignment using bowtie2 (version 2.3.5.1) with the following flags: --very-sensitive, -- local.^70^ Bowtie2 alignment reference sequences contained target sequences without the barcode region for the G18S, eae, CR18S, LT, and ipaH assays, and without the barcode or priming region for the remaining assays. For sub-analysis of *hipO* alleles, both *hipO* variants from *C. jejuni* subsp. *jejuni* and *C. jejuni* subsp *doylei* were included in the reference sequences.

Reads that passed bowtie2 alignment were further aligned to the fully barcoded target sequences. Consensus sequences for each target within each sample were generated from these aligned reads using medaka (ONT, version 1.12.1) commands: ‘consensus’ and ‘stitch’.^71^ For the ‘consensus’ command, no lower limit on the number of sequences required to generate a consensus was placed at consensus generation stage. However, only alignments generated from at least 10 sequences were used for downstream analysis. The ‘stitch’ command used the following flag: –no-fillgaps. Consensus alignment % ID was calculated using the BLAST Command Line (blastn) with the following flags: --outfmt 10, --max-target-seqs 1.

### TaqMan^TM^ Array Card assays

1X TaqMan^TM^ Fast Advanced PCR master mix (Thermo Fisher Scientific) was used for all assays. A maximum amount of either 1400 ng or 20 µL of DNA was loaded into a TaqMan^TM^ Custom Plated Assay Microarray Card **(Table S7**). Six samples were run on each card with a positive and negative control. For the positive control, 1×10^3^ copies/µL of synthetic templates from IDT containing all 20 targets were used (sequences provided in **Table S6**). For the negative control, volume of template DNA was replaced by nuclease-free water. Before running, the loaded card was spun down twice at 300 x g for 1 min. The TaqMan^TM^ Array Card Sealer was then used to seal the card. The QuantStudio 7 Flex System (Thermo Fisher Scientific) was used, with qPCR cycling conditions set at 92 °C for 10 min, followed by 40 cycles of 95 °C for 1 s and 60 °C for 20 s. Data analysis was performed using Design & Analysis Software (version 2.8.0). The fecal samples were run as part of the ECoMiD study using TAC cards with AgPath-ID One-Step RT-PCR master mix and did not have the *mcr-1* assay.

### Comparison of read distributions between the SAMRS-AEGIS 20-plex and standard DNA 20-plex assays

Of the 30 environmental samples collected in this work, 10 wastewater, 10 soil, and 10 fecal samples were processed by the SAMRS-AEGIS 20-plex assay while 9 wastewater, 8 soil, and 3 fecal samples were processed by the standard DNA 20-plex assay. For both assay results, nanopore reads were demultiplexed then binned into one of four categories. Reads were classified as “Target (full)” if they successfully mapped to the intended target following the pipeline outlined in the “Nanopore data collection, basecalling, and processing” section. This pipeline used an initial 95% query mapping filter to remove partial alignments. Reads that mapped to barcodes, primer region, and target amplicon region, but with <95% coverage, were binned as “Target (partial)”. Reads that did not align to the target amplicon region, but did align to barcode and primer regions were binned as “primer”. Reads in this category could include primer dimers and other non-specific amplification products. The remaining reads, which did not fall under the previous categories, were binned as “None”. Reads mapping to G18S, LT, and ipaH were excluded from this analysis since they were detected in the NTC of the standard DNA 20-plex assay. Visuals for read bins were generated using R (version 4.3.2).

### Comparison of target detection between the SAMRS-AEGIS 20-plex assay and the TaqMan^TM^ array cards in environmental samples

10 wastewater, 10 soil, and 10 fecal samples were analyzed for the presence of gene targets by both the SAMRS-AEGIS 20-plex assay and TaqMan^TM^ array cards (TAC). For the SAMRS-AEGIS 20-plex assay, an assay wa considered positive if at least one read successfully mapped to its target according to the pipeline described in the “Nanopore data collection, basecalling, and processing” section; otherwise, it was considered negative. For TAC, an assay was considered positive if at least one of the two replicates in a card reported a Ct value <40; otherwise, it was considered negative. For TAC assays in fecal samples, the *mcr-1* assay was not available and was excluded from analysis.

Agreement and disagreement between SAMRS-AEGIS and TAC for each assay and across all samples were visualized on a plotted matrix. Plots were generated using Python (version 3.8.0). Percent positive agreement (PPA) and percent negative agreement (NPA) was calculated using the following formula:

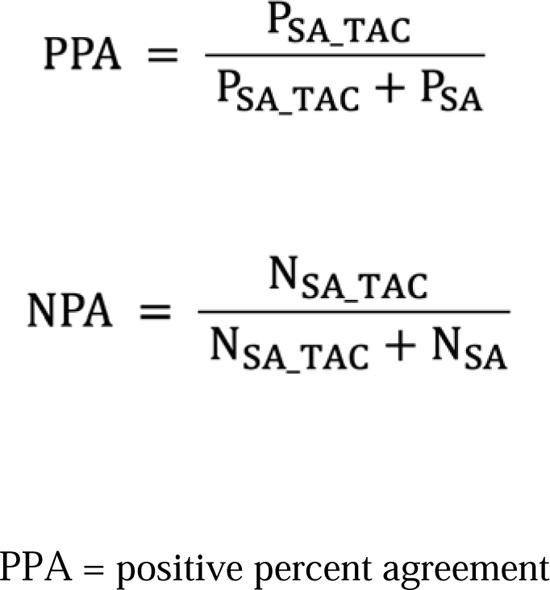

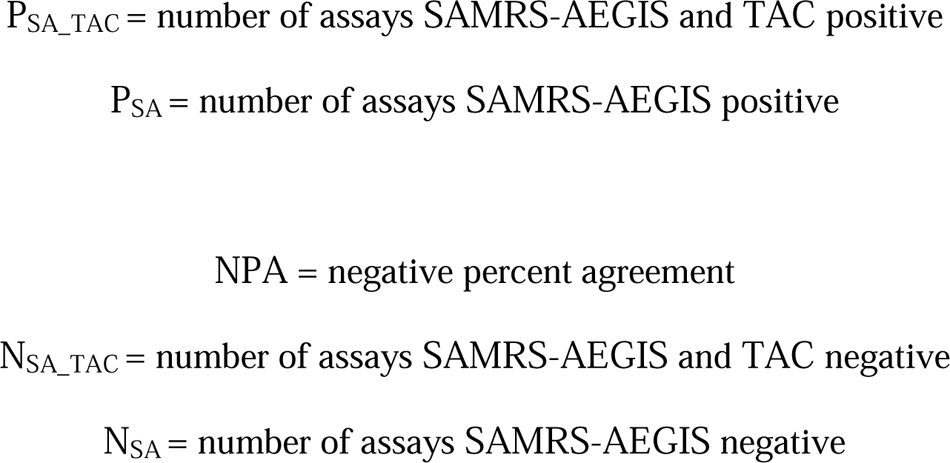

### Identification and visualization of pathogen and antimicrobial resistance gene alleles

Reads were processed as described previously with the inclusion of *hipO* variant sub-analysis specifications. Consensus sequences generated in each sample with a coverage ≥10 reads were aligned to reference sequences of target gene, then dereplicated. Alignments were visualized using Integrative Genomics Viewer (version 2.16.2).^72^ Regions of interest were manually extracted and expanded for visualization. To identify if putative allele sequences had previously been observed, BLASTN webserver was used to map consensus sequences against NCBI core non-redundant nucleic acid database (core_nt).

## Data Availability

Assay results summary (number of mapped reads and TAC Ct values) are provided in a Supplementary data file upon publication in a peer reviewed journal. The demultiplexed nanopore sequencing basecalls (FASTQ) for each sample analyzed in this work have been deposited in the sequence reads archive (SRA) under Bioproject PRJNA1150247.

The demultiplexed nanopore sequencing basecalls (FASTQ) for each sample analyzed in this work have been deposited in the sequence reads archive (SRA) under Bioproject PRJNA1150247 (**Table S9**).

## Acknowledgments

We thank the wastewater treatment plants for collecting samples for this work.

## Author Contribution

Project conceptualization was performed by ERF, JAM, and ZY. Methodology for this work was developed by HK, SMP, LM, and ZY. SAMRS-AEGIS oligonucleotides were synthesized by CC and CM. Fecal samples were contributed by KL and NAZ. Laboratory experiments were performed by HK and NAZ. Data analysis was conducted by HK, SMP, KB, ZY, JAM, and ERF. Visualization of data and results was performed by HK, JAM, and ERF. This project was supervised by ERF. Writing of original draft was carried out by HK, JAM, ERF, and ZY. Reviewing and editing of the manuscript was performed by all.

## References

(1) Baker, R. E.; Mahmud, A. S.; Miller, I. F.; Rajeev, M.; Rasambainarivo, F.; Rice, B. L.; Takahashi, S.; Tatem, A. J.; Wagner, C. E.; Wang, L. F.; Wesolowski, A.; Metcalf, C. J. E. Infectious Disease in an Era of Global Change. Nat. Rev. Microbiol. Nature Research April 1, 2022, pp 193–205. 10.1038/s41579-021-00639-z.

(2) Murray, C. J. L. Global Burden of 288 Causes of Death and Life Expectancy Decomposition in 204 Countries and Territories and 811 Subnational Locations, 1990–2021: A Systematic Analysis for the Global Burden of Disease Study 2021. The Lancet 2024, 403 (10440), 2100–2132. 10.1016/S0140-6736(24)00367-2.

(3) Knee, J.; Sumner, T.; Adriano, Z.; Anderson, C.; Bush, F.; Capone, D.; Casmo, V.; Holcomb, D.; Kolsky, P.; Macdougall, A.; Molotkova, E.; Braga, J. M.; Russo, C.; Schmidt, W. P.; Stewart, J.; Zambrana, W.; Zuin, V.; Nalá, R.; Cumming, O.; Brown, J. Effects of an Urban Sanitation Intervention on Childhood Enteric Infection and Diarrhea in Maputo, Mozambique: A Controlled before-and-after Trial. Elife 2021, 10, e62278. 10.7554/ELIFE.62278.

(4) Grembi, J. A.; Lin, A.; Karim, M. A.; Islam, M. O.; Miah, R.; Arnold, B. F.; Rogawski McQuade, E. T.; Ali, S.; Rahman, M. Z.; Hussain, Z.; Shoab, A. K.; Famida, S. L.; Hossen, M. S.; Mutsuddi, P.; Rahman, M.; Unicomb, L.; Haque, R.; Taniuchi, M.; Liu, J.; Platts-Mills, J. A.; Holmes, S. P.; Stewart, C. P.; Benjamin-Chung, J.; Colford, J. M.; Houpt, E. R.; Luby, S. P. Effect of Water, Sanitation, Handwashing, and Nutrition Interventions on Enteropathogens in Children 14 Months Old: A Cluster-Randomized Controlled Trial in Rural Bangladesh. J. Infect. Dis. 2023, 227 (3), 434–447. 10.1093/infdis/jiaa549.

(5) Platts-Mills, J. A.; Babji, S.; Bodhidatta, L.; Gratz, J.; Haque, R.; Havt, A.; McCormick, B. J. J.; McGrath, M.; Olortegui, M. P.; Samie, A.; Shakoor, S.; Mondal, D.; Lima, I. F. N.; Hariraju, D.; Rayamajhi, B. B.; Qureshi, S.; Kabir, F.; Yori, P. P.; Mufamadi, B.; Amour, C.; Carreon, J. D.; Richard, S. A.; Lang, D.; Bessong, P.; Mduma, E.; Ahmed, T.; Lima, A. A. A. M.; Mason, C. J.; Zaidi, A. K. M.; Bhutta, Z. A.; Kosek, M.; Guerrant, R. L.; Gottlieb, M.; Miller, M.; Kang, G.; Houpt, E. R.; Chavez, C. B.; Trigoso, D. R.; Flores, J. T.; Vasquez, A. O.; Pinedo, S. R.; Acosta, A. M.; Ahmed, I.; Alam, D.; Ali, A.; Rasheed, M.; Soofi, S.; Turab, A.; Yousafzai, A. K.; Bose, A.; Jennifer, M. S.; John, S.; Kaki, S.; Koshy, B.; Muliyil, J.; Raghava, M. V.; Ramachandran, A.; Rose, A.; Sharma, S. L.; Thomas, R. J.; Pan, W.; Ambikapathi, R.; Charu, V.; Dabo, L.; Doan, V.; Graham, J.; Hoest, C.; Knobler, S.; Mohale, A.; Nayyar, G.; Psaki, S.; Rasmussen, Z.; Seidman, J. C.; Wang, V.; Blank, R.; Tountas, K. H.; Swema, B. M.; Yarrot, L.; Nshama, R.; Ahmed, A. M. S.; Tofail, F.; Hossain, I.; Islam, M.; Mahfuz, M.; Chandyo, R. K.; Shrestha, P. S.; Shrestha, R.; Ulak, M.; Black, R.; Caulfield, L.; Checkley, W.; Chen, P.; Lee, G.; Murray-Kolb, L. E.; Schaefer, B.; Pendergast, L.; Abreu, C.; Costa, H.; Di Moura, A.; Filho, J. Q.; Leite, Á.; Lima, N.; Maciel, B.; Moraes, M.; Mota, F.; Oriá, R.; Quetz, J.; Soares, A.; Patil, C. L.; Mahopo, C.; Mapula, A.; Nesamvuni, C.; Nyathi, E.; Barrett, L.; Petri, W. A.; Scharf, R.; Shrestha, B.; Shrestha, S. K.; Strand, T.; Svensen, E. Pathogen-Specific Burdens of Community Diarrhoea in Developing Countries: A Multisite Birth Cohort Study (MAL-ED). Lancet Glob. Health 2015, 3 (9), e564– e575. 10.1016/S2214-109X(15)00151-5.

(6) Liu, J.; Platts-Mills, J. A.; Juma, J.; Kabir, F.; Nkeze, J.; Okoi, C.; Operario, D. J.; Uddin, J.; Ahmed, S.; Alonso, P. L.; Antonio, M.; Becker, S. M.; Blackwelder, W. C.; Breiman, R. F.; Faruque, A. S. G.; Fields, B.; Gratz, J.; Haque, R.; Hossain, A.; Hossain, M. J.; Jarju, S.; Qamar, F.; Iqbal, N. T.; Kwambana, B.; Mandomando, I.; McMurry, T. L.; Ochieng, C.; Ochieng, J. B.; Ochieng, M.; Onyango, C.; Panchalingam, S.; Kalam, A.; Aziz, F.; Qureshi, S.; Ramamurthy, T.; Roberts, J. H.; Saha, D.; Sow, S. O.; Stroup, S. E.; Sur, D.; Tamboura, B.; Taniuchi, M.; Tennant, S. M.; Toema, D.; Wu, Y.; Zaidi, A.; Nataro, J. P.; Kotloff, K. L.; Levine, M. M.; Houpt, E. R. Use of Quantitative Molecular Diagnostic Methods to Identify Causes of Diarrhoea in Children: A Reanalysis of the GEMS Case-Control Study. The Lancet 2016, 388 (10051), 1291–1301. 10.1016/S0140-6736(16)31529-X.

(7) Tokars, J. I.; Olsen, S. J.; Reed, C. Seasonal Incidence of Symptomatic Influenza in the United States. Clin. Infect. Dis. 2018, 66 (10), 1511–1518. 10.1093/cid/cix1060.

(8) Khabbaz, R. F.; Moseley, R. R.; Steiner, R. J.; Levitt, A. M.; Bell, B. P. Challenges of Infectious Diseases in the USA. The Lancet. Elsevier B.V. 2014, pp 53–63. 10.1016/S0140-6736(14)60890-4.

(9) Kilaru, P.; Hill, D.; Anderson, K.; Collins, M. B.; Green, H.; Kmush, B. L.; Larsen, D. A. Wastewater Surveillance for Infectious Disease: A Systematic Review. Am. J. Epidemiol. Oxford University Press February 1, 2023, pp 305–322. 10.1093/aje/kwac175.

(10) Asghar, H.; Diop, O. M.; Weldegebriel, G.; Malik, F.; Shetty, S.; El Bassioni, L.; Akande, A. O.; Al Maamoun, E.; Zaidi, S.; Adeniji, A. J.; Burns, C. C.; Deshpande, J.; Oberste, M. S.; Lowther, S. A. Environmental Surveillance for Polioviruses in the Global Polio Eradication Initiative. J. Infect. Dis. 2014, 210 (suppl 1), S294–S303. 10.1093/infdis/jiu384.

(11) Kitakawa, K.; Kitamura, K.; Yoshida, H. Monitoring Enteroviruses and SARS-CoV-2 in Wastewater Using the Polio Environmental Surveillance System in Japan. Appl. Environ. Microbiol. 2023, 89 (4), e01853–22. 10.1128/aem.01853-22.

(12) Zhou, N. A.; Fagnant-Sperati, C. S.; Komen, E.; Mwangi, B.; Mukubi, J.; Nyangao, J.; Hassan, J.; Chepkurui, A.; Maina, C.; van Zyl, W. B.; Matsapola, P. N.; Wolfaardt, M.; Ngwana, F. B.; Jeffries-Miles, S.; Coulliette-Salmond, A.; Peñaranda, S.; Shirai, J. H.; Kossik, A. L.; Beck, N. K.; Wilmouth, R.; Boyle, D. S.; Burns, C. C.; Taylor, M. B.; Borus, P.; Meschke, J. S. Feasibility of the Bag-Mediated Filtration System for Environmental Surveillance of Poliovirus in Kenya. Food Environ. Virol. 2020, 12, 35–47. 10.1007/s12560-019-09412-1.

(13) Naughton, C. C.; Roman, F. A.; Alvarado, A. G. F.; Tariqi, A. Q.; Deeming, M. A.; Kadonsky, K. F.; Bibby, K.; Bivins, A.; Medema, G.; Ahmed, W.; Katsivelis, P.; Allan, V.; Sinclair, R.; Rose, J. B. Show Us the Data: Global COVID-19 Wastewater Monitoring Efforts, Equity, and Gaps. FEMS Microbes 2023, 4, xtad003. 10.1093/femsmc/xtad003.

(14) Ahmed, W.; Smith, W. J. M.; Metcalfe, S.; Jackson, G.; Choi, P. M.; Morrison, M.; Field, D.; Gyawali, P.; Bivins, A.; Bibby, K.; Simpson, S. L. Comparison of RT-qPCR and RT-dPCR Platforms for the Trace Detection of SARS-CoV-2 RNA in Wastewater. ACS ES&T Water 2022, 2 (11), 1871–1880. 10.1021/acsestwater.1c00387.

15. Peccia, J.; Zulli, A.; Brackney, D. E.; Grubaugh, N. D.; Kaplan, E. H.; Casanovas-Massana, A.; Ko, A. I.; Malik, A. A.; Wang, D.; Wang, M.; Warren, J. L.; Weinberger, D. M.; Arnold, W.; Omer, S. B. Measurement of SARS-CoV-2 RNA in Wastewater Tracks Community Infection Dynamics. Nat. Biotechnol. 2020, 38 (10), 1164–1167. 10.1038/s41587-020-0684-z.

(16) Kilpatrick, D. R.; Yang, C.-F.; Ching, K.; Vincent, A.; Iber, J.; Campagnoli, R.; Mandelbaum, M.; De, L.; Yang, S.-J.; Nix, A.; Kew, O. M. Rapid Group-, Serotype-, and Vaccine Strain-Specific Identification of Poliovirus Isolates by Real-Time Reverse Transcription-PCR Using Degenerate Primers and Probes Containing Deoxyinosine Residues. J. Clin. Microbiol. 2009, 47 (6), 1939–1941. 10.1128/JCM.00702-09.

(17) Adams, C.; Bias, M.; Welsh, R. M.; Webb, J.; Reese, H.; Delgado, S.; Person, J.; West, R.; Shin, S.; Kirby, A. The National Wastewater Surveillance System (NWSS): From Inception to Widespread Coverage, 2020–2022, United States. Sci. Total Environ. 2024, 924, 171566. 10.1016/j.scitotenv.2024.171566.

(18) Wagner, E. G.; Lanoix, J. N. Excreta Disposal for Rural Areas and Small Communities. World Health Organization; 1958.

(19) Manuel, M.; Amato, H. K.; Pilotte, N.; Chieng, B.; Araka, S. B.; Edoux Eric Siko, J.; Harris, M.; Nadimpalli, M.; Janagaraj, V.; Houngbegnon, P.; Rajendiran, R.; Thamburaj, J.; Puthupalayam Kaliappan, S.; Sirois, A. R.; Walch, G.; Oswald, W. E.; Asbjornsdottir, K. H.; Galagan, S. R.; Walson, J. L.; Williams, S. A.; F Luty, A. J.; Njenga, S. M.; Ibikounlé, M.; Ajjampur, S. S.; Pickering, A. J. Soil Surveillance for Monitoring Soil-Transmitted Helminth Infections: Method Development and Field Testing in Three Countries. medRxiv 2023. 10.1101/2023.09.26.23296174.

(20) Harvey, A. P.; Fuhrmeister, E. R.; Cantrell, M. E.; Pitol, A. K.; Swarthout, J. M.; Powers, J. E.; Nadimpalli, M. L.; Julian, T. R.; Pickering, A. J. Longitudinal Monitoring of SARS-CoV-2 RNA on High-Touch Surfaces in a Community Setting. Environ. Sci. Technol. Lett. 2020, 8 (2), 168–175. 10.1021/acs.estlett.0c00875.

(21) Pitol, A. K.; Julian, T. R. Community Transmission of SARS-CoV-2 by Surfaces: Risks and Risk Reduction Strategies. Environ. Sci. Technol. Lett. 2021, 8 (3), 263–269. 10.1021/acs.estlett.0c00966.

(22) Genné-Bacon, E. A.; Bascom-Slack, C. A. The PARE Project: A Short Course-Based Research Project for National Surveillance of Antibiotic-Resistant Microbes in Environmental Samples. J. Microbiol. Biol. Educ. 2018, 19 (3). 10.1128/jmbe.v19i3.1603.

(23) Liguori, K.; Keenum, I.; Davis, B. C.; Calarco, J.; Milligan, E.; Harwood, V. J.; Pruden, A. Antimicrobial Resistance Monitoring of Water Environments: A Framework for Standardized Methods and Quality Control. Environ. Sci. Technol. 2022, 56 (13), 9149–9160. 10.1021/acs.est.1c08918.

(24) Hart, A.; Warren, J.; Wilkinson, H.; Schmidt, W. Environmental Surveillance of Antimicrobial Resistance (AMR), Perspectives from a National Environmental Regulator in 2023. Euro. Surveill. 2023, 28 (11). 10.2807/1560-7917.ES.2023.28.11.2200367.

(25) Elnifro, E. M.; Ashshi, A. M.; Cooper, R. J.; Klapper, P. E. Multiplex PCR: Optimization and Application in Diagnostic Virology. Clin. Microbiol. Rev. 2000, 13 (4), 559–570.

(26) Qiagen. QIAcuity Application Guide Version 2. In QIAcuity User Manual Extension; 2023.

(27) Kodani, M.; Yang, G.; Conklin, L. M.; Travis, T. C.; Whitney, C. G.; Anderson, L. J.; Schrag, S. J.; Taylor, T. H.; Beall, B. W.; Breiman, R. F.; Feikin, D. R.; Njenga, M. K.; Mayer, L. W.; Oberste, M. S.; Tondella, M. L. C.; Winchell, J. M.; Lindstrom, S. L.; Erdman, D. D.; Fields, B. S. Application of TaqMan Low-Density Arrays for Simultaneous Detection of Multiple Respiratory Pathogens. J. Clin. Microbiol. 2011, 49 (6), 2175–2182. 10.1128/JCM.02270-10.

(28) Applied Biosystems. TaqManTM Gene Expression Assays— TaqManTM Array Cards; User Guide; 2021.

(29) Hoshika, S.; Chen, F.; Leal, N. A.; Benner, S. A. Self-Avoiding Molecular Recognition Systems (SAMRS). Nucleic acids symp. ser. 2008, 52 (1), 129–130. 10.1093/nass/nrn066.

(30) Hoshika, S.; Chen, F.; Leal, N. A.; Benner, S. A. Artificial Genetic Systems: Self□avoiding DNA in PCR and Multiplexed PCR. Angew. Chem. Int. Ed. Engl. 2010, 49 (32), 5554–5557. 10.1002/anie.201001977.

(31) Yang, Z.; Le, J. T.; Hutter, D.; Bradley, K. M.; Overton, B. R.; McLendon, C.; Benner, S. A. Eliminating Primer Dimers and Improving SNP Detection Using Self-Avoiding Molecular Recognition Systems. Biol. Methods Protoc. 2020, 5 (1), bpa004. 10.1093/biomethods/bpaa004.

(32) Benner, S. A. Understanding Nucleic Acids Using Synthetic Chemistry. Acc. Chem. Res. 2004, 37 (10), 784–797. 10.1021/ar040004z.

(33) Yang, Z.; Hutter, D.; Sheng, P.; Sismour, A. M.; Benner, S. A. Artificially Expanded Genetic Information System: A New Base Pair with an Alternative Hydrogen Bonding Pattern. Nucleic Acids Res. 2006, 34 (21), 6095–6101. 10.1093/nar/gkl633.

(34) Hoshika, S.; Leal, N. A.; Kim, M.-J.; Kim, M.-S.; Karalkar, N. B.; Kim, H.-J.; Bates, A. M.; Watkins, N. E.; SantaLucia, H. A.; Meyer, A. J.; DasGupta, S.; Piccirilli, J. A.; Ellington, A. D.; SantaLucia Jr., J.; Georgiadis, M. M.; Benner, S. A. Hachimoji DNA and RNA: A Genetic System with Eight Building Blocks. Science (1979) 2019, 363 (6429), 884–887. 10.1126/science.aat0971.

(35) Yang, Z.; Chen, F.; Chamberlin, S. G.; Benner, S. A. Expanded Genetic Alphabets in the Polymerase Chain Reaction. Angew. Chem. Int. Ed. 2010, 49 (1), 177–180. 10.1002/anie.200905173.

(36) Glushakova, L. G.; Bradley, A.; Bradley, K. M.; Alto, B. W.; Hoshika, S.; Hutter, D.; Sharma, N.; Yang, Z.; Kim, M. J.; Benner, S. A. High-Throughput Multiplexed XMAP Luminex Array Panel for Detection of Twenty Two Medically Important Mosquito-Borne Arboviruses Based on Innovations in Synthetic Biology. J. Virol. Methods 2015, 214, 60–74. 10.1016/j.jviromet.2015.01.003.

(37) Glushakova, L. G.; Alto, B. W.; Kim, M. S.; Bradley, A.; Yaren, O.; Benner, S. A. Detection of Chikungunya Viral RNA in Mosquito Bodies on Cationic (Q) Paper Based on Innovations in Synthetic Biology. J. Virol. Methods 2017, 246, 104–111. 10.1016/j.jviromet.2017.04.013.

(38) Yaren, O.; Glushakova, L. G.; Bradley, K. M.; Hoshika, S.; Benner, S. A. Standard and AEGIS Nicking Molecular Beacons Detect Amplicons from the Middle East Respiratory Syndrome Coronavirus. J. Virol. Methods 2016, 236, 54–61. 10.1016/j.jviromet.2016.07.008.

(39) Glushakova, L. G.; Sharma, N.; Hoshika, S.; Bradley, A. C.; Bradley, K. M.; Yang, Z.; Benner, S. A. Detecting Respiratory Viral RNA Using Expanded Genetic Alphabets and Self-Avoiding DNA. Anal. Biochem. 2015, 489, 62–72. 10.1016/j.ab.2015.08.015.

(40) Wang, Y.; Jiao, W.-W.; Wang, Y.; Wang, Y.-C.; Shen, C.; Qi, H.; Shen A-Dong. Graphene Oxide and Self-Avoiding Molecular Recognition Systems-Assisted Recombinase Polymerase Amplification Coupled with Lateral Flow Bioassay for Nucleic Acid Detection. Microchim Acta 2020, 187 (667), 1–11. 10.1007/s00604-020-04637-5.

(41) Yaren, O.; Bradley, K. M.; Moussatche, P.; Hoshika, S.; Yang, Z.; Zhu, S.; Karst, S. M.; Benner, S. A. A Norovirus Detection Architecture Based on Isothermal Amplification and Expanded Genetic Systems. J. Virol. Methods 2016, 237, 64–71. 10.1016/j.jviromet.2016.08.012.

(42) Montero, L.; Smith, S. M.; Jesser, K. J.; Paez, M.; Ortega, E.; Peña-Gonzalez, A.; Soto-Girón, M. J.; Hatt, J. K.; Sánchez, X.; Puebla, E.; Endara, P.; Cevallos, W.; Konstantinidis, K. T.; Trueba, G.; Levy, K. Distribution of *Escherichia coli* Pathotypes along an Urban–Rural Gradient in Ecuador. Am. J. Trop. Med. Hyg. 2023, 109 (3), 559–567. 10.4269/ajtmh.23-0167.

(43) Kotloff, K. L.; Nataro, J. P.; Blackwelder, W. C.; Nasrin, D.; Farag, T. H.; Panchalingam, S.; Wu, Y.; Sow, S. O.; Sur, D.; Breiman, R. F.; Faruque, A. S. G.; Zaidi, A. K. M.; Saha, D.; Alonso, P. L.; Tamboura, B.; Sanogo, D.; Onwuchekwa, U.; Manna, B.; Ramamurthy, T.; Kanungo, S.; Ochieng, J. B.; Omore, R.; Oundo, J. O.; Hossain, A.; Das, S. K.; Ahmed, S.; Qureshi, S.; Quadri, F.; Adegbola, R. A.; Antonio, M.; Hossain, M. J.; Akinsola, A.; Mandomando, I.; Nhampossa, T.; Acácio, S.; Biswas, K.; O’Reilly, C. E.; Mintz, E. D.; Berkeley, L. Y.; Muhsen, K.; Sommerfelt, H.; Robins-Browne, R. M.; Levine, M. M. Burden and Aetiology of Diarrhoeal Disease in Infants and Young Children in Developing Countries (the Global Enteric Multicenter Study, GEMS): A Prospective, Case-Control Study. The Lancet 2013, 382 (9888), 209–222. 10.1016/S0140-6736(13)60844-2.

(44) Pullan, R. L.; Smith, J. L.; Jasrasaria, R.; Brooker, S. J. Global Numbers of Infection and Disease Burden of Soil Transmitted Helminth Infections in 2010. Parasit. Vectors 2014, 7 (37), 1–19. 10.1186/1756-3305-7-37.

(45) Holland, C.; Sepidarkish, M.; Deslyper, G.; Abdollahi, A.; Valizadeh, S.; Mollalo, A.; Mahjour, S.; Ghodsian, S.; Ardekani, A.; Behniafar, H.; Gasser, R. B.; Rostami, A. Global Prevalence of *Ascaris* Infection in Humans (2010–2021): A Systematic Review and Meta-Analysis. Infec. Dis. Poverty 2022, 11 (113). 10.1186/s40249-022-01038-z.

(46) Centers for Disease Control and Prevention. Estimated Annual Number of Episodes of Illnesses Caused by 31 Pathogens Transmitted Commonly by Food, United States; 2019. www.cdc.gov/EID/content/17/1/7-Techapp4.pdf.

(47) Wang, R.; Van Dorp, L.; Shaw, L. P.; Bradley, P.; Wang, Q.; Wang, X.; Jin, L.; Zhang, Q.; Liu, Y.; Rieux, A.; Dorai-Schneiders, T.; Weinert, L. A.; Iqbal, Z.; Didelot, X.; Wang, H.; Balloux, F. The Global Distribution and Spread of the Mobilized Colistin Resistance Gene *mcr-1*. Nat. Commun. 2018, 9 (1), 1179. 10.1038/s41467-018-03205-z.

(48) Khan, A. U.; Maryam, L.; Zarrilli, R. Structure, Genetics and Worldwide Spread of New Delhi Metallo-β-Lactamase (NDM): A Threat to Public Health. BMC Microbiol. BioMed Central Ltd. April 27, 2017. 10.1186/s12866-017-1012-8.

(49) Rao, G.; Capone, D.; Zhu, K.; Knoble, A.; Linden, Y.; Clark, R.; Lai, A.; Kim, J.; Huang, C.-H.; Bivins, A.; Brown, J. Simultaneous Detection and Quantification of Multiple Pathogen Targets in Wastewater. PLOS Water 2024, 3 (2), e0000224. 10.1371/journal.pwat.0000224.

(50) Capone, D.; Berendes, D.; Cumming, O.; Holcomb, D.; Knee, J.; Konstantinidis, K. T.; Levy, K.; Nalá, R.; Risk, B. B.; Stewart, J.; Brown, J. Impact of an Urban Sanitation Intervention on Enteric Pathogen Detection in Soils. Environ. Sci. Technol. 2021, 55 (14), 9989–10000. 10.1021/acs.est.1c02168.

(51) Lappan, R.; Henry, R.; Chown, S. L.; Luby, S. P.; Higginson, E. E.; Bata, L.; Jirapanjawat, T.; Schang, C.; Openshaw, J. J.; O’Toole, J.; Lin, A.; Tela, A.; Turagabeci, A.; Wong, T. H. F.; French, M. A.; Brown, R. R.; Leder, K.; Greening, C.; McCarthy, D. Monitoring of Diverse Enteric Pathogens across Environmental and Host Reservoirs with TaqMan Array Cards and Standard qPCR: A Methodological Comparison Study. *Lancet Planet*. Health 2021, 5 (5), e297–e308. 10.1016/S2542-5196(21)00051-6.

(52) Baker, K. K.; Senesac, R.; Sewell, D.; Sen Gupta, A.; Cumming, O.; Mumma, J. Fecal Fingerprints of Enteric Pathogen Contamination in Public Environments of Kisumu, Kenya, Associated with Human Sanitation Conditions and Domestic Animals. Environ. Sci. Technol. 2018, 52 (18), 10263–10274. 10.1021/acs.est.8b01528.

(53) Fuhrmeister, E. R.; Ercumen, A.; Pickering, A. J.; Jeanis, K. M.; Ahmed, M.; Brown, S.; Arnold, B. F.; Hubbard, A. E.; Alam, M.; Sen, D.; Islam, S.; Kabir, M. H.; Kwong, L. H.; Islam, M.; Unicomb, L.; Rahman, M.; Boehm, A. B.; Luby, S. P.; Colford, J. M.; Nelson, K. L. Predictors of Enteric Pathogens in the Domestic Environment from Human and Animal Sources in Rural Bangladesh. Environ. Sci. Technol. 2019, 53 (17), 10023–10033. 10.1021/acs.est.8b07192.

(54) Gerba, C. P. Environmentally Transmitted Pathogens. In Environmental Microbiology; 2015; pp 509–550. 10.1016/B978-0-12-394626-3.00022-3.

(55) BoDlin, I.; Wiklund, G.; Qadri, F.; Torres, O.; Bourgeois, A. L.; Savarino, S.; Svennerholm, A.-M. Enterotoxigenic *Escherichia coli* with STh and STp Genotypes Is Associated with Diarrhea Both in Children in Areas of Endemicity and in Travelers. J. Clin. Microbiol. 2006, 44 (11), 3872–3877. 10.1128/JCM.00790-06.

(56) Alcock, B. P.; Raphenya, A. R.; Lau, T. T. Y.; Tsang, K. K.; Bouchard, M.; Edalatmand, A.; Huynh, W.; Nguyen, A.-L. V; Cheng, A. A.; Liu, S.; Min, S. Y.; Miroshnichenko, A.; Tran, H.-K.; Werfalli, R. E.; Nasir, J. A.; Oloni, M.; Speicher, D. J.; Florescu, A.; Singh, B.; Faltyn, M.; Hernandez-Koutoucheva, A.; Sharma, A. N.; Bordeleau, E.; Pawlowski, A. C.; Zubyk, H. L.; Dooley, D.; Griffiths, E.; Maguire, F.; Winsor, G. L.; Beiko, R. G.; Brinkman, F. S. L.; Hsiao, W. W. L.; Domselaar, G. V; McArthur, A. G. CARD 2020: Antibiotic Resistome Surveillance with the Comprehensive Antibiotic Resistance Database. Nucleic Acids Res. 2020, 48 (D1), D517–D525. 10.1093/nar/gkz935.

(57) Karst, S. M.; Ziels, R. M.; Kirkegaard, R. H.; Sørensen, E. A.; McDonald, D.; Zhu, Q.; Knight, R.; Albertsen, M. High-Accuracy Long-Read Amplicon Sequences Using Unique Molecular Identifiers with Nanopore or PacBio Sequencing. Nat. Methods 2021, 18 (2), 165–169. 10.1038/s41592-020-01041-y.

(58) Liu, J.; Gratz, J.; Amour, C.; Kibiki, G.; Becker, S.; Janaki, L.; Verweij, J. J.; Taniuchi, M.; Sobuz, S. U.; Haque, R.; Haverstick, D. M.; Houpt, E. R. A Laboratory-Developed TaqMan Array Card for Simultaneous Detection of 19 Enteropathogens. J. Clin. Microbiol. 2013, 51 (2), 472–480. 10.1128/JCM.02658-12.

(59) Liu, J.; Gratz, J.; Amour, C.; Nshama, R.; Walongo, T.; Maro, A.; Mduma, E.; Platts-Mills, J.; Boisen, N.; Nataro, J.; Haverstick, D. M.; Kabir, F.; Lertsethtakarn, P.; Silapong, S.; Jeamwattanalert, P.; Bodhidatta, L.; Mason, C.; Begum, S.; Haque, R.; Praharaj, I.; Kang, G.; Houpt, E. R. Optimization of Quantitative PCR Methods for Enteropathogen Detection. PLoS One 2016, 11 (6), e0158199. 10.1371/journal.pone.0158199.

(60) Amour, C.; Gratz, J.; Mduma, E.; Svensen, E.; Rogawski, E. T.; McGrath, M.; Seidman, J. C.; McCormick, B. J. J.; Shrestha, S.; Samie, A.; Mahfuz, M.; Qureshi, S.; Hotwani, A.; Babji, S.; Trigoso, D. R.; Lima, A. A. M.; Bodhidatta, L.; Bessong, P.; Ahmed, T.; Shakoor, S.; Kang, G.; Kosek, M.; Guerrant, R. L.; Lang, D.; Gottlieb, M.; Houpt, E. R.; Platts-Mills, J. A. Epidemiology and Impact of *Campylobacter* Infection in Children in 8 Low-Resource Settings: Results from the MAL-ED Study. Clin. Infect. Dis. 2016, 63 (9), 1171–1179. 10.1093/cid/ciw542.

(61) Ford, L.; Healy, J. M.; Cui, Z.; Ahart, L.; Medalla, F.; Ray, L. C.; Reynolds, J.; Laughlin, M. E.; Vugia, D. J.; Hanna, S.; Bennett, C.; Chen, J.; Rose, E. B.; Bruce, B. B.; Payne, D. C.; Francois Watkins, L. K. Epidemiology and Antimicrobial Resistance of Campylobacter Infections in the United States, 2005–2018. Open Forum Infect. Dis. 2023, 10 (8), ofad378. 10.1093/ofid/ofad378.

(62) Parker, C. T.; Miller, W. G.; Horn, S. T.; Lastovica, A. J. Common Genomic Features of *Campylobacter jejuni* Subsp. *doylei* Strains Distinguish Them from *C. jejuni* Subsp. *jejuni*. BMC Microbiol. 2007, 7, 50. 10.1186/1471-2180-7-50.

(63) Rojas-López, L.; Marques, R. C.; Svärd, S. G. Giardia Duodenalis. Trends in Parasitol. 2022, 38 (7), 605–606. 10.1016/j.pt.2022.01.001.

(64) Hadfield, S. J.; Robinson, G.; Elwin, K.; Chalmers, R. M. Detection and Differentiation of *Cryptosporidium* Spp. in Human Clinical Samples by Use of Real-Time PCR. J. Clin. Microbiol. 2011, 49 (3), 918–924. 10.1128/JCM.01733-10.

(65) Carey, C. M.; Lee, H.; Trevors, J. T. Biology, Persistence and Detection of Cryptosporidium parvum and Cryptosporidium hominis Oocyst. Water Res. 2004, 38 (4), 818–862. 10.1016/j.watres.2003.10.012.

(66) Lee, G. O.; Eisenberg, J. N. S.; Uruchima, J.; Vasco, G.; Smith, S. M.; Van Engen, A.; Victor, C.; Reynolds, E.; MacKay, R.; Jesser, K. J.; Castro, N.; Calvopiña, M.; Konstantinidis, K. T.; Cevallos, W.; Trueba, G.; Levy, K. Gut Microbiome, Enteric Infections and Child Growth across a Rural– Urban Gradient: Protocol for the ECoMiD Prospective Cohort Study. BMJ Open 2021, 11 (10), e046241. 10.1136/bmjopen-2020-046241.

(67) Oxford Nanopore Technologies. Pod5-File-Format. 2024. https://github.com/nanoporetech/pod5-file-format (accessed 2024-08-16).

(68) Oxford Nanopore Technologies. Dorado. 2024. https://github.com/nanoporetech/dorado (accessed 2024-08-16).

(69) Madden, T. The BLAST Sequence Analysis Tool. In The NCBI handbook 2.*5*; 2013; pp 425–436.

(70) Langmead, B.; Salzberg, S. L. Fast Gapped-Read Alignment with Bowtie 2. Nat. Methods 2012, 9 (4), 357–359. 10.1038/nmeth.1923.

(71) Oxford Nanopore Technologies. Medaka. 2022. https://github.com/nanoporetech/medaka (accessed 2024-08-16).

(72) Robinson, J. T.; Thorvaldsdóttir, H.; Winckler, W.; Guttman, M.; Lander, E. S.; Getz, G.; Mesirov, J. P. Integrative Genomics Viewer. Nat Biotechnol 2011, 29 (1), 24–26. 10.1038/nbt.1754.

