## Supporting Information for "Harnessing non-standard nucleic acids for highly sensitive icosaplex (20-plex) detection of microbial threats"

**Supplementary Methods**

| *Commercial materials* | S3 |
| --- | --- |
| *One-pot singleplex PCR reaction conditions* | S3 |
| *One-pot multiplex PCR reaction conditions* | S4 |
| *qPCR reaction conditions for melt curve generation* | S4 |

**Supplementary Results**

| *Supplementary Table 1. Full sequences of qPCR primers and probes for target genes used in this work* | S5 |
| --- | --- |
| *Supplementary Table 2. SAMRS-AEGIS PCR primer sequences* | S6 |
| *Supplementary Table 3. AEGIS barcode primer sequences used for sample multiplexing* | S7 |
| *Supplementary Table 4. Standard DNA primer sequences for PCR* | S8 |
| *Supplementary Table 5. Standard DNA barcode primer sequences* | S9 |
| *Supplementary Table 6. Full sequences of synthetic templates used in this work* | S10 |
| *Supplementary Table 7. Wastewater, soil, and fecal samples used in this work* | S12 |
| *Supplementary Table 8. Nanopore sequencing run overview* | S13 |
| *Supplementary Table 9. Nanopore datasets uploaded to Sequence Reads Archive (SRA)* | S13 |
| *Supplementary Table 10. Analysis comparing costs between Oxford nanopore sequencing, Illumina sequencing, and the TaqMan^TM^ array card* | S15 |
| *Supplementary Figure 1. Amplification of targets using synthetic templates* | S16 |
| *Supplementary Figure 2. Melt curves of singleplex qPCR using SAMRS-AEGIS primers* | S17 |
| *Supplementary Figure 3. 20-plex PCR using synthetic templates* | S18 |
| *Supplementary Figure 4. One-pot and sequential PCR to minimize amplification bias* | S19 |
| *Supplementary Figure 5. Sequential PCR cycling optimization to minimize amplification bias* | S20 |
| *Supplementary Figure 6. Primer concentration optimization in the first round of sequential PCR* | S21 |
| *Supplementary Figure 7. Comparison of 20-plex amplification in the absence of template using either SAMRS-AEGIS primers or standard DNA primers* | S22 |
| *Supplementary Figure 8. 20-plex PCR of all 20 targets using synthetic templates* | S23 |
| *Supplementary Figure 9. Raw and normalized read count for SAMRS-AEGIS 20-plex target amplification using synthetic templates* | S24 |
| *Supplementary Figure 10. 20-plex PCR of 5 fecal (F-1 to F-5) and 5 wastewater (WW-1 to WW-5) samples using SAMRS-AEGIS primers* | S25 |
| *Supplementary Figure 11. 20-plex PCR of 5 fecal (F-6 to F-10) and 5 wastewater (WW-6 to WW-10) samples using SAMRS-AEGIS primers* | S26 |
| *Supplementary Figure 12. 20-plex PCR of 10 soil samples using SAMRS-AEGIS primers* | S27 |
| *Supplementary Figure 13. 20-plex PCR of 4 wastewater, 3 fecal, and 3 soil samples using standard DNA primers* | S28 |
| *Supplementary Figure 14. 20-plex PCR of 5 soil and 5 wastewater samples using standard DNA primers* | S29 |

| **References** | S30 |
| --- | --- |

**Supplementary Methods**

**Commercial Materials. Synthetic gene blocks (gBlocks) and standard DNA oligonucleotides were purchased from Integrated DNA Technologies (IDT; Coralville, IA).** dP phosphoramidite (dP-PA-102), SAMRS phosphoramidites, dZTP (dZTP-101) were supplied by Firebird Biomolecular Sciences (Alachua, FL). All gene blocks, oligonucleotides, and dZTP were resuspended in Tris-EDTA buffer. dA(Bz) CPG (MM1-1000-1), dC(Ac)-CPG (MM1-1100A-1), dT-CPG (MM1-1300-1) and dG(dmf)-CPG (MM1-1200F-1) were ordered from Biosearch Technologies (Hoddesdon, United Kingdom). SepPak C18 cartridges (WAT020515) were purchased from Waters Corporation (Milford, MA). Glen-pak cartridges (60-5000-96) were purchased from Glen Research (Sterling, VA). QuantiTect Multiplex PCR NoROX (204743), DNeasy PowerSoil Pro (47014), and AllPrep PowerViral DNA/RNA extraction kit (41105519) were purchased from Qiagen (Hilden, Germany). Diethylamine (149450010), Tris-EDTA buffer (J75793-AE), Dionex DNAPac PA-100 22x250 column (088759), Dionex DNAPac PA-100 2x250 column (088760), agarose (0710-500G; electrophoresis grade), GeneRuler 1kb Plus DNA Ladder (SM1331), O’RangeRuler 20 bp DNA ladder (SM1323), TriTrack DNA Loading Dye (R1161), MicroAmp^TM^ Optical Adhesive Film (4311971), and TaqMan^TM^ Custom Plated Assays in MicroArray Cards (4485459) were purchased from Thermo Fisher Scientific (Waltham, MA). 96-well PCR plates, low profile, semi skirted, clear (HSL9601) for qPCR were ordered from Bio-Rad (Hercules, CA). GelGreen 10,000X in DMSO (41004) was purchased from Biotium (Fremont, CA). Ammonium hydroxide (A669-500), 10X Tris-Borate_EDTA solution (BP13334) and Biotium EvaGreen 20X in water (NC0521178) were purchased from Fisher Scientific (Waltham, MA). Sodium chloride (S3014-5KG) was purchased from Sigma-Aldrich (St. Louis, MO). Magnetic DNA-binding beads (1040) were purchased from Sergi Lab Supplies (Seattle, WA). MinION Flow Cells R10.4.1 (FLO-MIN114), MinION sequencing device (MIN-101B), and Ligation Sequencing Kit (SQK-LSK114) were purchased from Oxford Nanopore Technologies (ONT; Oxford, United Kingdom). NEBNext® Companion Module for Oxford Nanopore Technologies® Ligation Sequencing (E7180L) was purchased from New England Biolabs (Ipswitch, MA).

**One-pot singleplex PCR reaction conditions.** SAMRS-AEGIS primer PCR reaction conditions for target amplification (**Figure S1**) included 1X of QuantiTect Multiplex PCR NoROX master mix, 0.4 µM primer **(Table S2**)**,** and 0.05 mM dZTP. Synthetic templates (IDT, **Table S6**) at 0.6 ng for each target (1.2×10^8^ - 4.5×10^8^ copies/µL) was used as the template**.** The PCR was amplified using the following cycling conditions: initial denaturation at 95 °C for 15 min; followed by 30 cycles of (1) 94 °C for 60 s and (2) 60 °C for 60 s; then holding step at 12 °C. Amplicons were run on a 2% (w/v) agarose gel stained with GelGreen, and visualized using a blue light transilluminator.

**One-pot multiplex PCR reaction conditions.** Both standard DNA and SAMRS-AEGIS PCR reaction conditions for a one-pot PCR reaction (**Figure 2c, Figure S3, S4a**) included 1X of QuantiTect Multiplex PCR NoROX master mix, 0.05 or 0.1 µM of primer as specified **(Table S2, S4),** and 2 µM of barcoding primer containing 12-nt barcodes **(Table S3, S5).** The PCR with the SAMRS-AEGIS primers also contained 0.05 mM of dZTP. Synthetic templates (IDT, **Table S6**) at 0.5 ng (3.3 × 10^7^ - 1.3 × 10^8^ copies/µL) or 10 - 1 × 10^5^ copies/µL for each target was used as specified. PCR cycling conditions were as follows: 95 °C for 15 min, then 35 cycles at 95 °C for 30 s and 60 °C for 60 s, followed by 72 °C for 5 min, and a final holding step at 12 °C.  Amplicons were run on a 3% (w/v) agarose gel stained with GelGreen, and visualized using a blue light transilluminator.

**qPCR reaction conditions for melt curve generation.** qPCR reaction conditions included 1X of QuantiTect Multiplex PCR NoROX master mix, 0.2 µM primer **(Table S2, S4),** and 1X EvaGreen dye. SAMRS-AEGIS primer reactions also contained 0.05 mM of dZTP. All 20 synthetic templates (IDT, **Table S6**) at 5 × 10^3^ copies/µL along with individual primer pairs were used in the 20 µL single-plex reaction. qPCR was performed on a Bio-Rad CFX96 instrument and Bio-Rad CFX Maestro 2.2 (version 5.2.008.0222). qPCR cycling conditions were as follows: 95 °C for 15 min, then 40 cycles of 95 °C for 30 s and 60 °C for 60 s, followed by 72°C for 5 min. Melt curve generation was performed as follows: 95 °C for 30 s, 60 °C for 30 s, heating at 0.5 °C/s until reaching 95 °C, 95 °C for 30 s, and a final step at 37 °C for 30 s.  The 20-plex no template control reactions were run on a 3% (w/v) agarose gel stained with GelGreen, and visualized using a blue light transilluminator **(Figure S7**)**.**

**Supplementary Tables**

| **Supplementary Table 1. Full sequences of qPCR primers and probes for target genes used in this work.** Gene target name (Target), amplicon length (Length (bp)), forward primer sequence (Forward primer), reverse primer sequence (Reverse primer), probe sequence (Probe (5′-FAM)), and Reference (Ref) of target genes used in this work. All sequences are shown in the 5′ to 3′ direction. Primer and probe sequences are used in TAC assays. | | | | | |
| --- | --- | --- | --- | --- | --- |
| **Target** | **Length (bp)** | **Forward primer** | **Reverse primer** | **Probe (5′-FAM)** | **Ref** |
| stx1 | 132 | ACTTCTCGACTGCAAAGACGTATG | ACAAATTATCCCCTGWGCCACTATC | CTCTGCAATAGGTACTCCA | ^1^ |
| stx2 | 93 | CCACATCGGTGTCTGTTATTAACC | GGTCAAAACGCGCCTGATAG | TTGCTGTGGATATACGAGG | ^2^ |
| aaiC | 215 | ATTGTCCTCAGGCATTTCAC | ACGACACCCCTGATAAACAA | TAGTGCATACTCATCATTTAAG | ^1^ |
| aatA | 237 | CTGGCGAAAGACTGTATCAT | TTTTGCTTCATAAGCCGATAGA | TGGTTCTCATCTATTACAGACAGC | ^1^ |
| aggR | 95 | GCAATCAGATTAARCAGCGATACA | TTCGGACAACTRCAAGCATC | AAGACGCCTAAAGGATGCCC | ^3,4^ |
| eae | 102 | CATTGATCAGGATTTTTCTGGTGATA | CTCATGCGGAAATAGCCGTTA | ATACTGGCGAGACTATTTCAA | ^1^ |
| bfpA | 110 | TGGTGCTTGCGCTTGCT | CGTTGCGCTCATTACTTCTG | CAGTCTGCGTCTGATTCCAA | ^1^ |
| LT | 62 | TTCCCACCGGATCACCAA | CAACCTTGTGGTGCATGATGA | CTTGGAGAGAAGAACCCT | ^2^ |
| STh | 147 | GCTAAACCAGYAGRGTCTTCAAAA | CCCGGTACARGCAGGATTACAACA | TGGTCCTGAAAGCATGAA | ^1^ |
| STp | 136 | TGAATCACTTGACTCTTCAAAA | GGCAGGATTACAACAAAGTT | TGAACAACACATTTTACTGCT | ^1^ |
| ipaH | 64 | CCTTTTCCGCGTTCCTTGA | CGGAATCCGGAGGTATTGC | CGCCTTTCCGATACCGTCTCTGCA | ^5^ |
| virF | 101 | TCTGAGGAGGAGGTTTCTATCGATT | GAAACAGCTGATAAAAGGCAAGCT | CCGAAAGGCATCTCTTTT | ^6^ |
| ttr | 95 | CTCACCAGGAGATTACAACATGG | AGCTCAGACCAAAAGTGACCATC | CACCGACGGCGAGACCGACTTT | ^4^ |
| hipO | 122 | CTTGCGGTCATGATGGACATAC | AGCACCACCCAAACCCTCTTCA | TGCTTGCTGCAAAGTATT | ^4^ |
| GlyA | 125 | AAACCAAAGCTTATCGTGTGC | AGTGCAGCAATGTGTGCAAT | TAAGCTCCAACTTCATCCG | ^4^ |
| CR18S | 126 | GGGTTGTATTTATTAGATAAAGAACCA | AGGCCAATACCCTACCGTCT | TGACATATCATTCAAGTTTCTGAC | ^1^ |
| G18S | 63 | GACGGCTCAGGACAACGGTT | TTGCCAGCGGTGTCCG | CCCGCGGCGGTCCCTGCTAG | ^7^ |
| ITS1 | 88 | GTAATAGCAGTCGGCGGTTTCTT | GCCCAACATGCCACCTATTC | TTGGCGGACAATTGCATGCGAT | ^8^ |
| mcr-1 | 108 | GATCGCTGTCGTGCTCTTTG | ACCGCGCCCATGATTAATAG | CGATGCTACTGATCACCACG | ^9^ |
| NDM | 109 | ATATCACCGTTGGGATCGAC | TAGTGCTCAGTGTCGGCATC | AAGGACAGCAAGGCCAAGTCG | ^10^ |

**Supplementary Table 2. SAMRS-AEGIS PCR primer sequences.** Gene target name (Target), forward primer sequence (Forward primer), reverse primer sequence (Reverse primer), and their melting temperatures, Tm (°C), of SAMRS-AEGIS PCR primers used in this work. SAMRS bases are noted with an * after the base. AEGIS bases are bolded. All sequences are shown in the 5′ to 3′ direction.

| **Target** | **Forward primer** | **Tm (°C)** | **Reverse primer** | **Tm (°C)** |
| --- | --- | --- | --- | --- |
| stx1 | AGC**P**CTCG**P**TTCACTTCTCGACTGCAAAGACG*TA*TG | 80.09 | AGC**P**CTCG**P**TTCACAAATTATCCCCTGWGCCAC*TA*TC | 80.09 |
| stx2 | AGC**P**CTCG**P**TTCCCACATCGGTGTCTGTTATTAA*C*C | 80.35 | AGC**P**CTCG**P**TTCGGTCAAAACGCGCCTGA*TA*G | 82.02 |
| aaiC | AGC**P**CTCG**P**TTCATTGTCCTCAGGCATTTCA*C | 79.50 | AGC**P**CTCG**P**TTCACGACACCCCTGATAAAC*AA | 80.45 |
| aatA | AGC**P**CTCG**P**TTCCTGGCGAAAGACTGTATC*AT | 81.08 | AGC**P**CTCG**P**TTCTTTTGCTTCATAAGCCGA*TA*GA | 78.15 |
| aggR | AGC**P**CTCG**P**TTCGCAATCAGATTAARCAGCGATA*C*A | 79.97 | AGC**P**CTCG**P**TTCTTCGGACAACTRCAAGC*A*TC | 78.54 |
| eae | AGC**P**CTCG**P**TTCCATTGATCAGGATTTTTCTGGTG*A*TA | 77.93 | AGC**P**CTCG**P**TTCCTCATGCGGAAATAGCC*G*TTA | 81.39 |
| bfpA | AGC**P**CTCG**P**TTCTGGTGCTTGCGCTTG*CT | 83.50 | AGC**P**CTCG**P**TTCCGTTGCGCTCATTACTTC*TG | 82.27 |
| LT | AGC**P**CTCG**P**TTCTTCCCACCGGATCAC*C*AA | 81.42 | AGC**P**CTCG**P**TTCCAACCTTGTGGTGCATGA*TG*A | 80.49 |
| STh | AGC**P**CTCG**P**TTCGCTAAACCAGYAGRGTCTTCAA*A*A | 80.60 | AGC**P**CTCG**P**TTCCCCGGTACARGCAGGATTACAA*C*A | 83.02 |
| STp | AGC**P**CTCG**P**TTCTGAATCACTTGACTCTTCAAA*A | 78.45 | AGC**P**CTCG**P**TTCGGCAGGATTACAACAAAG*TT | 79.86 |
| ipaH | AGC**P**CTCG**P**TTCCCTTTTC*CGCGTTCCTTG*A | 81.92 | AGC**P**CTCG**P**TTCCGGAATCCGGAGGTA*TTG*C | 80.53 |
| virF | AGC**P**CTCG**P**TTCTCTGAGGAGGAGGTTTCTATC*G*ATT | 80.80 | AGC**P**CTCG**P**TTCGAAACAGCTGATAAAAGGCAA*GC*T | 81.34 |
| ttr | AGC**P**CTCG**P**TTCCTCACCAGGAGATTACAACA*TG*G | 78.49 | AGC**P**CTCG**P**TTCAGCTCAGACCAAAAGTGACC*A*TC | 81.17 |
| hipO | AGC**P**CTCG**P**TTCCTTGCGGTCATGATGGACA*TA*C | 80.28 | AGC**P**CTCG**P**TTCAGCACCACCCAAACCCTCTTC*A | 85.49 |
| GlyA | AGC**P**CTCG**P**TTCAAACCAAAGCTTATCGTGTG*C | 79.46 | AGC**P**CTCG**P**TTCAGTGCAGCAATGTGTGC*A*AT | 82.97 |
| CR18S | AGC**P**CTCG**P**TTCGGGTTGTATTTATTAGATAAAGAACC*A | 78.06 | AGC**P**CTCG**P**TTCAGGCCAATACCCTACCG*TC*T | 82.25 |
| G18S | AGC**P**CTCG**P**TTCGACGGCTCAGGACA*AC*GG*TT | 82.49 | AGC**P**CTCG**P**TTCTTGCCAGCGGTG*TCC*G | 82.77 |
| ITS1 | AGC**P**CTCG**P**TTCGTAATAGCAGTCGGCGGTTTC*TT | 83.67 | AGC**P**CTCG**P**TTCGCCCAACATGCCACCTA*TTC | 83.21 |
| mcr-1 | AGC**P**CTCG**P**TTCGATCGCTGTCGTGCTC*TTTG | 83.70 | AGC**P**CTCG**P**TTCACCGCGCCCATGATTAATA*G | 82.42 |
| NDM | AGC**P**CTCG**P**TTCATATCACCGTTGGGATC*GA*C | 79.05 | AGC**P**CTCG**P**TTCTAGTGCTCAGTGTCGGC*A*TC | 81.13 |

**Supplementary Table 3. AEGIS barcode primer sequences used for sample multiplexing.** Barcode number (BC) for barcodes used in sequencing, barcode name (Barcode name) and full sequence (Sequence) of sample barcoding primers used in this work. AEGIS bases are bolded. All sequences are shown in the 5′ to 3′ direction. Barcodes BC11-BC20 contain 12-nt barcode sequences, while NB01-NB10 contain 24-nt barcode sequences.

| **BC** | **Barcode name** | **Sequence** |
| --- | --- | --- |
| - | BC11-5RKB009-Homo-2P | AAGAACCGAGCCAGC**P**CTCG**P**TTC |
| - | BC12-5RKB025-Homo-2P | ACGCTGAAGAATAGC**P**CTCG**P**TTC |
| - | BC13-5RKB029-Homo-2P | AGATTCCGCCGTAGC**P**CTCG**P**TTC |
| - | BC14-5RKB033-Homo-2P | ATCGTCGTGCCTAGC**P**CTCG**P**TTC |
| - | BC15-5RKB035-Homo-2P | ATTCATACCAGCAGC**P**CTCG**P**TTC |
| - | BC16-7RKB036-Homo-2P | CAATACGCTGCAAGC**P**CTCG**P**TTC |
| - | BC17-7RKB056-Homo-2P | CTGAACTCCGCAAGC**P**CTCG**P**TTC |
| - | BC18-7RKB066-Homo-2P | GCTGCAATATACAGC**P**CTCG**P**TTC |
| - | BC19-7RKB074-Homo-2P | TAGTGACGATGGAGC**P**CTCG**P**TTC |
| - | BC20-7RKB77-2Homo-2P | TCCGTATTAGCCAGC**P**CTCG**P**TTC |
| 1 | AMR-NB01-24BC-2P | CACAAAGACACCGACAACTTTCTTAGC**P**CTCG**P**TTC |
| 2 | AMR-NB02-24BC-2P | ACAGACGACTACAAACGGAATCGAAGC**P**CTCG**P**TTC |
| 3 | AMR-NB03-24BC-2P | CCTGGTAACTGGGACACAAGACTCAGC**P**CTCG**P**TTC |
| 4 | AMR-NB04-24BC-2P | TAGGGAAACACGATAGAATCCGAAAGC**P**CTCG**P**TTC |
| 5 | AMR-NB05-24BC-2P | AAGGTTACACAAACCCTGGACAAGAGC**P**CTCG**P**TTC |
| 6 | AMR-NB06-24BC-2P | GACTACTTTCTGCCTTTGCGAGAAAGC**P**CTCG**P**TTC |
| 7 | AMR-NB07-24BC-2P | AAGGATTCATTCCCACGGTAACACAGC**P**CTCG**P**TTC |
| 8 | AMR-NB08-24BC-2P | ACGTAACTTGGTTTGTTCCCTGAAAGC**P**CTCG**P**TTC |
| 9 | AMR-NB09-24BC-2P | AACCAAGACTCGCTGTGCCTAGTTAGC**P**CTCG**P**TTC |
| 10 | AMR-NB10-24BC-2P | GAGAGGACAAAGGTTTCAACGCTTAGC**P**CTCG**P**TTC |

**Supplementary Table 4. Standard DNA primer sequences for PCR.** Gene target name (Target), forward primer sequence (Forward primer), and reverse primer sequence (Reverse primer) of standard DNA PCR primers used in this work. All primers contain a tag for downstream barcoding. All sequences are shown in the 5′ to 3′ direction.

| **Target** | **Forward primer** | **Reverse primer** |
| --- | --- | --- |
| stx1 | AGCGCTCGGTTCACTTCTCGACTGCAAAGACGTATG | AGCGCTCGGTTCACAAATTATCCCCTGWGCCACTATC |
| stx2 | AGCGCTCGGTTCCCACATCGGTGTCTGTTATTAACC | AGCGCTCGGTTCGGTCAAAACGCGCCTGATAG |
| aaiC | AGCGCTCGGTTCATTGTCCTCAGGCATTTCAC | AGCGCTCGGTTCACGACACCCCTGATAAACAA |
| aatA | AGCGCTCGGTTCCTGGCGAAAGACTGTATCAT | AGCGCTCGGTTCTTTTGCTTCATAAGCCGATAGA |
| aggR | AGCGCTCGGTTCGCAATCAGATTAARCAGCGATACA | AGCGCTCGGTTCTTCGGACAACTRCAAGCATC |
| eae | AGCGCTCGGTTCCATTGATCAGGATTTTTCTGGTGATA | AGCGCTCGGTTCCTCATGCGGAAATAGCCGTTA |
| bfpA | AGCGCTCGGTTCTGGTGCTTGCGCTTGCT | AGCGCTCGGTTCCGTTGCGCTCATTACTTCTG |
| LT | AGCGCTCGGTTCTTCCCACCGGATCACCAA | AGCGCTCGGTTCCAACCTTGTGGTGCATGATGA |
| STh | AGCGCTCGGTTCGCTAAACCAGYAGRGTCTTCAAAA | AGCGCTCGGTTCCCCGGTACARGCAGGATTACAACA |
| STp | AGCGCTCGGTTCTGAATCACTTGACTCTTCAAAA | AGCGCTCGGTTCGGCAGGATTACAACAAAGTT |
| ipaH | AGCGCTCGGTTCCCTTTTCCGCGTTCCTTGA | AGCGCTCGGTTCCGGAATCCGGAGGTATTGC |
| virF | AGCGCTCGGTTCTCTGAGGAGGAGGTTTCTATCGATT | AGCGCTCGGTTCGAAACAGCTGATAAAAGGCAAGCT |
| ttr | AGCGCTCGGTTCCTCACCAGGAGATTACAACATGG | AGCGCTCGGTTCAGCTCAGACCAAAAGTGACCATC |
| hipO | AGCGCTCGGTTCCTTGCGGTCATGATGGACATAC | AGCGCTCGGTTCAGCACCACCCAAACCCTCTTCA |
| GlyA | AGCGCTCGGTTCAAACCAAAGCTTATCGTGTGC | AGCGCTCGGTTCAGTGCAGCAATGTGTGCAAT |
| CR18S | AGCGCTCGGTTCGGGTTGTATTTATTAGATAAAGAACCA | AGCGCTCGGTTCAGGCCAATACCCTACCGTCT |
| G18S | AGCGCTCGGTTCGACGGCTCAGGACAACGGTT | AGCGCTCGGTTCTTGCCAGCGGTGTCCG |
| ITS1 | AGCGCTCGGTTCGTAATAGCAGTCGGCGGTTTCTT | AGCGCTCGGTTCGCCCAACATGCCACCTATTC |
| mcr-1 | AGCGCTCGGTTCGATCGCTGTCGTGCTCTTTG | AGCGCTCGGTTCACCGCGCCCATGATTAATAG |
| NDM | AGCGCTCGGTTCATATCACCGTTGGGATCGAC | AGCGCTCGGTTCTAGTGCTCAGTGTCGGCATC |

**Supplementary Table 5. Standard DNA barcode primer sequences**. Barcode number (BC) for barcodes used in sequencing, barcode name (Barcode name) and sequence (Sequence) of standard DNA barcode primers used in this work. All sequences shown in 5′ to 3′ direction. BC11-BC20 contain 12-nt barcode sequences, while NB01-NB10 contain 24-nt barcode sequences.

| **BC** | **Barcode name** | **Sequence** |
| --- | --- | --- |
| - | BC11-5RKB009-Homo-2P-Std | AAGAACCGAGCCAGCGCTCGGTTC |
| - | BC12-5RKB025-Homo-2P-Std | ACGCTGAAGAATAGCGCTCGGTTC |
| - | BC13-5RKB029-Homo-2P-Std | AGATTCCGCCGTAGCGCTCGGTTC |
| - | BC14-5RKB033-Homo-2P-Std | ATCGTCGTGCCTAGCGCTCGGTTC |
| - | BC15-5RKB035-Homo-2P-Std | ATTCATACCAGCAGCGCTCGGTTC |
| - | BC16-7RKB036-Homo-2P-Std | CAATACGCTGCAAGCGCTCGGTTC |
| - | BC17-7RKB056-Homo-2P-Std | CTGAACTCCGCAAGCGCTCGGTTC |
| - | BC18-7RKB066-Homo-2P-Std | GCTGCAATATACAGCGCTCGGTTC |
| - | BC19-7RKB074-Homo-2P-Std | TAGTGACGATGGAGCGCTCGGTTC |
| - | BC20-7RKB77-2Homo-2P-Std | TCCGTATTAGCCAGCGCTCGGTTC |
| 1 | AMR-NB01-24BC-Std | CACAAAGACACCGACAACTTTCTTAGCGCTCGGTTC |
| 2 | AMR-NB02-24BC-Std | ACAGACGACTACAAACGGAATCGAAGCGCTCGGTTC |
| 3 | AMR-NB03-24BC-Std | CCTGGTAACTGGGACACAAGACTCAGCGCTCGGTTC |
| 4 | AMR-NB04-24BC-Std | TAGGGAAACACGATAGAATCCGAAAGCGCTCGGTTC |
| 5 | AMR-NB05-24BC-Std | AAGGTTACACAAACCCTGGACAAGAGCGCTCGGTTC |
| 6 | AMR-NB06-24BC-Std | GACTACTTTCTGCCTTTGCGAGAAAGCGCTCGGTTC |
| 7 | AMR-NB07-24BC-Std | AAGGATTCATTCCCACGGTAACACAGCGCTCGGTTC |
| 8 | AMR-NB08-24BC-Std | ACGTAACTTGGTTTGTTCCCTGAAAGCGCTCGGTTC |
| 9 | AMR-NB09-24BC-Std | AACCAAGACTCGCTGTGCCTAGTTAGCGCTCGGTTC |
| 10 | AMR-NB10-24BC-Std | GAGAGGACAAAGGTTTCAACGCTTAGCGCTCGGTTC |

| **Supplementary Table 6. Full sequences of synthetic templates used in this work.** Gene target name (Target), length of the synthetic templates in bp (Length), and full sequence (Sequence) of synthetic templates used in this work. LT, ipaH, G18S, and ITS1 were ordered as two oligos (noted as target_F and target_R); all other targets were ordered as IDT gBlock Gene Fragments. Extra bases were added as padding to targets shorter than 125 base pairs to meet the minimum gBlock length requirement. All sequences are shown in the 5′ to 3′ direction. | | |
| --- | --- | --- |
| **Target** | **Length** | **Sequence** |
| stx1 | 132 | ACTTCTCGACTGCAAAGACGTATGTAGATTCGCTGAATGTCATTCGCTCTGCAATAGGTACTCCATTACAGACTATTTCATCAGGAGGTACGTCTTTACTGATGATTGATAGTGGCACAGGGGATAATTTGT |
| stx2 | 125 | CCACATCGGTGTCTGTTATTAACCACACCCCACCGGGCAGTTATTTTGCTGTGGATATACGAGGGCTTGATGTCTATCAGGCGCGTTTTGACCACATTAATTGCGTTGCGCTCACTGCCCGCTTT |
| aaiC | 215 | ATTGTCCTCAGGCATTTCACGCTTTTTCAGGAATTGACGGTACTGTTTTTGATTTAATTAATTTGAAGCTTAGGGTTACTAAACACATACAAGACCTTCTGGAGAACTTTTTTAAGAGAGGTGAAAAAGAAGTTAAAATAGAAATTTTACGTAGGGAATCAACTAAATCAGGTAGTGCATACTCATCATTTAAGGTTGTTTATCAGGGGTGTCGT |
| aatA | 237 | CTGGCGAAAGACTGTATCATTGATAATTTCTTTCAGAAAAGCATCCAGTTTAATTCTTATTCTCTTGATATCGAAGAGTTAGATATTAATAAACATAACAATATAAAAACGATGTTACCAGATATAAATATAGGGTTAGGGCAGTATATAAACAACAATCAATGGTTCTCATCTATTACAGACAGCCATTTTTATTTATCATTATCCTATAATCTTCTATCGGCTTATGAAGCAAAA |
| aggR | 125 | GCAATCAGATTAAGCAGCGATACATTAAGACGCCTAAAGGATGCCCTGATGATAATATACGGAATATCAAAAGTAGATGCTTGCAGTTGTCCGAAGCCTGGGGTGCCTAATGAGTGAGCTAACTC |
| eae | 125 | CATTGATCAGGATTTTTCTGGTGATAATACCCGTTTAGGTATTGGTGGCGAATACTGGCGAGACTATTTCAAAAGTAGCGTTAACGGCTATTTCCGCATGAGCCAGTCGGGAAACCTGTCGTGCC |
| bfpA | 125 | TGGTGCTTGCGCTTGCTGCAACCGTTACTGCCGGTGTGATGTTTTACTACCAGTCTGCGTCTGATTCCAATAAGTCACAGAATGCCATTTCAGAAGTAATGAGCGCAACGCCGCTTCCTCGCTCA |
| LT_F | 62 | TTCCCACCGGATCACCAAGCTTGGAGAGAAGAACCCTGGATTCATCATGCACCACAAGGTTG |
| LT_R | 62 | CAACCTTGTGGTGCATGATGAATCCAGGGTTCTTCTCTCCAAGCTTGGTGATCCGGTGGGAA |
| STh | 147 | GCTAAACCAGCAGGGTCTTCAAAAGAAAAAATTACACTAGAATCGAAAAAATGTAACATTGTAAAAAAAAATAATGAAAGTAGTCCTGAAAGCATGAATAGTAGCAATTACTGCTGTGAATTGTGTTGTAATCCTGCTTGTACCGGG |
| STp | 147 | GGCCGCGTTGCTGAATCACTTGACTCTTCAAAAGAGAAAATTACATTAGAGACTAAAAAGTGTGATGTTGTAAAAAACAACAGTGAAAAAAAATCAGAAAATATGAACAACACATTTTACTGCTGTGAACTTTGTTGTAATCCTGCC |
| ipaH_F | 64 | CCTTTTCCGCGTTCCTTGACCGCCTTTCCGATACCGTCTCTGCACGCAATACCTCCGGATTCCG |
| ipaH_R | 64 | CGGAATCCGGAGGTATTGCGTGCAGAGACGGTATCGGAAAGGCGGTCAAGGAACGCGGAAAAGG |
| virF | 125 | TCTGAGGAGGAGGTTTCTATCGATTTGTTCAAATCTATAAAAGAGATGCCTTTTGGCAAAAGAAAGATCTATAGTTTAGCTTGCCTTTTATCAGCTGTTTCAGCTGCATTAATGAATCGGCCAAC |
| ttr | 125 | CTCACCAGGAGATTACAACATGGCTAATTTAACCCGTCGTCAGTGGCTAAAAGTCGGTCTCGCCGTCGGTGGGATGGTCACTTTTGGTCTGAGCTCTGACTCGCTGCGCTCGGTCGTTCGGCTGC |
| hipO | 125 | CTTGCGGTCATGATGGACATACTACTTCTTTATTGCTTGCTGCAAAGTATTTAGCAAGTCAGAATTTTAATGGCACTTTAAATCTTTATTTTCAACCTGCTGAAGAGGGTTTGGGTGGTGCTGCG |
| GlyA | 125 | AAACCAAAGCTTATCGTGTGCGGTGCGAGTGCTTATGCTCGTATTATTGACTTTTCAAAATTTAGAGAGATTGCGGATGAAGTTGGAGCTTATCTTTTTGCAGACATTGCACACATTGCTGGACT |
| CR18S | 126 | GGGTTGTATTTATTAGATAAAGAACCAATATAATTGGTGACTCATAATAACTTTACGGATCACATTAAATGTGACATATCATTCAAGTTTCTGACCTATCAGCTTTAGACGGTAGGGTATTGGCCT |
| G18S_F | 63 | GACGGCTCAGGACAACGGTTGCACCCCCCGCGGCGGTCCCTGCTAGCCGGACACCGCTGGCAA |
| G18S_R | 63 | TTGCCAGCGGTGTCCGGCTAGCAGGGACCGCCGCGGGGGGTGCAACCGTTGTCCTGAGCCGTC |
| ITS1_F | 88 | GTAATAGCAGTCGGCGGTTTCTTTTTTTTTGGCGGACAATTGCATGCGATTTGCTATGTGTTGAGGGAGAATAGGTGGCATGTTGGGC |
| ITS1_R | 88 | GCCCAACATGCCACCTATTCTCCCTCAACACATAGCAAATCGCATGCAATTGTCCGCCAAAAAAAAAGAAACCGCCGACTGCTATTAC |
| mcr-1 | 125 | GATCGCTGTCGTGCTCTTTGGCGCGATGCTACTGATCACCACGCTGTTATCATCGTATCGCTATGTGCTAAAGCCTGTGTTGATTTTGCTATTAATCATGGGCGCGGTCGGGGAGAGGCGGTTTG |
| NDM | 125 | ATATCACCGTTGGGATCGACGGCACCGACATCGCTTTTGGTGGCTGCCTGATCAAGGACAGCAAGGCCAAGTCGCTCGGCAATCTCGGTGATGCCGACACTGAGCACTACGTATTGGGCGCTCTT |

**Supplementary Table 7. Wastewater, soil, and fecal samples used in this work.** Sample name (Name), sample matrix type (Matrix Type), template input from each sample for PCR (PCR Input), and template input from each sample for TAC (TAC Input) for all wastewater, soil, and fecal samples used in this work.

| **Name** | **Matrix Type** | **PCR Input** | **TAC Input** |
| --- | --- | --- | --- |
| WW-1 | Wastewater | 100 ng | 1400 ng |
| WW-2 | Wastewater | 100 ng | 1400 ng |
| WW-3 | Wastewater | 100 ng | 1400 ng |
| WW-4 | Wastewater | 100 ng | 1400 ng |
| WW-5 | Wastewater | 100 ng | 1400 ng |
| WW-6 | Wastewater | 100 ng | 1400 ng |
| WW-7 | Wastewater | 100 ng | 1400 ng |
| WW-8 | Wastewater | 100 ng | 1400 ng |
| WW-9 | Wastewater | 100 ng | 1400 ng |
| WW-10 | Wastewater | 100 ng | 1400 ng |
| S-1 | Soil | 100 ng | 1400 ng |
| S-2 | Soil | 100 ng | 1400 ng |
| S-3 | Soil | 100 ng | 1400 ng |
| S-4 | Soil | 100 ng | 20 µL |
| S-5 | Soil | 100 ng | 1400 ng |
| S-6 | Soil | 100 ng | 1400 ng |
| S-7 | Soil | 100 ng | 1400 ng |
| S-8 | Soil | 100 ng | 20 µL |
| S-9 | Soil | 100 ng | 1400 ng |
| S-10 | Soil | 100 ng | 1400 ng |
| F-1 | Feces | 100 ng | 20 µL |
| F-2 | Feces | 100 ng | 20 µL |
| F-3 | Feces | 100 ng | 20 µL |
| F-4 | Feces | 100 ng | 20 µL |
| F-5 | Feces | 100 ng | 20 µL |
| F-6 | Feces | 5 µL | 20 µL |
| F-7 | Feces | 100 ng | 20 µL |
| F-8 | Feces | 5 µL | 20 µL |
| F-9 | Feces | 100 ng | 20 µL |
| F-10 | Feces | 100 ng | 20 µL |

**Supplementary Table 8. Nanopore sequencing run overview.** Run IDs (Run ID), contents of run (Run contents), whether the primer type used was SAMRS-AEGIS (S/A) or standard DNA (Primer (S/A or std)), barcodes used (BC), and read count (Reads) for the nanopore runs in this work. All sequencing was done using R10.4.1 MinION flowcells.

| **Run ID** | **Run contents** | **Primer (S/A or std)** | **BC** | **Reads** |
| --- | --- | --- | --- | --- |
| SA_synth | synthetic | S/A, std | 4,5,9,10 | 20.48 M |
| SA_ww_fecal | Wastewater, fecal | S/A | 1-10 | 13.43 M |
| SA_ww_fecal_set2 | Wastewater, fecal | S/A | 1-10 | 11.19 M |
| std_4ww_3fecal_3soil | Wastewater, fecal, soil | std | 1-10 | 12.87 M |
| std_5soil_5ww | Soil, wastewater | std | 1-10 | 13.59 M |
| SA_10soil_100ng | Soil | S/A | 1-10 | 13.50 M |
| SA_std_NTC | No template controls for a subset of the samples | S/A, std | 1-10 | 74.81 k |

**Supplementary Table 9. Nanopore datasets uploaded to Sequence Reads Archive (SRA).** Sample name (Sample Name), the contents of each sample (Contents), and the accession number of each sample (Sample Accession) are listed. Demultiplexed FASTQ basecalls are provided for each sample. All samples can be found under SRA Bioproject PRJNA1150247 - Nanopore sequencing data for 20-plex PCR assays using SAMRS-AEGIS primers for target detection in environmental and fecal matrices.

| **Sample Name** | **Contents** | **Sample Accession** |
| --- | --- | --- |
| Standard_20  synthetic_10cpuL | Standard DNA 20-plex assay on 20 synthetic templates at 10 cp/uL. | SAMN43275846 |
| Standard_20  synthetic_10_4cpuL | Standard DNA 20-plex assay on 20 synthetic templates at 1×10^4^ cp/uL. | SAMN43275847 |
| SA_20synthetic_  10cpuL | SAMRS-AEGIS 20-plex assay on 20 synthetic templates at 10 cp/uL. | SAMN43275848 |
| SA_20synthetic_  10_4cpuL | SAMRS-AEGIS 20-plex assay on 20 synthetic templates at 1×10^4^ cp/uL. | SAMN43275849 |
| SA_WW1 | SAMRS-AEGIS 20-plex assay on wastewater sample 1. | SAMN43275462 |
| SA_WW2 | SAMRS-AEGIS 20-plex assay on wastewater sample 2. | SAMN43275463 |
| SA_WW3 | SAMRS-AEGIS 20-plex assay on wastewater sample 3. | SAMN43275464 |
| SA_WW4 | SAMRS-AEGIS 20-plex assay on wastewater sample 4. | SAMN43275465 |
| SA_WW5 | SAMRS-AEGIS 20-plex assay on wastewater sample 5. | SAMN43275466 |
| SA_WW6 | SAMRS-AEGIS 20-plex assay on wastewater sample 6. | SAMN43275467 |
| SA_WW7 | SAMRS-AEGIS 20-plex assay on wastewater sample 7. | SAMN43275468 |
| SA_WW8 | SAMRS-AEGIS 20-plex assay on wastewater sample 8. | SAMN43275469 |
| SA_WW9 | SAMRS-AEGIS 20-plex assay on wastewater sample 9. | SAMN43275470 |
| SA_WW10 | SAMRS-AEGIS 20-plex assay on wastewater sample 10. | SAMN43275471 |
| SA_S1 | SAMRS-AEGIS 20-plex assay on soil sample 1. | SAMN43275472 |
| SA_S2 | SAMRS-AEGIS 20-plex assay on soil sample 2. | SAMN43275473 |
| SA_S3 | SAMRS-AEGIS 20-plex assay on soil sample 3. | SAMN43275474 |
| SA_S4 | SAMRS-AEGIS 20-plex assay on soil sample 4. | SAMN43275475 |
| SA_S5 | SAMRS-AEGIS 20-plex assay on soil sample 5. | SAMN43275476 |
| SA_S6 | SAMRS-AEGIS 20-plex assay on soil sample 6. | SAMN43275477 |
| SA_S7 | SAMRS-AEGIS 20-plex assay on soil sample 7. | SAMN43275478 |
| SA_S8 | SAMRS-AEGIS 20-plex assay on soil sample 8. | SAMN43275479 |
| SA_S9 | SAMRS-AEGIS 20-plex assay on soil sample 9. | SAMN43275480 |
| SA_S10 | SAMRS-AEGIS 20-plex assay on soil sample 10. | SAMN43275481 |
| SA_F1 | SAMRS-AEGIS 20-plex assay on fecal sample 1. | SAMN43275482 |
| SA_F2 | SAMRS-AEGIS 20-plex assay on fecal sample 2. | SAMN43275483 |
| SA_F3 | SAMRS-AEGIS 20-plex assay on fecal sample 3. | SAMN43275484 |
| SA_F4 | SAMRS-AEGIS 20-plex assay on fecal sample 4. | SAMN43275485 |
| SA_F5 | SAMRS-AEGIS 20-plex assay on fecal sample 5. | SAMN43275486 |
| SA_F6 | SAMRS-AEGIS 20-plex assay on fecal sample 6. | SAMN43275487 |
| SA_F7 | SAMRS-AEGIS 20-plex assay on fecal sample 7. | SAMN43275488 |
| SA_F8 | SAMRS-AEGIS 20-plex assay on fecal sample 8. | SAMN43275489 |
| SA_F9 | SAMRS-AEGIS 20-plex assay on fecal sample 9. | SAMN43275490 |
| SA_F10 | SAMRS-AEGIS 20-plex assay on fecal sample 10. | SAMN43275491 |
| std_WW1 | Standard DNA 20-plex assay on wastewater sample 1. | SAMN43275492 |
| std_WW2 | Standard DNA 20-plex assay on wastewater sample 2. | SAMN43275493 |
| std_WW3 | Standard DNA 20-plex assay on wastewater sample 3. | SAMN43275494 |
| std_WW4 | Standard DNA 20-plex assay on wastewater sample 4. | SAMN43275495 |
| std_WW5 | Standard DNA 20-plex assay on wastewater sample 5. | SAMN43275496 |
| std_WW7 | Standard DNA 20-plex assay on wastewater sample 7. | SAMN43275497 |
| std_WW8 | Standard DNA 20-plex assay on wastewater sample 8. | SAMN43275498 |
| std_WW9 | Standard DNA 20-plex assay on wastewater sample 9. | SAMN43275499 |
| std_WW10 | Standard DNA 20-plex assay on wastewater sample 10. | SAMN43275500 |
| std_S1 | Standard DNA 20-plex assay on soil sample 1. | SAMN43275501 |
| std_S2 | Standard DNA 20-plex assay on soil sample 2. | SAMN43275502 |
| std_S3 | Standard DNA 20-plex assay on soil sample 3. | SAMN43275503 |
| std_S4 | Standard DNA 20-plex assay on soil sample 4. | SAMN43275504 |
| std_S5 | Standard DNA 20-plex assay on soil sample 5. | SAMN43275505 |
| std_S6 | Standard DNA 20-plex assay on soil sample 6. | SAMN43275506 |
| std_S7 | Standard DNA 20-plex assay on soil sample 7. | SAMN43275507 |
| std_S8 | Standard DNA 20-plex assay on soil sample 8. | SAMN43275508 |
| std_F2 | Standard DNA 20-plex assay on fecal sample 2. | SAMN43275509 |
| std_F5 | Standard DNA 20-plex assay on fecal sample 5. | SAMN43275510 |
| std_F10 | Standard DNA 20-plex assay on fecal sample 10. | SAMN43275511 |
| SA_F1_NTC | No template control for SAMRS-AEGIS 20-plex assay on fecal sample 1. | SAMN43275850 |
| SA_S3_NTC | No template control for SAMRS-AEGIS 20-plex assay on soil sample 3. | SAMN43275851 |
| SA_F8_NTC | No template control for SAMRS-AEGIS 20-plex assay on fecal sample 8. | SAMN43275852 |
| SA_S4_NTC | No template control for SAMRS-AEGIS 20-plex assay on soil sample 4. | SAMN43275853 |
| std_S5_NTC | No template control for standard DNA 20-plex assay on soil sample 5. | SAMN43275854 |
| SA_WW6_NTC | No template control for SAMRS-AEGIS 20-plex assay on wastewater sample 6. | SAMN43275855 |
| std_F10_NTC | No template control for standard DNA 20-plex assay on fecal sample 10. | SAMN43275856 |
| std_WW3_NTC | No template control for standard DNA 20-plex assay on wastewater sample 3. | SAMN43275857 |
| SA_WW4_NTC | No template control for SAMRS-AEGIS 20-plex assay on wastewater sample 4. | SAMN43275858 |
| std_S8_NTC | No template control for standard DNA 20-plex assay on soil sample 8. | SAMN43275859 |

**Supplementary Table 10. Analysis comparing costs between Oxford nanopore sequencing, Illumina sequencing, and the TaqMan^TM^** **array card.** Method (Method), estimated capital cost (Capital Cost), the cost per sample (Per sample cost), and an estimated maximum number of samples that can be processed at once using each method (Number of Samples) are provided. Details and assumptions for this analysis can be found in **Supplementary File 2**.

| **Method** | **Capital Cost** | **Per sample cost^1^** | **Number of Samples^2^** |
| --- | --- | --- | --- |
| MinION | $1,999 | $79.93 | 10 |
| PromethION | $9,555 | $22.31 | 63 |
| Illumina NovaSeq | - | $11.22 | 4536 |

^1^: Per sample cost assuming the number of samples in "Number of Samples” column are processed.

^2^: Assumes ≈1.1 million reads per sample is required for sufficient coverage.

**Supplementary Figures**

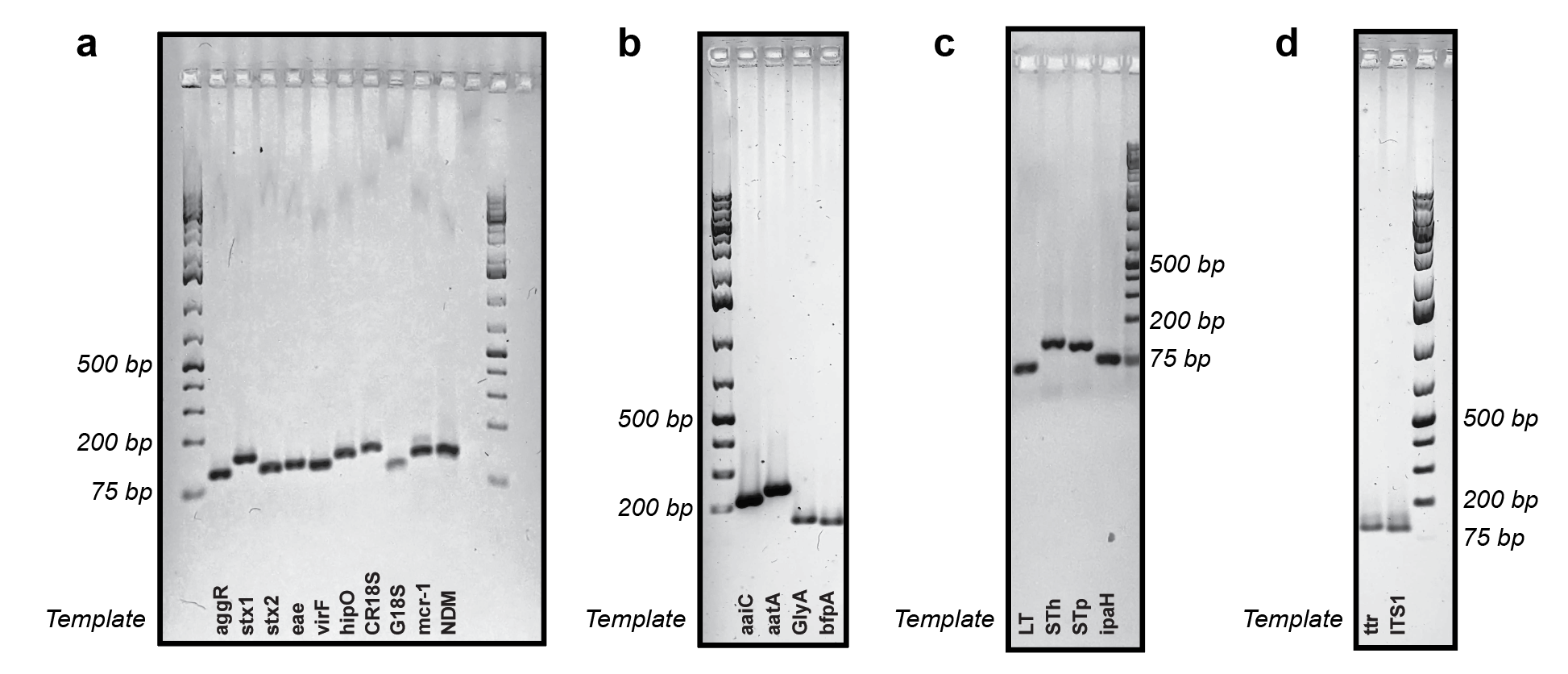

**Supplementary Figure 1. Amplification of targets using synthetic templates. (a-d)** Singleplex, one-pot PCR of all targets using synthetic templates (**Table S6**) using SAMRS-AEGIS primers from **Table S2**. Products were imaged on a 2% (w/v) agarose gel alongside GeneRuler 1kb Plus DNA Ladder.

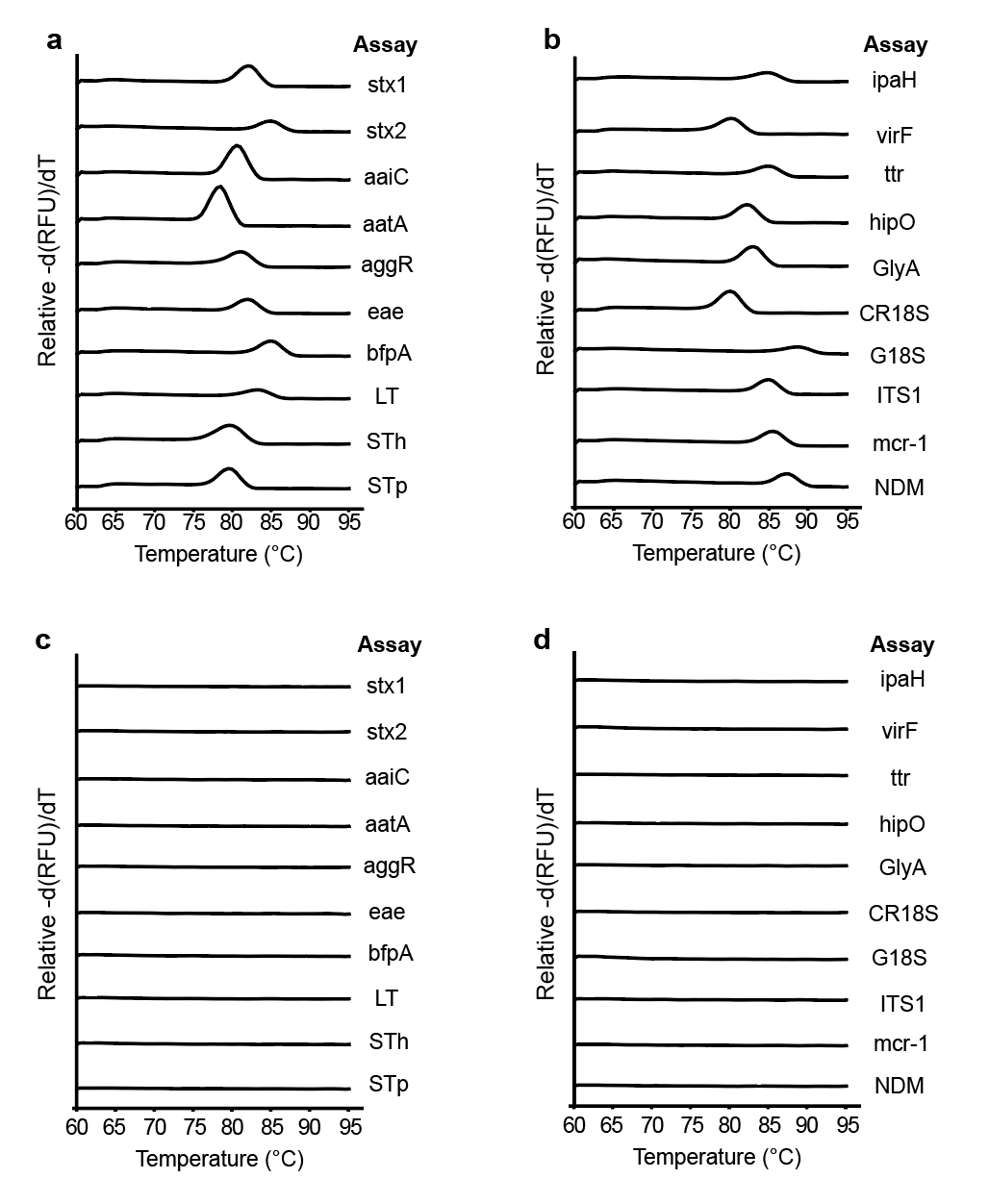

**Supplementary Figure 2. Melt curves of singleplex qPCR using SAMRS-AEGIS primers.** All 40 SAMRS-AEGIS primers (listed in **Table S2)** were used for singleplex amplification of the 20 targets from **Table 1**. All 20 targets were input at 5 × 10^3^ copies/µL, and one set of primers per assay was used for each reaction. (**a-b**) Singleplex melt curves for each assay and (**c-d**) singleplex melt curves for the no template control of each assay.

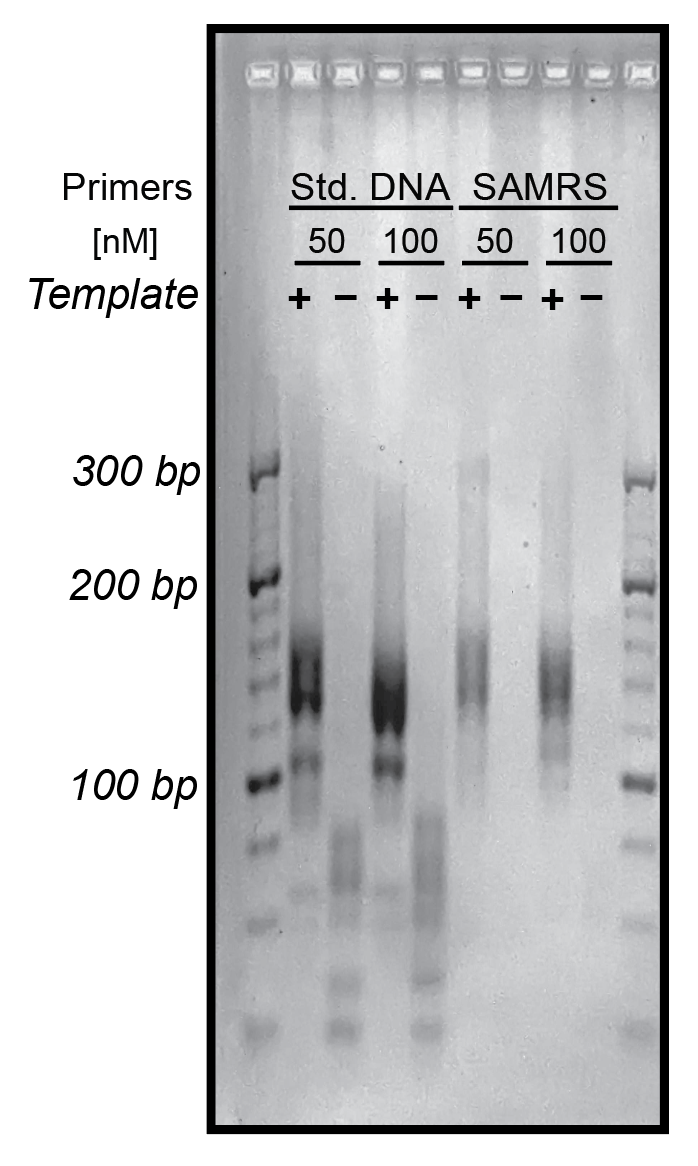

**Supplementary Figure 3. 20-plex PCR using synthetic templates.** 20-plex, one-pot PCR of all targets using synthetic templates (**Table S6**) at 0.5 ng per target (3.3 × 10^7^ - 1.3 × 10^8^ copies/µL). “Template” indicates the presence or absence of template DNA. Both standard DNA (Std. DNA) and SAMRS-AEGIS primers from **Table S2 and S4** were used at either 0.05 µM (50 nM) or 0.1 µM (100 nM), and 12-nt barcoding primers were used at 2 µM (**Table S3 and S5)**.

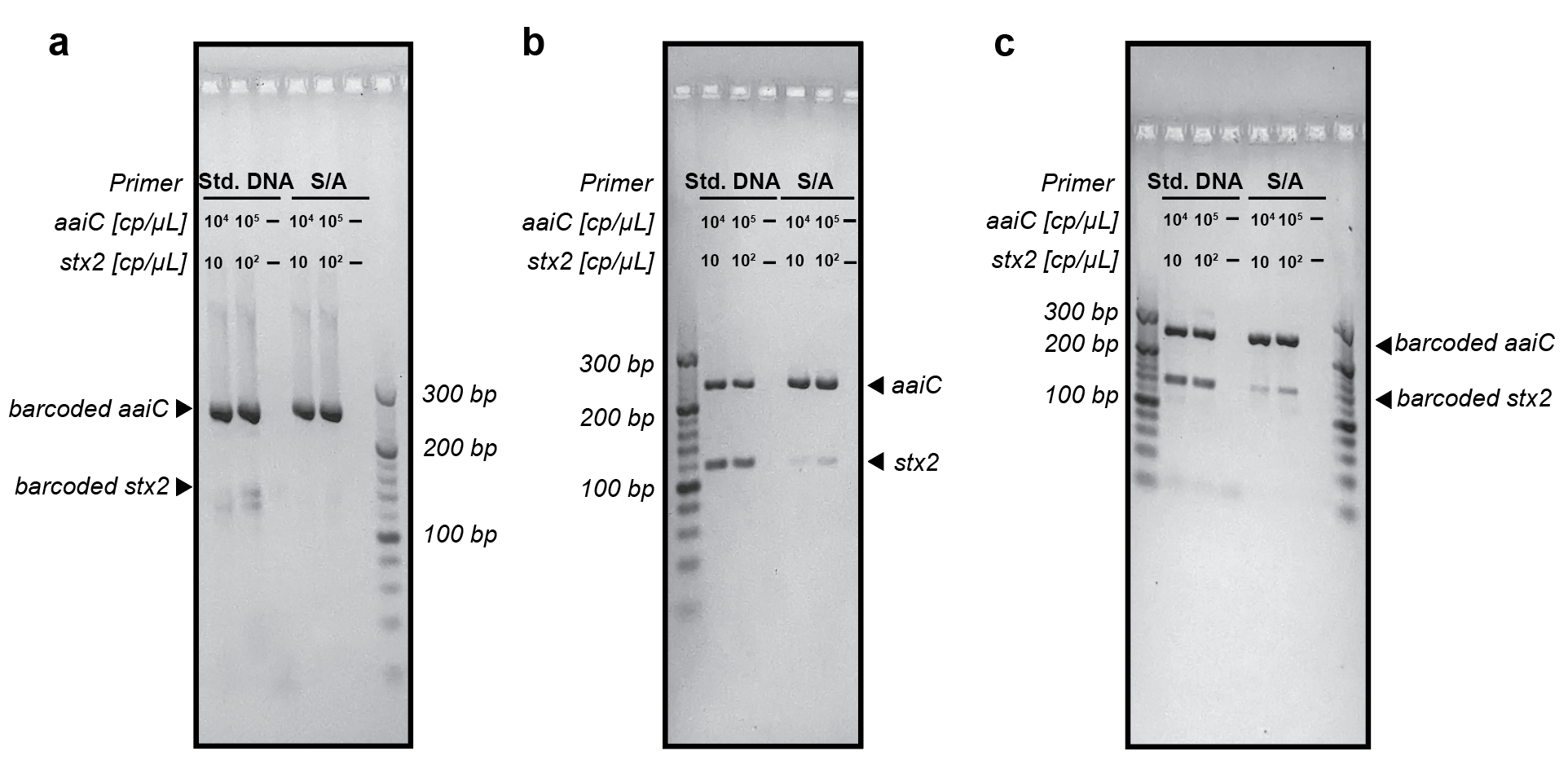

**Supplementary Figure 4. One-pot and sequential PCR to minimize amplification bias.** One-pot PCR and sequential PCR were used to amplify stx2 at 10 and 10^2^ copies/µL (cp/µL), and aaiC at 10^4^ and 10^5^ copies/µL in 20 µL. For sequential PCR, the second round of PCR was done in 20 µL of volume using 1 µL of first round PCR as template. ‘–’ lane shows no template control. Both standard DNA (Std. DNA) and SAMRS-AEGIS (S/A) primers were present at 0.1 µM **(Table S2, S4)**. PCR primers used had 12-nt barcodes (**Table S3, S5)**. (**a**) One-pot PCR amplification (35 cycles). Expected product sizes for fully barcoded amplicons are aaiC: 263 bp and stx2: 141 bp. (**b**) The first step of sequential PCR amplification (40 cycles). Expected amplicon sizes are aaiC: 239 bp and stx2: 117 bp. (**c**) The second step of sequential PCR amplification (15 cycles). Expected sizes for fully barcoded amplicons are aaiC: 263 bp and stx2: 141 bp.

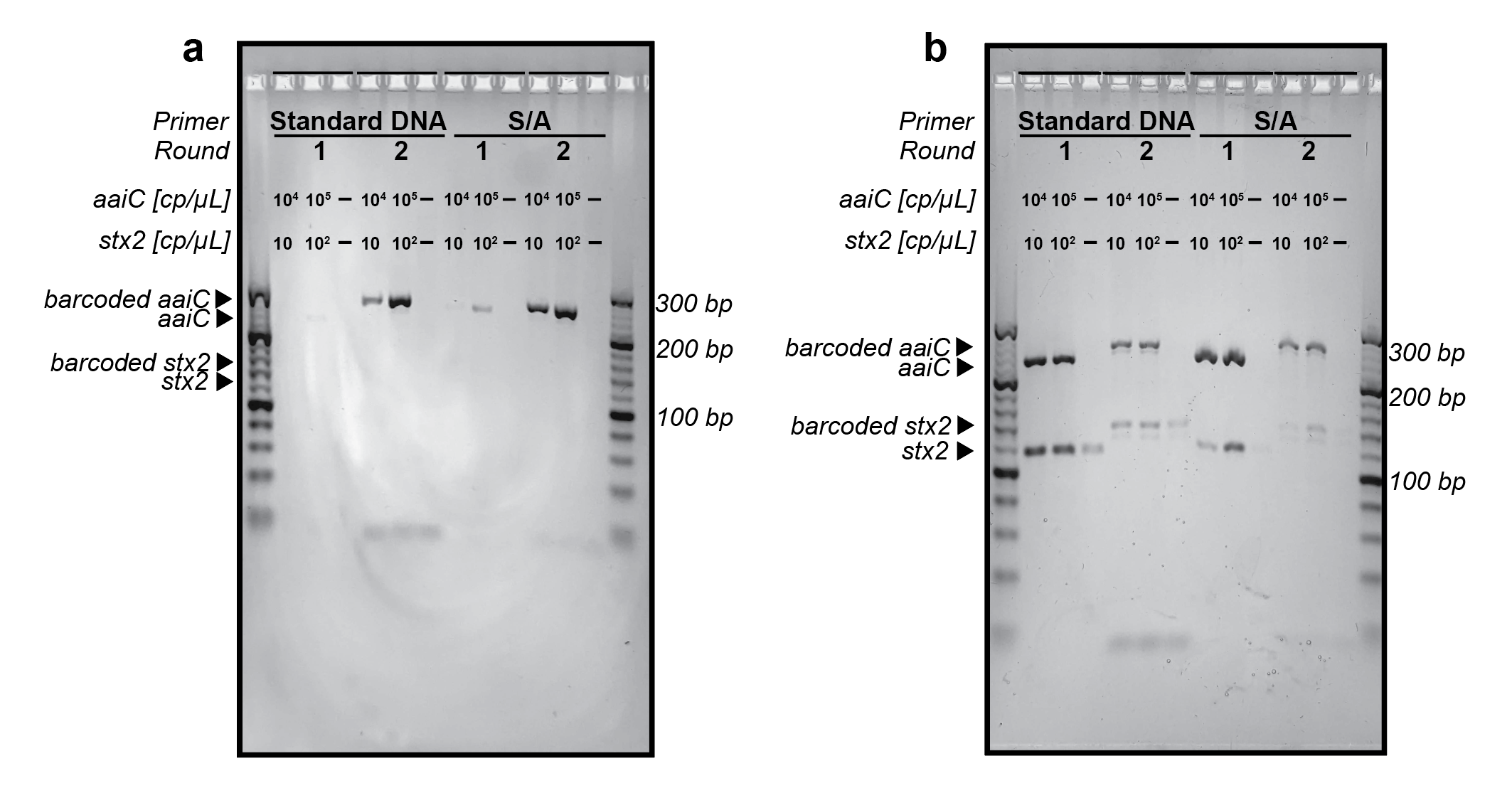

**Supplementary Figure 5. Sequential PCR cycling optimization to minimize amplification bias.** Sequential PCR was used to amplify stx2 at 10 and 10^2^ copies/µL (cp/µL), and aaiC at 10^4^ and 10^5^ copies/µL in 20 µL. The second round of PCR was performed in 20 µL of volume using 1 µL of first round PCR as template. ‘–’ lane shows no template control. Both standard DNA and SAMRS-AEGIS (S/A) primers were present at 0.1 µM **(Table S2, S4)**. Primers used had 12-nt barcodes (**Table S3, S5)**. (**a**) The first round of amplification consisted of 30 cycles, followed by 20 cycles of barcoding. (**b**) The first round of amplification consisted of 40 cycles, followed by 6 cycles of barcoding. For all reaction conditions, expected amplicon sizes after the first round are aaiC: 239 bp and stx2: 117 bp. Expected sizes for fully barcoded amplicons are aaiC: 263 bp and stx2: 141 bp.

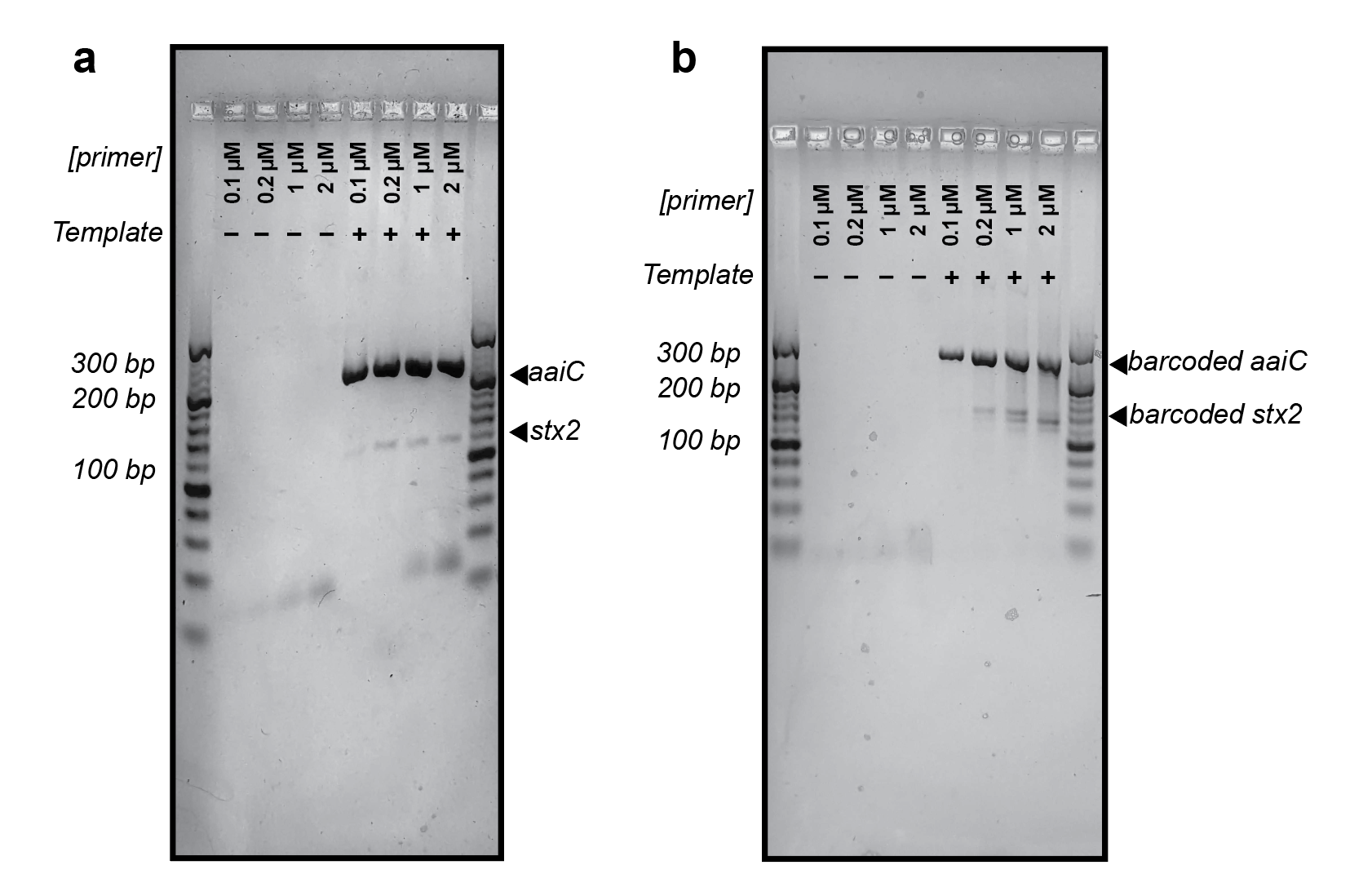

**Supplementary Figure 6. Primer concentration optimization in the first round of sequential PCR.** Sequential PCR with different SAMRS-AEGIS primer concentrations in the first round was used to amplify stx2 at 10 copies/µL, and aaiC at 1 × 10^4^ copies/µL. The second round of PCR was done in 20 µL using 1 µL of first round PCR as template. “Template” indicates presence or absence of template. (**a**) The first round of amplification at 0.1, 0.2, 1, and 2 µM of primer. For all reaction conditions, expected amplicon sizes after the first round are aaiC: 239 bp and stx2: 117 bp. (**b**) The second round of amplification was done using 2 µM of 12-nt barcoding primer **(Table S3, S5)** across all primer concentrations from the first round of amplification. Expected sizes for fully barcoded amplicons are aaiC: 263 bp and stx2: 141 bp.

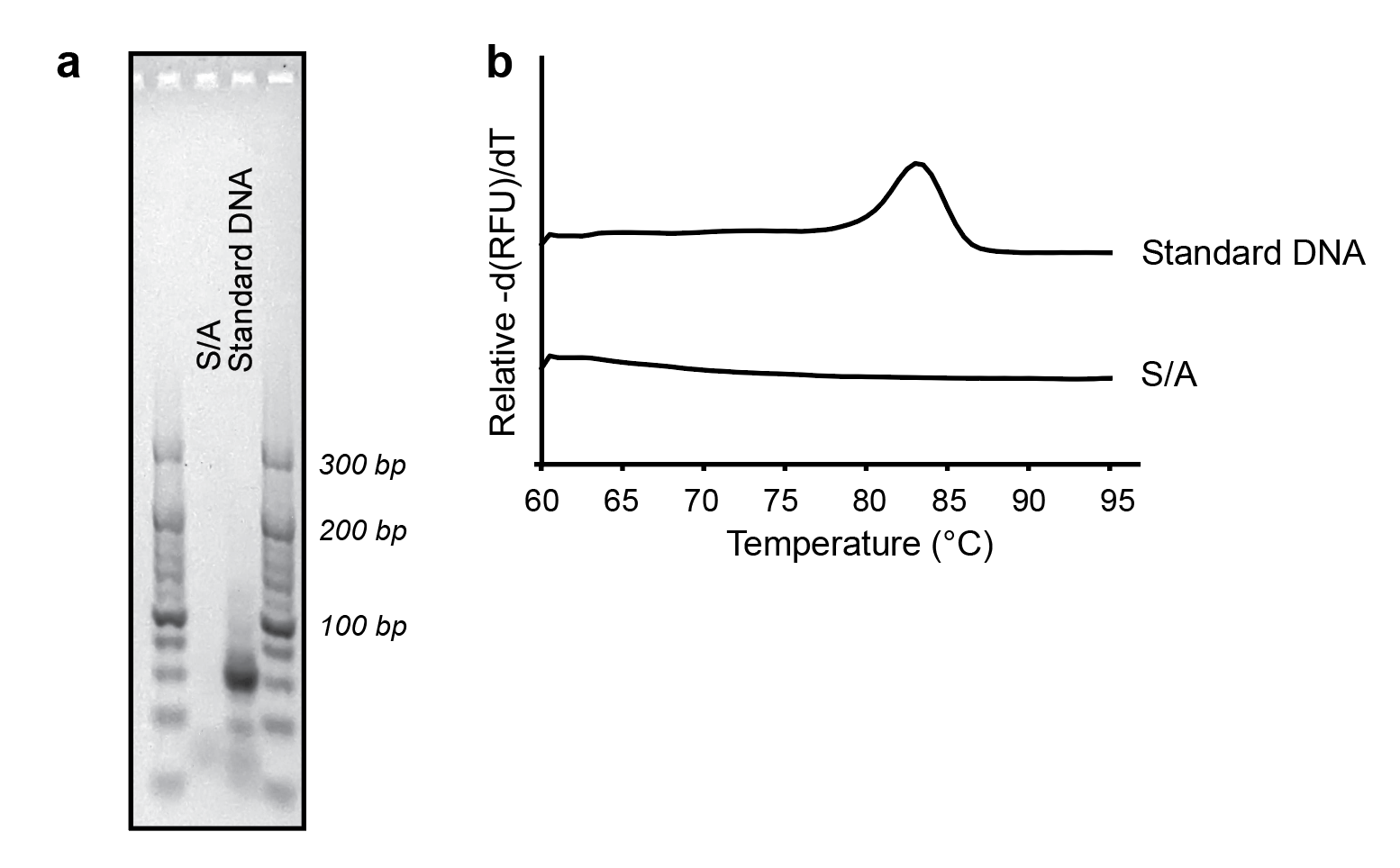

**Supplementary Figure 7. Comparison of 20-plex amplification in the absence of template using either SAMRS-AEGIS primers or standard DNA primers.** 20-plex PCR reactions contained mixture of 40 primers marked either as SAMRS-AEGIS (S/A) or standard DNA primers (**Table S2, S4**). No template was present in the reaction. (**a**) Agarose gel for 20-plex no template reactions with S/A and standard DNA primers. (**b**) Melt curves analysis for 20-plex no template reactions with S/A and standard DNA primers.

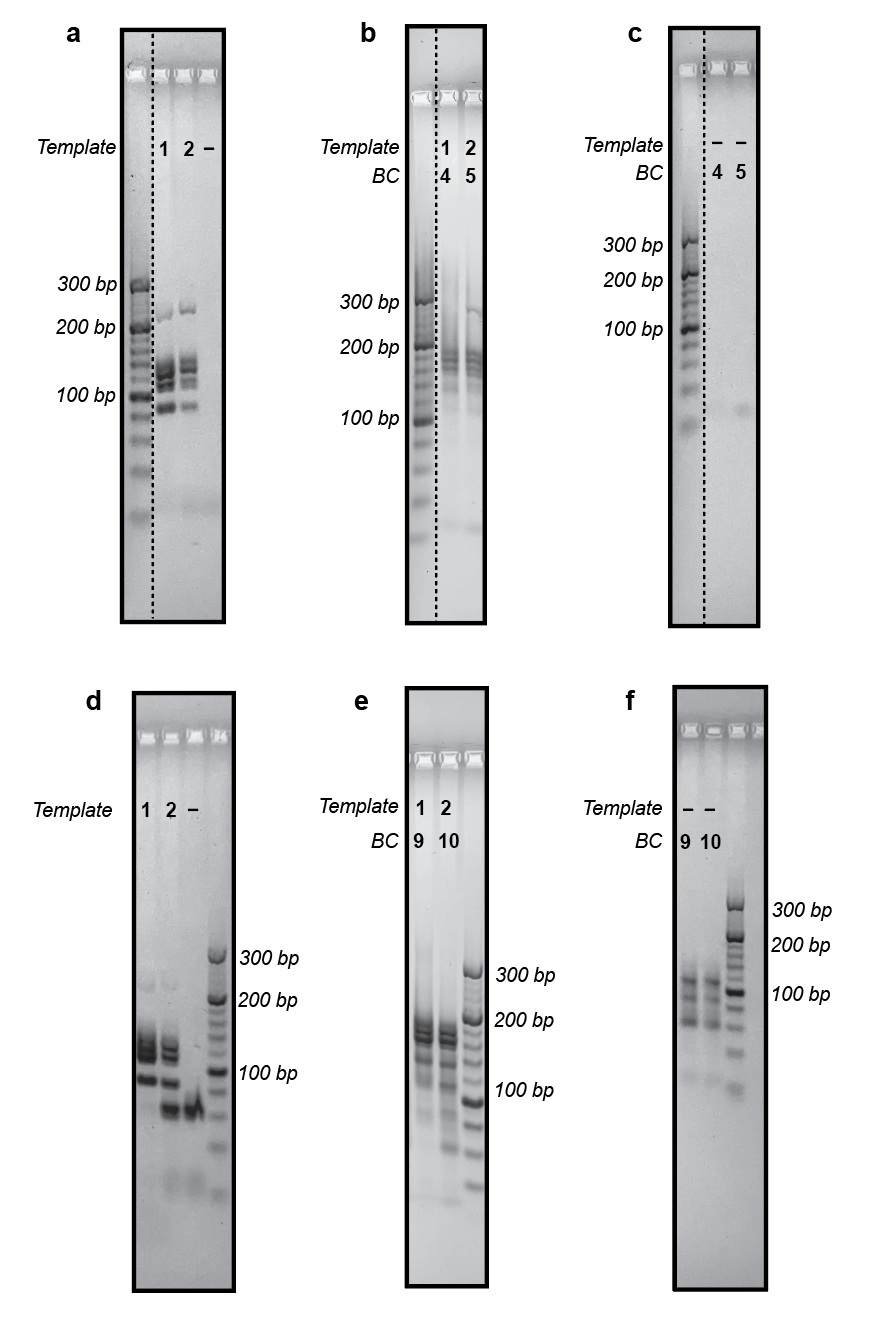

**Supplementary Figure 8. 20-plex PCR of all 20 targets using synthetic templates.** 20 pooled targets using synthetic templates (**Table S6**) were amplified using sequential PCR. The second round of PCR was performed in 20 µL of volume using 1 µL of first round PCR as template. Both standard DNA and SAMRS-AEGIS primers were assayed **(Tables S2, S4).** Lanes show template contents: 1) 20 pooled synthetic templates with each target at 1 × 10^4^ copies/µL, 2) 20 pooled synthetic templates with each target at 10 copies/µL. ’–’ lane shows no template control. Dashed line denotes discontinuity in gel image to show ladder lane. (**a**) The first round of amplification using SAMRS-AEGIS primers. (**b**) The second round of amplification using the AEGIS barcoding primers. (**c**) The no template controls for the second round of amplification using AEGIS barcoding primers. (**d**) The first round of amplification using standard DNA primers. Note that the primer dimerization band in the no template control disappears when there is a high concentration of synthetic templates present in the reaction. (**e**) The second round of amplification using standard DNA barcoding primers. (**f**) The no template controls for the second round of amplification using standard DNA barcoding primers. ‘*BC*’ indicates identity of the 24-nt barcode primer used **(Tables S3, S5**)**.**

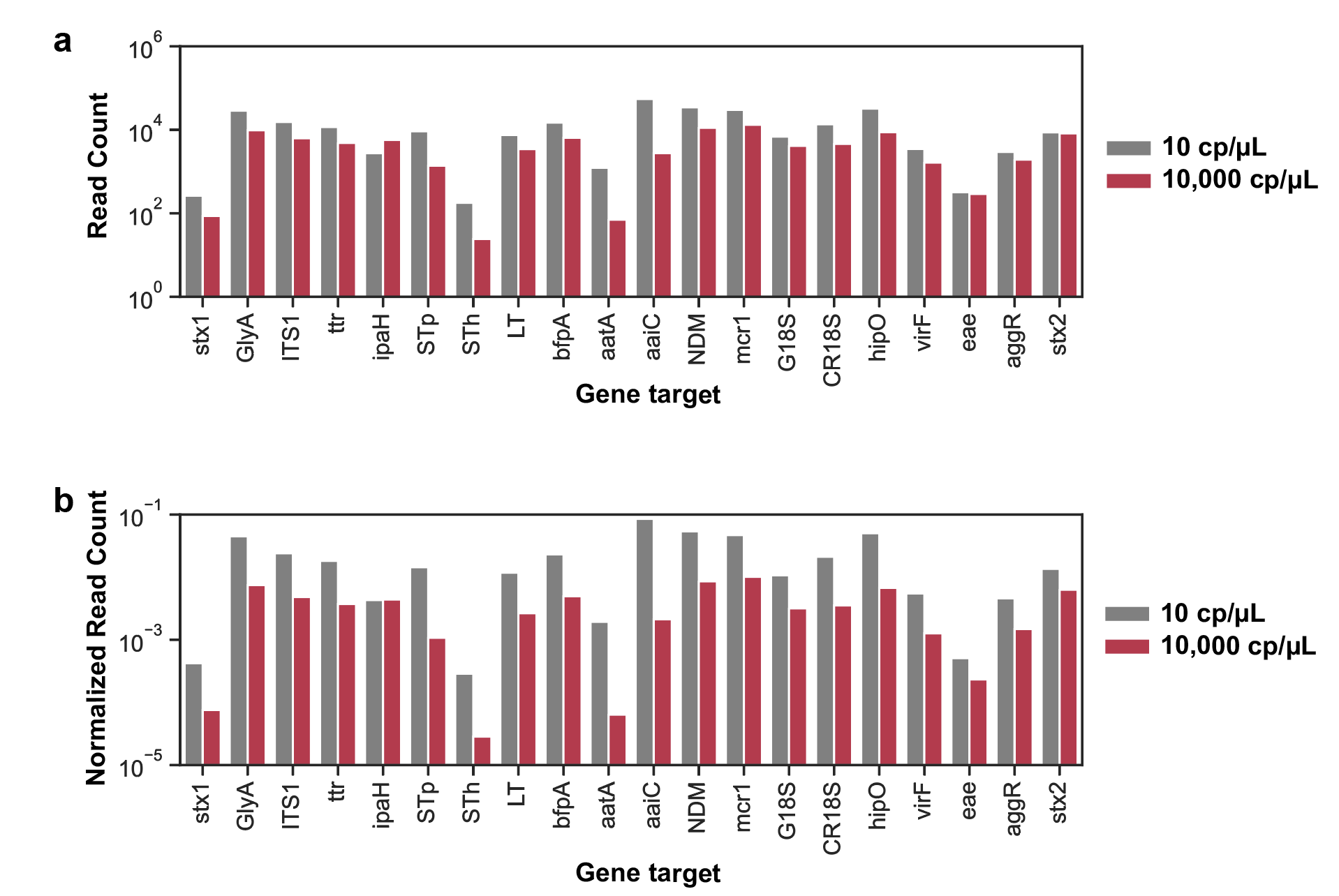

**Supplementary Figure 9. Raw and normalized read count for SAMRS-AEGIS 20-plex target amplification using synthetic templates.** All 20 synthetic templates used in this study were pooled for a total template input of 10 and 1 × 10^4^ copies/µL (cp/µL) for each target **(Table S6**)**,** amplified using SAMRS-AEGIS primers **(Figure S8a-c),** and sequenced using nanopore sequencing. (**a**) Total raw read count for all 20 targets. (**b**) Read count normalized to each barcode for all 20 targets.

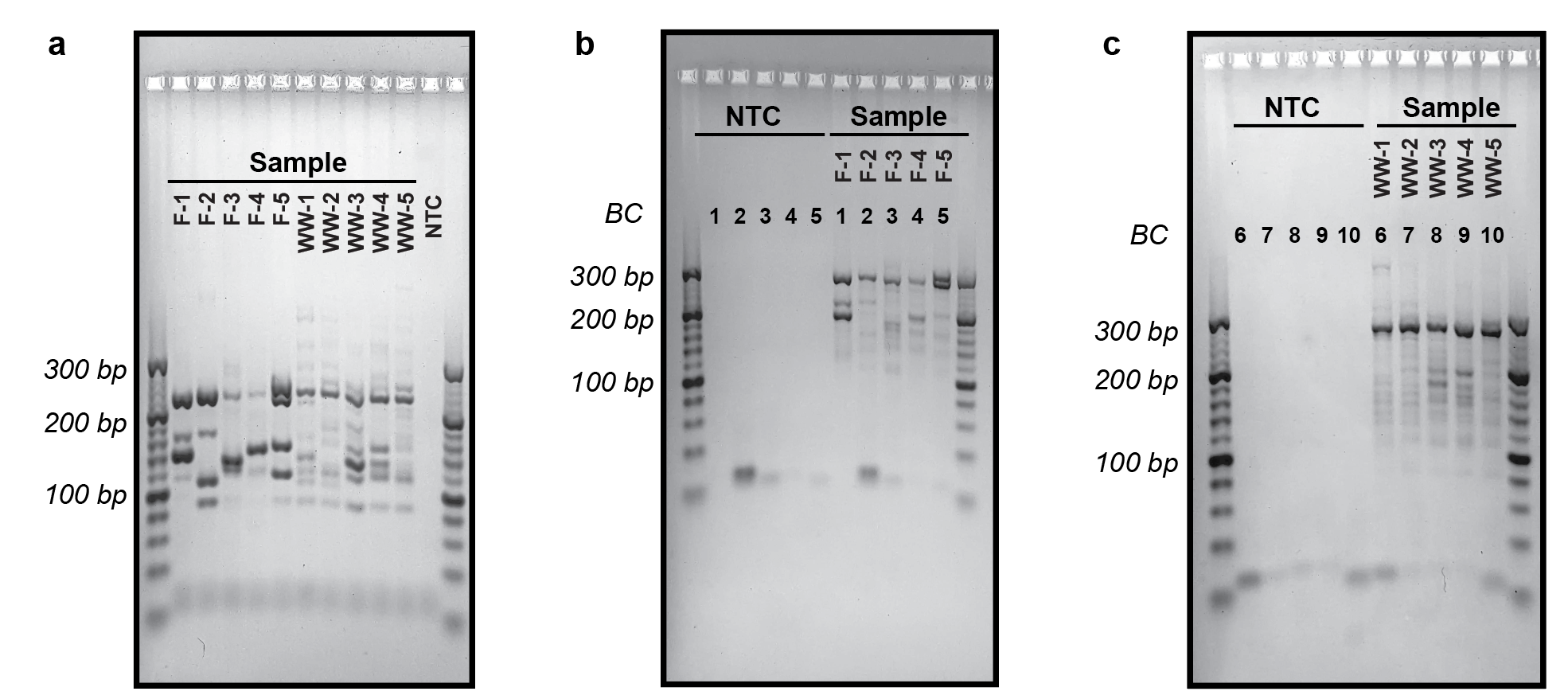

**Supplementary Figure 10. 20-plex PCR of 5 fecal (F-1 to F-5) and 5 wastewater (WW-1 to WW-5) samples using SAMRS-AEGIS primers.** 100 ng of DNA extracted from wastewater and fecal samples were amplified using SAMRS-AEGIS primers **(Tables S2, S7**)**.** ‘NTC’ lanes show no template controls. (**a**) The first round of amplification using SAMRS-AEGIS primers. (**b**) The second round of amplification for fecal samples using AEGIS barcoding primers. (**c**) The second round of amplification for wastewater samples using AEGIS barcoding primers. ‘*BC*’ indicates identity of the 24-nt barcode primer used **(Table S3**)**.**

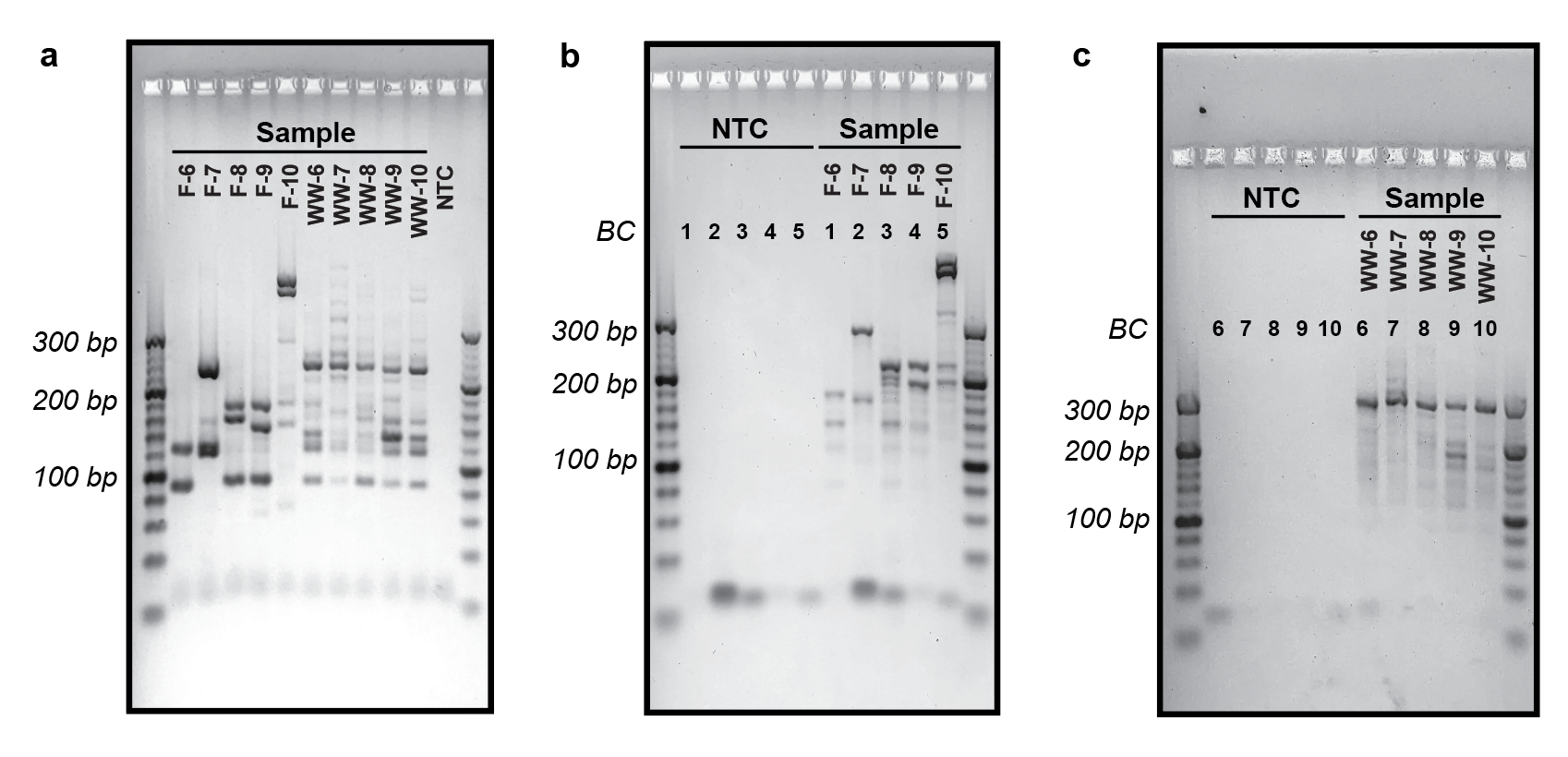

**Supplementary Figure 11. 20-plex PCR of 5 fecal (F-6 to F-10) and 5 wastewater (WW-6 to WW-10) samples using SAMRS-AEGIS primers.** Up to either 5 µL or 100 ng of DNA extracted from wastewater and fecal samples were amplified using SAMRS-AEGIS primers **(Tables S2, S7**)**.** ‘NTC’ lanes show no template controls. (**a**) The first round of amplification using SAMRS-AEGIS primers. (**b**) The second round of amplification for fecal samples using AEGIS barcoding primers. (**c**) The second round of amplification for wastewater samples using AEGIS barcoding primers. ‘*BC*’ indicates identity of the 24-nt barcode primer used **(Table S3**)**.**

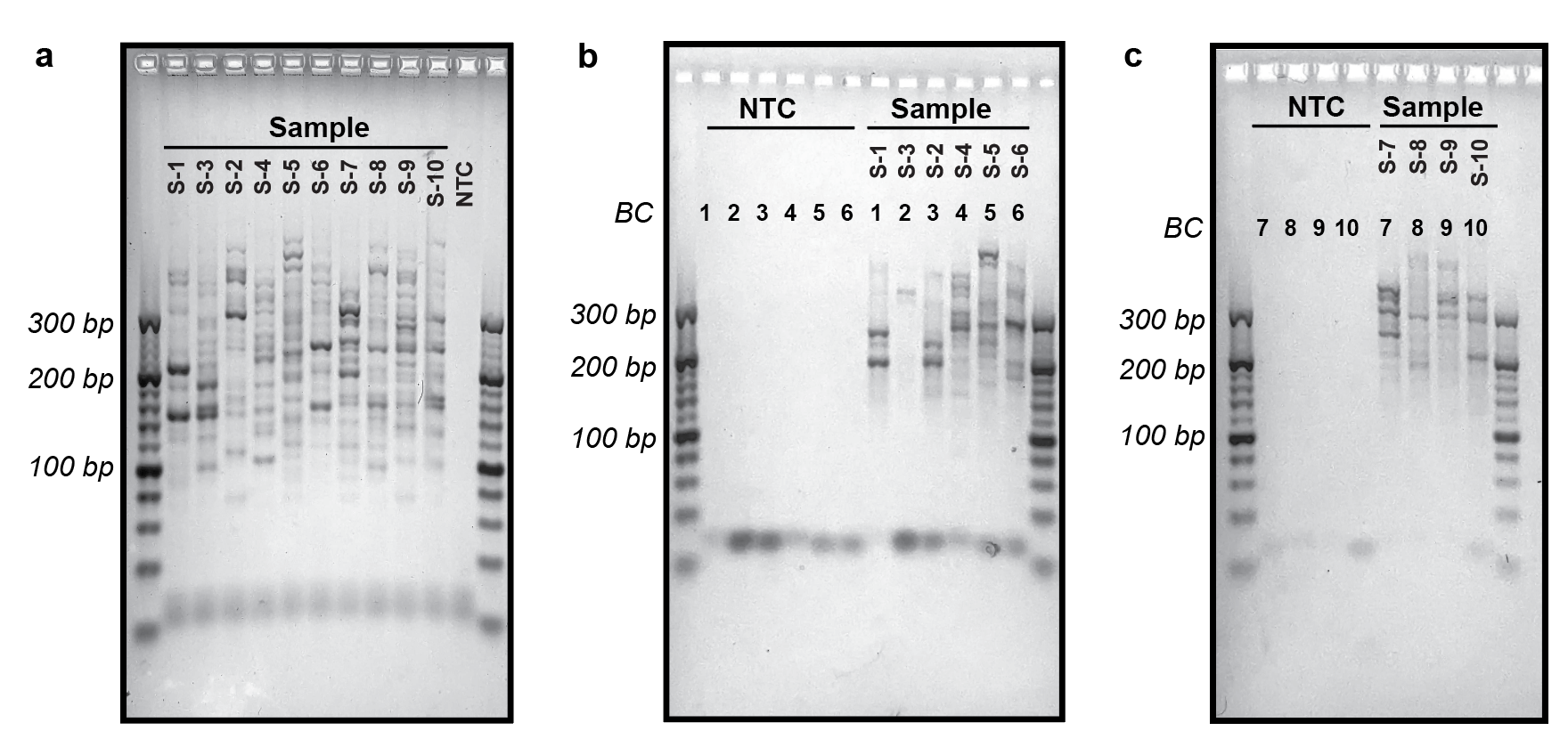

**Supplementary Figure 12. 20-plex PCR of 10 soil samples using SAMRS-AEGIS primers.** 100 ng of DNA extracted from soil samples were amplified using SAMRS-AEGIS primers **(Tables S2, S7**)**.** ‘NTC’ lanes show no template controls. (**a**) The first round of amplification using SAMRS-AEGIS primers. (**b-c**) The second round of amplification using AEGIS barcoding primers. ‘*BC*’ indicates identity of the 24-nt barcode primer used **(Table S3**)**.**

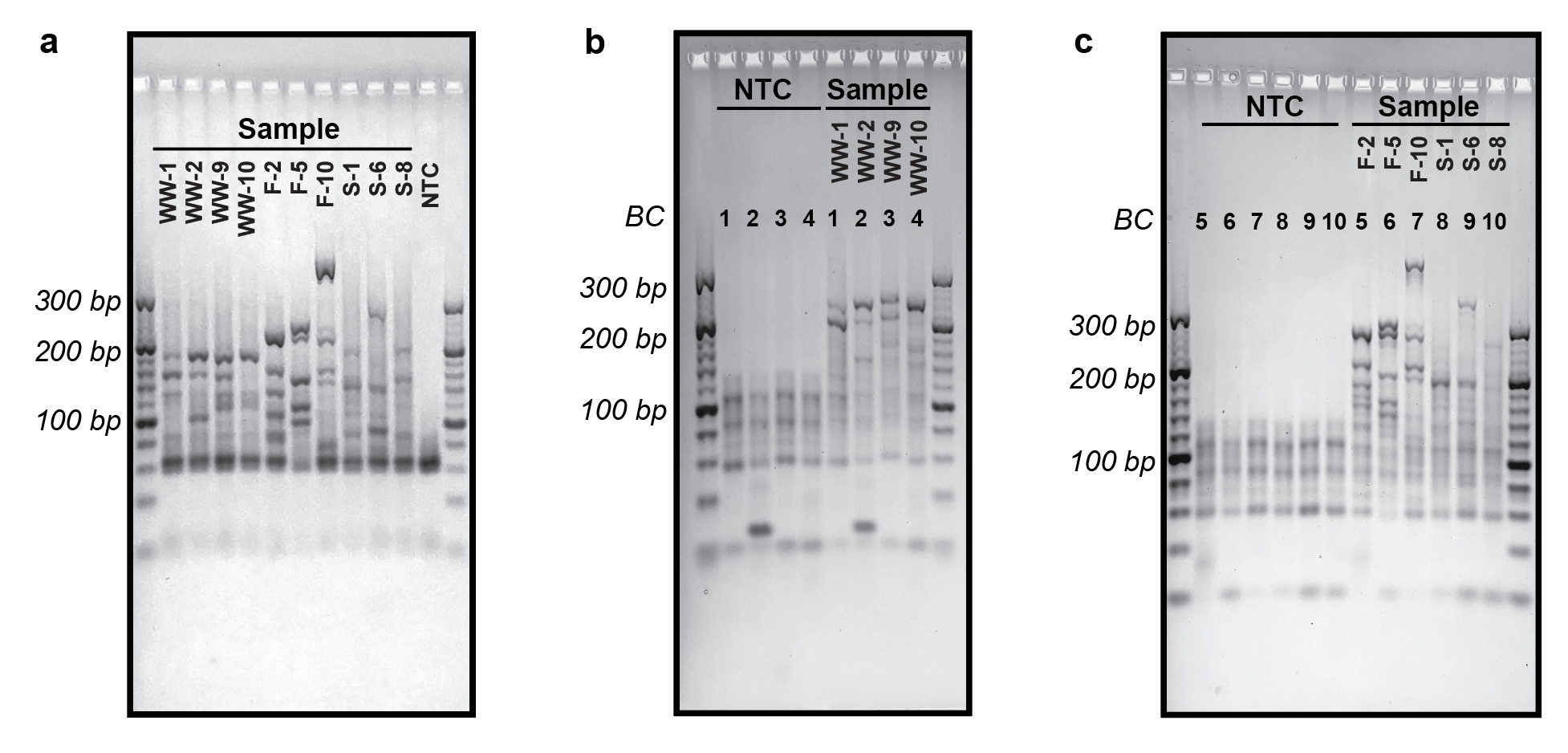

**Supplementary Figure 13. 20-plex PCR of 4 wastewater, 3 fecal, and 3 soil samples using standard DNA primers.** 100 ng of DNA extracted from wastewater, fecal, and soil samples were amplified using standard DNA primers **(Tables S4, S7**)**.** ‘NTC’ lanes show no template controls. (**a**) The first round of amplification using standard DNA primers. (**b**) The second round of amplification for wastewater samples using standard DNA barcoding primers. (**c**) The second round of amplification for fecal and soil samples using standard DNA barcoding primers. ‘*BC*’ indicates identity of the 24-nt barcode primer used **(Table S5**)**.**

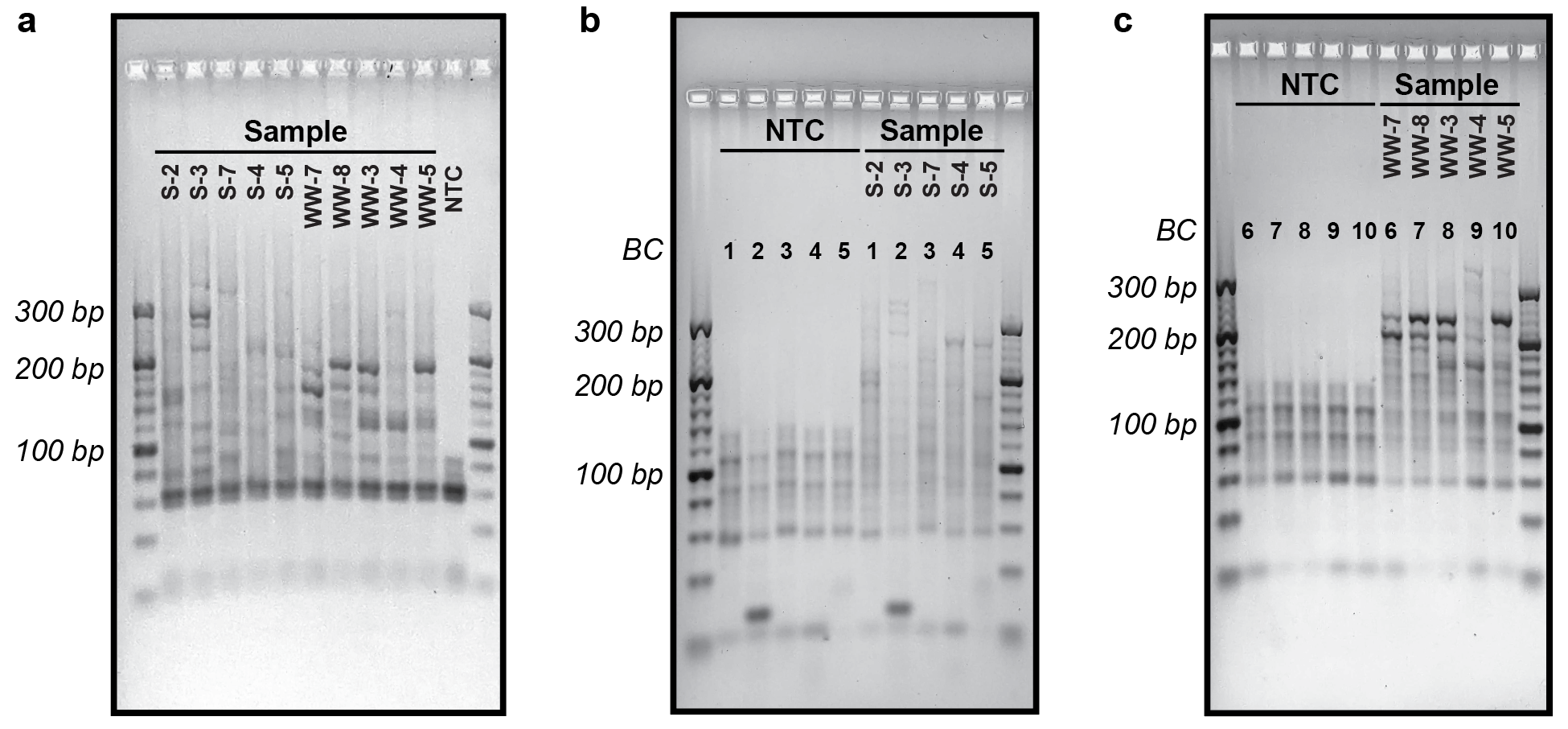

**Supplementary Figure 14. 20-plex PCR of 5 soil and 5 wastewater samples using standard DNA primers.** 100 ng of DNA extracted from soil and wastewater samples were amplified using standard DNA primers **(Tables S4, S7).** ‘NTC’ lanes show no template controls. (**a**) The first round of amplification using standard DNA primers. (**b**) The second round of amplification for soil samples using standard DNA barcoding primers. (**c**) The second round of amplification for wastewater samples using standard DNA barcoding primers. ‘*BC*’ indicates identity of the 24-nt barcode primer used **(Table S5**)**.**

**References.**

(1)	Liu, J.; Gratz, J.; Amour, C.; Kibiki, G.; Becker, S.; Janaki, L.; Verweij, J. J.; Taniuchi, M.; Sobuz, S. U.; Haque, R.; Haverstick, D. M.; Houpt, E. R. A Laboratory-Developed TaqMan Array Card for Simultaneous Detection of 19 Enteropathogens. *J. Clin. Microbiol.* **2013**, *51* (2), 472–480. <https://doi.org/10.1128/JCM.02658-12>.

(2)	Hidaka, A.; Hokyo, T.; Arikawa, K.; Fujihara, S.; Ogasawara, J.; Hase, A.; Hara-Kudo, Y.; Nishikawa, Y. Multiplex Real-Time PCR for Exhaustive Detection of Diarrhoeagenic *Escherichia coli. J. Appl. Microbiol.* **2009**, *106* (2), 410–420. <https://doi.org/10.1111/j.1365-2672.2008.04043.x>.

(3)	Boisen, N.; Scheutz, F.; Rasko, D. A.; Redman, J. C.; Persson, S.; Simon, J.; Kotloff, K. L.; Levine, M. M.; Sow, S.; Tamboura, B.; Toure, A.; Malle, D.; Panchalingam, S.; Krogfelt, K. A.; Nataro, J. P. Genomic Characterization of Enteroaggregative *Escherichia coli* from Children in Mali. *J. Infect. Dis.* **2012**, *205* (3), 431–444. <https://doi.org/10.1093/infdis/jir757>.

(4)	Liu, J.; Gratz, J.; Amour, C.; Nshama, R.; Walongo, T.; Maro, A.; Mduma, E.; Platts-Mills, J.; Boisen, N.; Nataro, J.; Haverstick, D. M.; Kabir, F.; Lertsethtakarn, P.; Silapong, S.; Jeamwattanalert, P.; Bodhidatta, L.; Mason, C.; Begum, S.; Haque, R.; Praharaj, I.; Kang, G.; Houpt, E. R. Optimization of Quantitative PCR Methods for Enteropathogen Detection. *PLoS One* **2016**, *11* (6), e0158199. <https://doi.org/10.1371/journal.pone.0158199>.

(5)	Thiem, V. D.; Sethabutr, O.; Von Seidlein, L.; Van Tung, T.; Canh, D. G.; Chien, B. T.; Tho, L. H.; Lee, H.; Houng, H. S.; Hale, T. L.; Clemens, J. D.; Mason, C.; Trach, D. D. Detection of *Shigella* by a PCR Assay Targeting the *ipaH* Gene Suggests Increased Prevalence of Shigellosis in Nha Trang, Vietnam. *J. Clin. Microbiol.* **2004**, *42* (5), 2031–2035. <https://doi.org/10.1128/JCM.42.5.2031-2035.2004>.

(6)	Ramos Moreno Ana Carolina; Gonçalves Ferreira, L.; Baquerizo Martinez, M. Enteroinvasive *Escherichia coli* vs. *Shigella flexneri*: How Different Patterns of Gene Expression Affect Virulence. *FEMS Microbiol. Lett.* **2009**, *301* (2), 156–163. <https://doi.org/10.1111/j.1574-6968.2009.01815.x>.

(7)	Verweij, J. J.; Blangé, R. A.; Templeton, K.; Schinkel, J.; Brienen, E. A. T.; Van Rooyen, M. A. A.; Van Lieshout, L.; Polderman, A. M. Simultaneous Detection of *Entamoeba histolytica*, *Giardia lamblia*, and *Cryptosporidium parvum* in Fecal Samples by Using Multiplex Real-Time PCR. *J. Clin. Microbiol.* **2004**, *42* (3), 1220–1223. <https://doi.org/10.1128/JCM.42.3.1220-1223.2004>.

(8)	Aprilianto E Wiria; Margaretta A Prasetyani; Firdaus Hamid; Linda J Wammes; Bertrand Lell; Iwan Ariawan; Hae Won Uh; Heri Wibowo; Yenny Djuardi; Sitti Wahyuni; Inge Sutanto; Linda May; Adrian JF Luty; Jaco J Verweij; Erliyani Sartono; Maria Yazdanbakhsh; Taniawati Supali. Does Treatment of Intestinal Helminth Infections Influence Malaria? Background and Methodology of a Longitudinal Study of Clinical, Parasitological and Immunological Parameters in Nangapanda, Flores, Indonesia (ImmunoSPIN Study). *BMC Infect. Dis.* **2010**, *10* (77), 1–12. <https://doi.org/https://doi.org/10.1186/1471-2334-10-77>.

(9)	Pholwat, S.; Liu, J.; Taniuchi, M.; Chinli, R.; Pongpan, T.; Thaipisutikul, I.; Ratanakorn, P.; Platts-Mills, J. A.; Fleece, M.; Stroup, S.; Gratz, J.; Mduma, E.; Mujaga, B.; Walongo, T.; Nshama, R.; Kimathi, C.; Foongladda, S.; Houpt, E. R. Genotypic Antimicrobial Resistance Assays for Use on *E. coli* Isolates and Stool Specimens. *PLoS One* **2019**, *14* (5), e0216747. <https://doi.org/10.1371/journal.pone.0216747>.

(10)	Chavda, K. D.; Satlin, M. J.; Chen, L.; Manca, C.; Jenkins, S. G.; Walsh, T. J.; Kreiswirth, B. N. Evaluation of a Multiplex PCR Assay to Rapidly Detect *Enterobacteriaceae* with a Broad Range of β-Lactamases Directly from Perianal Swabs. *Antimicrob. Agents Chemother.* **2016**, *60* (11), 6957–6961. <https://doi.org/10.1128/AAC.01458-16>.
